# Malaria treatment-seeking behaviour and its associated factors: A cross-sectional study in rural East Nusa Tenggara Province, Indonesia

**DOI:** 10.1101/2021.05.28.21258027

**Authors:** Robertus Dole Guntur, Jonathan Kingsley, Fakir M. Amirul Islam

## Abstract

**Introduction:** The World Health Organization recommends seeking medical treatment within 24 hours after transmission of malaria to reduce the risk of severe complications and its onwards spread. However, in some parts of Indonesia, including East Nusa Tenggara Province (ENTP), this adherence is not achieved for a range of reasons including delays in visiting health centres. This study aims to determine factors related to the poor understanding of appropriate malaria treatment-seeking behaviour (AMTSB) of rural adults in ENTP. AMTSB was defined as seeking treatment at professional health facilities within 24 hours of the onset of malaria symptoms.

**Methods:** A cross-sectional study was conducted in the East Sumba, Belu, and East Manggarai district of ENTP between October and December 2019. A multi-stage cluster sampling procedure was applied to enrol 1495 participants aged between 18 and 89 years old. Data were collected through face-to-face interview. Multivariable logistic regression analyses were used to assess significant factors associated with the poor understanding of AMTSB.

**Results:** 86% of participants were found to be familiar with the term malaria. However, poor understanding level of AMTSB in rural adults of ENTP achieved 60.4% with a 95% confidence interval (CI): 56.9 – 63.8. The most important factors associated with the poor understanding were lower levels of education, low socio-economic status (SES), occupation, and distance to the nearest health facilities. The poor understanding of AMTSB was significantly higher for adults with no education (adjusted odds ratio (AOR) 3.42, 95% CI: 1.81, 6.48); having low SES (AOR: 1.87, 95% CI: 1.19, 2.96); residing three km away from the nearest health facilities (AOR: 1.73, 95% CI: 1.2, 2.5); and working as housewife (AOR: 1.63, 95%CI: 1.01 – 2.63). Ethnicity, and family size were not associated with the poor understanding of AMTSB.

**Conclusion:** The proportion of rural adults having a poor understanding of AMTSB was high leading to ineffective implementation of artemisinin-based combination therapies as the method to treat malaria in ENTP. Improving awareness of AMTSB for rural adults having low-level education, low SES, working as a housewife, and living three km from the nearest health facilities is critical to support the efficacy of malaria treatment in ENTP, as well as to advance the Indonesian government’s objective to achieve malaria elimination by 2030.

## Introduction

The number of malaria cases in 2019 was estimated to be 229 million globally with a decrease of 4% in the last two decades [1]. This achievement was contributed to increasing coverage and access to various malaria control measures including the use of artemisinin-based combination therapies (ACTs) as the first-line method to treat malaria as recommended by the World Health Organization (WHO) [2-5]. The coverage of the use of ACTs increased significantly from 39% in the period from 2005 to 2011 to 81% in the period between 2015 and 2019 globally [1].

The effectiveness of this intervention depends greatly on various factors, including the behaviour of the community to seek timely treatment of the condition at health facilities [6-8]. An analysis of the malaria treatment-seeking behaviour database globally showed that treatment-seeking rates differ both within and among regions. For example, the rate of treatment-seeking behaviour of communities in the Southeast Asia Region (SEARO) at government facilities, which provide comprehensive diagnostic and treatment schedule for malaria [2,9] was only 27.6% (95% CI: 26.3 – 29.1), which is the lowest reported by WHO [10]. However, the rate of accessing any sources of treatment in SEARO communities was 78.8% (95%CI: 77.4 – 80.2), which was the highest proportion out of all countries reported in 2016 [10].

Time of treatment-seeking plays an important role in supporting the effectiveness of malaria management. The WHO recommends that treatment for malaria should occur within 24 hours of the onset of malaria symptoms to prevent the advancement of infection [2,5,11]. A recent systematic review on the impact of delaying in seeking malaria treatment indicates that the risk of severe malaria for patients seeking care more than 24 hours was higher than those seeking care less than 24 hours (odds ratio : 1.33, 95% CI: 1.07–1.64) [12]. Moreover, studies on various settings among Asia communities indicated that patients sought care for malaria at health facilities on average at least two days after the onset of symptoms [13-16]. One major reason of delaying treatment could be the use of traditional medicine in low to middle-income Asia-Pacific countries [17].

Indonesia remains the second largest contributor to the total number of malaria cases in South East Asia, with an estimate of 658,380 cases reported in 2019 [1]. Of this number, the most common species of malaria parasites were *Plasmodium falciparum* and *Plasmodium vivax* accounting for 32% and 20% cases respectively [1]. Malaria treatment applying ACTs has been in use in Indonesia since 2004 [18-20]. This medicine is available for free of charge in all government health facilities throughout Indonesia [19,20]. The coverage of malaria patients receiving ACTs to treat malaria has increased significantly over this period, especially in recent times, raised from 33.7% in 2013 [21] to 78.3% in 2018 [22]. This has been a part of the national effort to achieve malaria elimination by 2030 [19,20,23].

The high coverage of ACTs in Indonesia, however, is hampered by the behavior of people who treat their malaria without consultation from health practitioners. The prevalence of self-medication for malaria treatment among the Indonesian population accounted for 0.6% of the populace, with variation from 0.2% in Bali Province to 5.1% in West Papua Province [21]. Further to this, it has been found that 20% of adults among malaria affected people in high-risk malaria-endemic areas in the country applied for traditional medicine. This proportion was higher among poorer households [24].

A review of the current database of health-seeking behaviour of the Indonesian population notes overall that the Indonesian population tends to defer the seeking of health care until the disease has worsened [25]. To date, the majority of health-seeking behaviour research for communicable diseases including malaria has been conducted in the Western part of Indonesia. For the Eastern part of the country, including from the East Nusa Tenggara Province (ENTP), the literature is limited [25].

This is an omission given that ENTP, comprising of 16 main ethnicities [26,27] distributing across 624 islands in this archipelago province [28] is the province contributing the second largest malaria burden in Indonesia. The total number of positive cases reported in 2019 was 12,909 cases [23]. The proportion of malaria cases due to *Plasmodium vivax* in this province was higher compared to malaria cases because of *Plasmodium falciparum* [29,30].

Limited investigations into malaria treatment-seeking behaviour had been conducted in the ENTP [21,31-34]. One study drawing on a small sample at the sub-district level indicated that most participants sought care at health facilities after four days of the onset of symptoms [31]. Two population studies covering both rural and urban populations indicated a high prevalence of residents seeking a cure by self-medication, accounting for 2.7% [21]. This also saw a high prevalence of the local community buying malarial medicines at a kiosk without consultation with health professionals [33]. Another study covering a Tetun community in Timor islands indicated the prevalence of community members applying various local medicinal plants to treat malaria [32]. One study representing the rural population in the province level indicated that more than half of the population delayed in seeking malaria treatment [34]. However, factors associated with this poor understanding of AMTSB of rural communities have not been investigated yet. A better understanding of the factors contributed to the malaria health-seeking behaviour of the community has the potential to guide health policy makers in designing effective malaria treatment in the local context [35,36]. To date there is no study at population level on the factors associated with the poor understanding of AMTSB covering various ethnicities in rural population in the ENTP. To fill this gap, we conduct this study to investigate factors associated with the poor understanding of malaria treatment-seeking behaviour of rural community in the province to support national commitment to achieve zero local transmission by 2030.

## Methods

### Study area

A cross-sectional study was conducted in ENTP from October to December 2019. The province comprising of five main islands including Sumba, Flores, Timor, Alor, and Lembata has a total population of 5,456,203 with the ratio of males to females was 49.5% versus 50.5% [28]. Data collection was conducted in Sumba Island contributing to 70% of the total malaria cases in this province, Timor Island representing 10% of the total cases, and Flores island demonstrating 6% of the total malaria cases in this province [37].

Health care providers in this province are mainly supported by the government sector with state hospitals at district and province level and public health centre (PHC) at sub-district level, which is locally known as “Puskesmas”. When this research was carried out, the total number of PHC is 381 distributing across 309 sub-districts where the infrastructure in PHC is limited including no electricity in some PHCs [38]. The distribution of general practitioner (GP) in each PHC is uneven ranging from zero to ten with 47% of the total number of PCH was supported with one GP and 33% of them have no availability [38]. Following the guideline of the national malaria control program of the Indonesian government, the diagnosis of malaria cases in PHC was confirmed with laboratory tests or rapid diagnostic tests and the treatment for malaria with applied ACT as the first-line treatment [22,37]. Malaria patients caused by *Plasmodium falciparum* were treated with ACTs for three days and primaquine for one day, whilst malaria patients caused by *Plasmodium vivax* were treated with ACTs for 3 days and primaquine for fourteen days.

### Sample size and recruitment

The total sample was 1495 adults. This number was obtained after considering the prevalence of malaria in ENTP, the intra-class coefficient correlation for malaria prevention study in Indonesia, design effect, and participation rate of participants. The complete sample size calculation was presented previously [39].

A multi-stage random cluster sampling was conducted to obtain data from 49 clusters in the rural village of ENTP Indonesia. The description of sampling procedure was described previously [39]. Three districts were selected based on their malaria-endemic settings (MES), which were East Manggarai, Belu, and East Sumba district, representing as low, moderate, and high MES [40]. In each district, three sub-districts were selected randomly. The number of a cluster were selected in each selected district according to the size of the population. Since the size of each cluster was different, we selected 25 to 40 participants in each cluster proportion to the cluster size. The selection of this participants in each cluster applied a systematic random sampling method.

### Data collection

Data collection was conducted by local nurses who participated in a one-day intensive training in the capital city of selected districts before conducting the survey. The local language was used to interview participants face-to-face based on the questions that had been provided in a validated questionnaire. Data on socio-demographic characteristics and environmental variables were further collected. Access to a water tap in the household, and the ownership of items of durable assets including radio, television, hand phone, motorbike, bike, electricity, fridge, generator, tractor, car, and modern house were also gathered. These data were used to construct the social-economic status of participants. The comprehensive questionnaire has been published by the authors previously [39]. Three main questions as indicated in Table 1 were asked to participants to capture their understanding on malaria treatment-seeking behaviour.

**Table 1.**
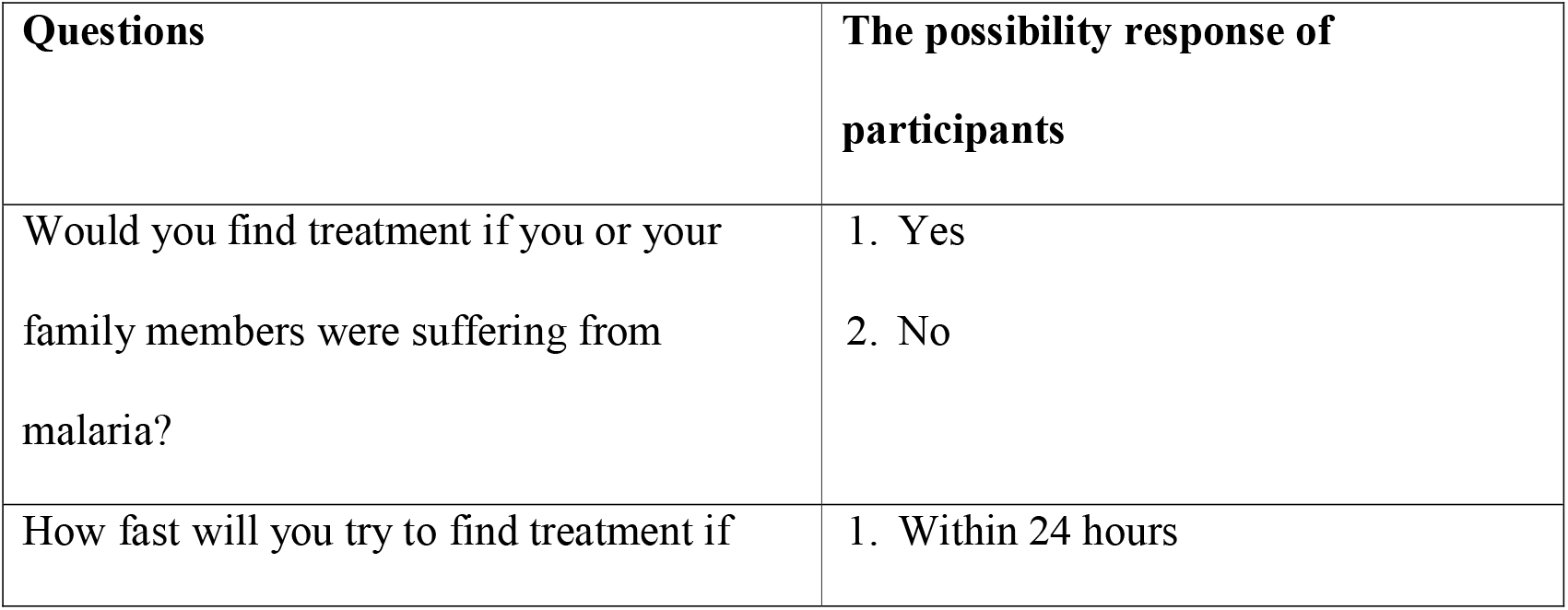
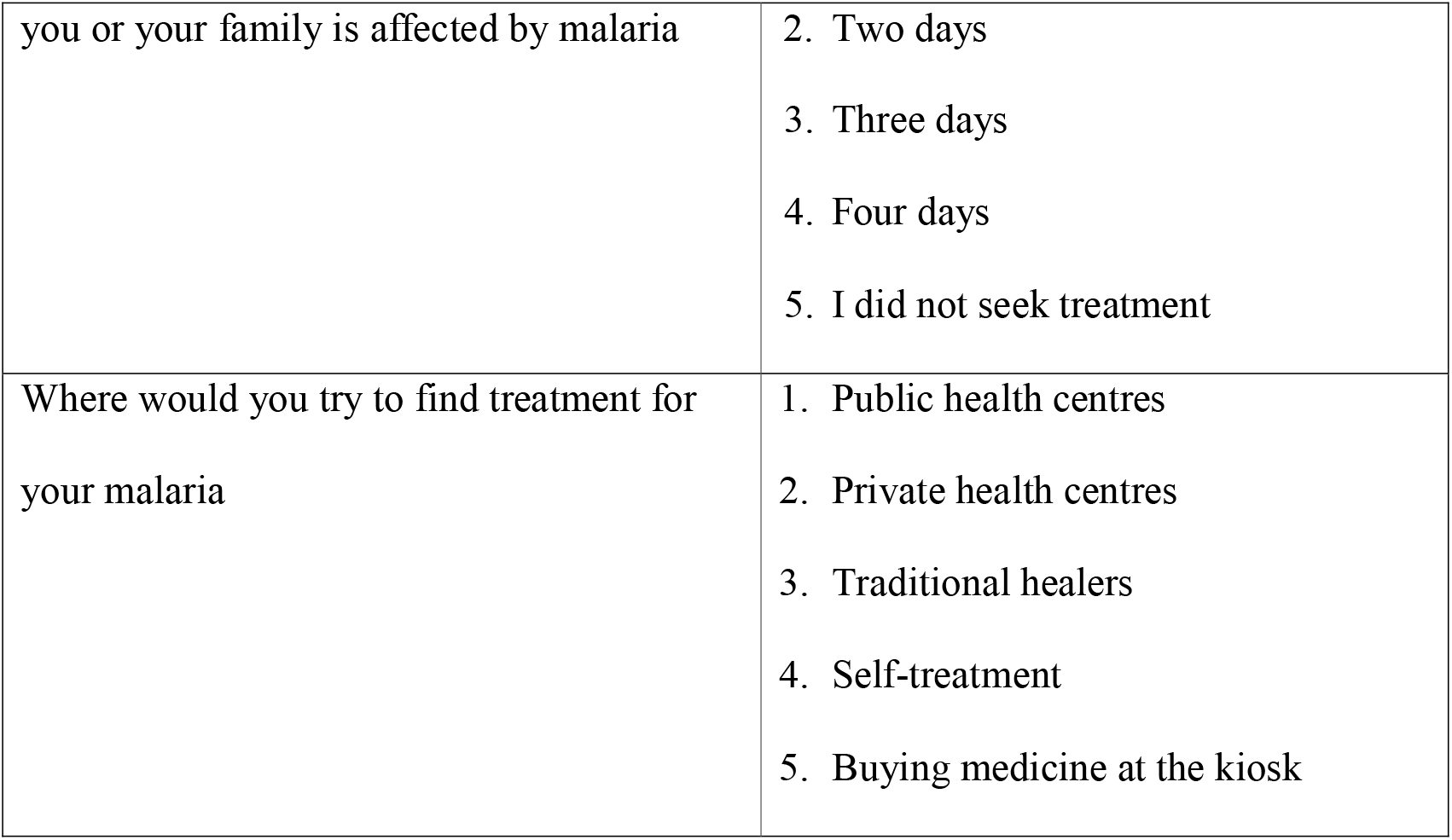
Questions used to explore understanding of treatment-seeking behaviour of participants.

### Outcome variables

There were three main outcomes of the study including perceptions on: (i) seeking malaria treatment after 24 hours, (ii) pursuing treatment at non-health facilities and, (iii) the poor understanding of appropriate malaria treatment-seeking behaviour (AMTSB). All these outcomes were dichotomous variables having categories yes which was marked as one or no that was marked as zero. The AMTSB was defined as seeking treatment at professional health facilities within 24 hours of the onset of the malaria symptom [2,41]. Based on the response of participants to the three main questions, they were categorized into three groups. The first group was the participants who sought treatment after 24 hours of detection of the malaria symptoms. The second group was the participants who sought treatment at non-health facilities. The third group comprised participants who sought treatment after 24 hours or at non-health facilities. All participants categorized as the third group was defined as having a poor understanding of AMTSB [42].

### Independent variables

Socio-demographic and environmental covariates that have shown the association with care-seeking malaria treatment were gender [42], age group [42-45]; education level [42,46,47]; social-economic status (SES) [46,48]; occupation [46]; income [49]; family size [43]; types of health facilities [41], distance to the health facilities [16,49,50]; and ethnicities [48,51]. In this study, gender was classified as males or females. The age group was divided into five categories: < 30 years, 30 – 40 years, 40 – 50 years, 50 – 60 years, and ≥ 60 years old. The education level of participants was classified as no education, primary education, junior education, senior education, and diploma or above education level. The main occupation of participants was categorized as a farmer, housewife, entrepreneur, government and non-government workers, and other occupations. The type of health facilities was classified as village maternity posts, village health posts, public health centres (PHC), and subsidiary PHC. The distance to the closest health facilities was defined as < 1 kilometres (km), 1 – 2 km, 2 – 3 km, ≥3 km. The ethnicities comprised Manggarai, Sumba, Timor, and other ethnicities. The SES of participants was classified as high, moderate, and low as indicated in the previous publication of the authors [34].

### Data Analysis

Participant’s characteristics including gender, age group, education level, main occupation, ethnicity, the nearest health service, and the distance to the nearest health service were analysed by descriptive statistics. The proportion of participants based on the first, second, and third outcome variables were tabulated based on the socio-demographic and environmental variables of participants. The association between outcomes and various independent variables was investigated by the chi-square test. Logistic regression methods were conducted to investigate the strengths of association of each independent variable with the outcome variables. The crude odds ratio (OR) and the adjusted OR with 95% confidence intervals were demonstrated as the result of logistic regression analysis. Any p values ≤ 0.05 was considered to be significant.

### Ethical consideration

This study was carried out in line with the principle of the Declaration of Helsinki to ensure that the rights, integrity, and privacy of participants were firmly treated. The ethical approval was obtained from the Swinburne University of Technology Human Ethics Committee with reference: 20191428-1490 and the Health Research Ethics Committee of Indonesia Health Ministry with reference: LB.02.01/2/KE.418/2019.

## Results

Of the total participants, 86% had heard malaria term, of which 88.7% were male, 82.6% finished their primary school, 88.1% were farmers, 82% were from low SES, and 84.1% were from a household with a family member of four or less. 13.9% (208) of participants have never heard of malaria. Of this number, 16.4% were female, 17.4% were completed their primary school, 21.1% were from Timor ethnicity, and 16.3% lived less than one kilometres from the nearest health facilities as shown in Fig 1.

**Fig 1.**
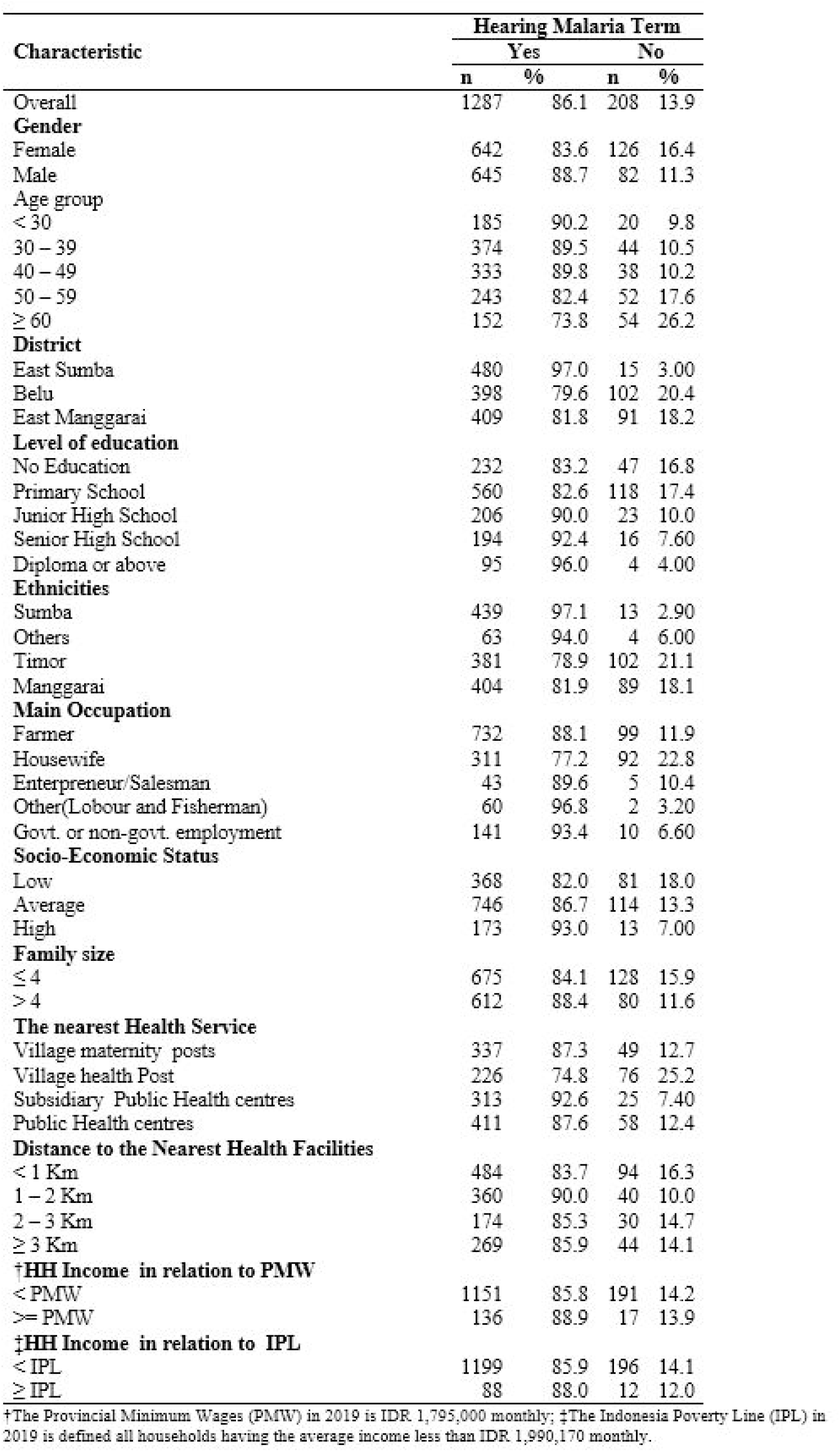
**Awareness of malaria and its association with the sociodemographic and environmental characteristics of respondents in the East Nusa Tenggara Province, Indonesia (n = 1495)**.

### Perception on finding malaria treatment

Perception of time and place to seek malaria treatment is presented in Table 2. Of 1287 participants hearing malaria term, almost all of them 98.8%, 95% confidence interval (CI): 98.2 – 99.4 reported to find malaria treatment if they or their family members suffering from malaria, and 82.4%, 95% CI: 80.2 – 84.7 sought treatment at public health facilities, however, only 46.8%, 95% CI: 42.8 – 50.8 of participants sought treatment within 24 hours of the onset of the malaria symptoms. Overall, 53.2%, 95% CI: 49.5 – 57.0, of participants had a perception to seek malaria treatment after 24 hours, and 15.7%, 95% CI: 10.7 – 20.7, believed to find treatment in non-health facilities. Overall, the percentage of participants having a poor understanding of AMTSB accounted for 60.4% with 95% CI: 56.9 – 63.8.

**Table 2.**
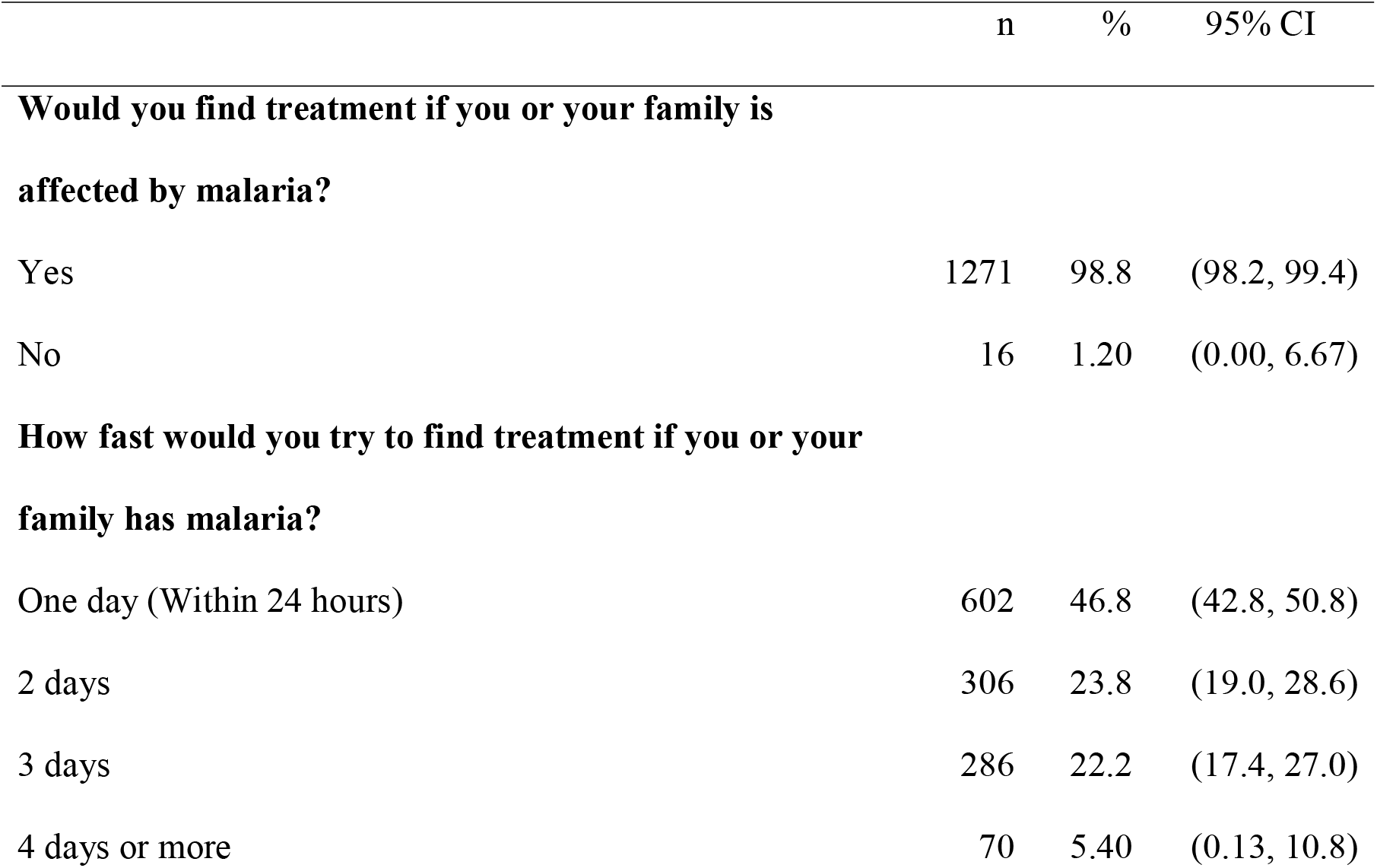
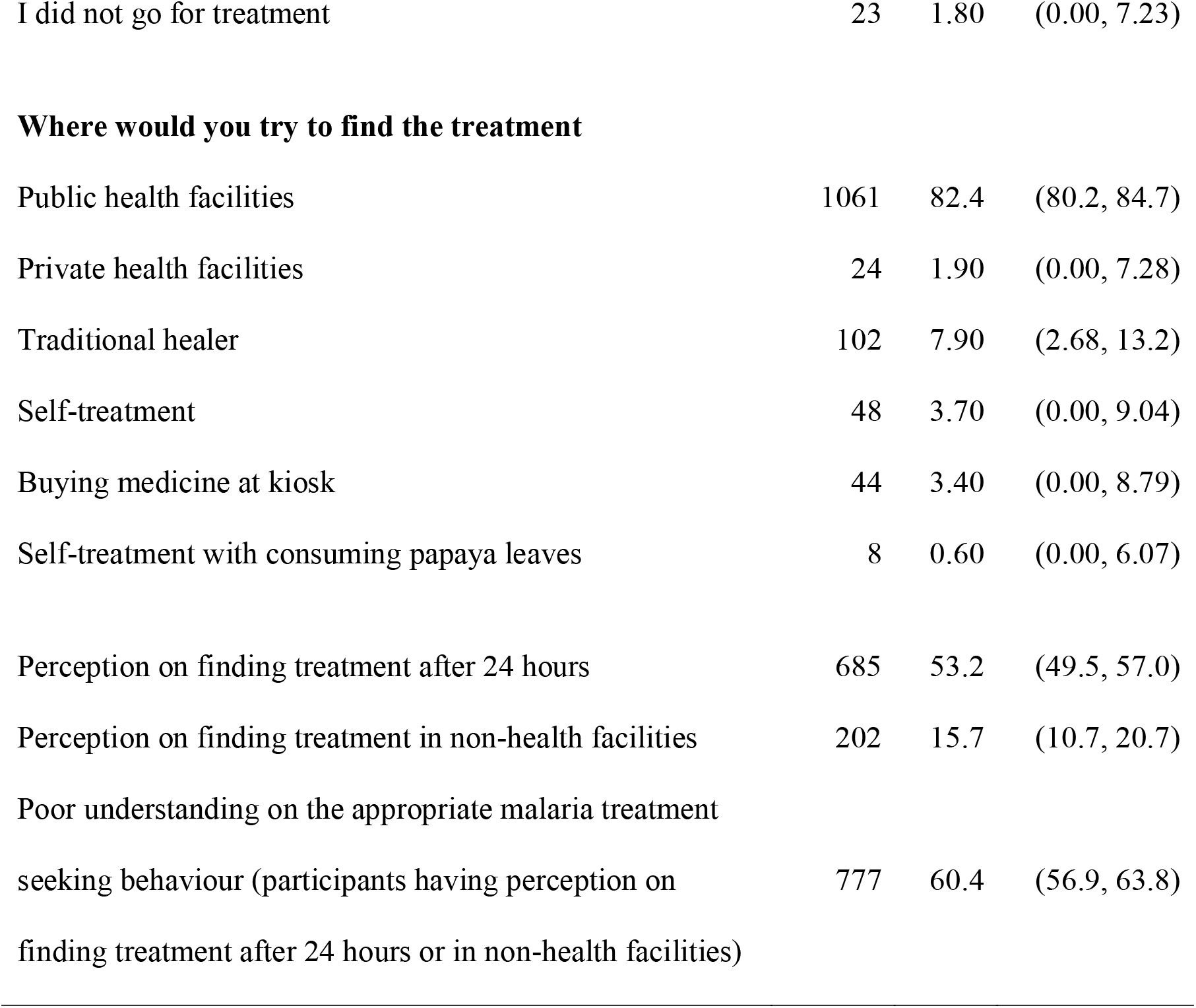
**Perception on finding treatment if respondents or their family members have any symptoms of malaria of people who were aware of malaria (n = 1287)**.

|  | n | % | 95% CI |
| --- | --- | --- | --- |
| **Would you find treatment if you or your family is affected by malaria?** |  |  |  |
| Yes | 1271 | 98.8 | (98.2, 99.4) |
| No | 16 | 1.20 | (0.00, 6.67) |
| **How fast would you try to find treatment if you or your family has malaria?** |  |  |  |
| One day (Within 24 hours) | 602 | 46.8 | (42.8, 50.8) |
| 2 days | 306 | 23.8 | (19.0, 28.6) |
| 3 days | 286 | 22.2 | (17.4, 27.0) |
| 4 days or more | 70 | 5.40 | (0.13, 10.8) |
| I did not go for treatment | 23 | 1.80 | (0.00, 7.23) |
| Where would you try to find the treatment |  |  |  |
| Public health facilities | 1061 | 82.4 | (80.2, 84.7) |
| Private health facilities | 24 | 1.90 | (0.00, 7.28) |
| Traditional healer | 102 | 7.90 | (2.68, 13.2) |
| Self-treatment | 48 | 3.70 | (0.00, 9.04) |
| Buying medicine at kiosk | 44 | 3.40 | (0.00, 8.79) |
| Self-treatment with consuming papaya leaves | 8 | 0.60 | (0.00, 6.07) |
| Perception on finding treatment after 24 hours | 685 | 53.2 | (49.5, 57.0) |
| Perception on finding treatment in non-health facilities | 202 | 15.7 | (10.7, 20.7) |
| Poor understanding on the appropriate malaria treatment seeking behaviour (participants having perception on finding treatment after 24 hours or in non-health facilities) | 777 | 60.4 | (56.9, 63.8) |

### Seeking malaria treatment beyond 24 hours and its factors associated

The distribution of participants seeking malaria treatment after 24 hours and their factors associated were presented in Fig 2. Overall, 53.2% of participants had the perception to seek malaria treatment after 24 hours. Compared to 44.5% of participants living closer to public health centres, 63.7% of participants resided closer to village health posts had the perception to seek malaria treatment after 24 hours. The highest percentage of seeking malaria treatment beyond 24 hours was in participants with no education level (69.8%), whilst the lowest was in participants with a diploma or above education level (27.4%).

**Fig 2.**
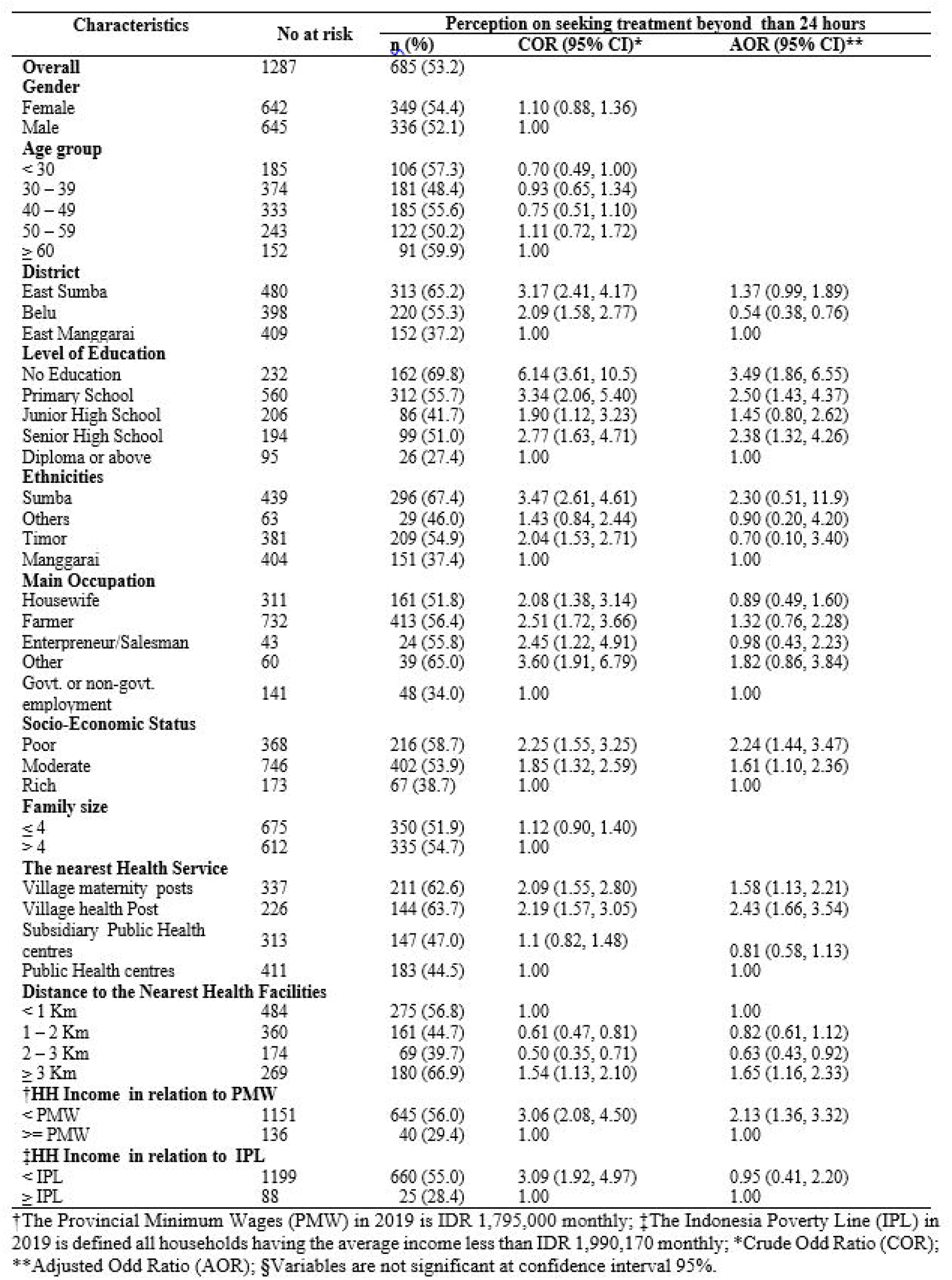
**The level of perception on seeking malaria treatments beyond 24 hours by different sociodemographic and environmental characteristics in the East Nusa Tenggara Province, Indonesia (n=685)**.

After adjustment for gender and age group, the following variables were significantly associated with the perception of seeking malaria treatment after 24 hours: no education level compared to those with a diploma or above education level (Adjusted Odds ratio (AOR) 3.49, 95% confidence interval (CI) 1.86 – 6.55,); poor SES compared to those were from high SES (AOR: 2.24, 95% CI: 1.44 – 3.47,); village health post compared to those were living close to the public health centre (AOR: 2.43, 95% CI: 1.66 – 3.54); living more than three kilometres compared to those were living less than one kilometres from health facilities (AOR: 1.65, 95% CI: 1.16 – 2.33); household income less than provincial minimum wages (PMW) compared to those were from the household with income more than PMW (AOR: 2.13, 95%CI: 1.36 – 3.32).

### Seeking malaria treatment at non-health facilities and its factors associated

The distribution of participants seeking malaria treatment at non-health facilities and their factors associated was presented in Fig 3. Of the total participants hearing malaria term, 15.7% had the perception to seek malaria treatment at non-health facilities. Compared to 11.7% of participants living closer to public health centres, 20.8% among those residing closer to village maternity post had the perception to seek malaria treatment at non-health facilities. The percentage of seeking malaria treatment at non-health facilities for those having no education was the highest (19.8%) compared to only 5.30% for those having a diploma or above education level.

**Fig 3.**
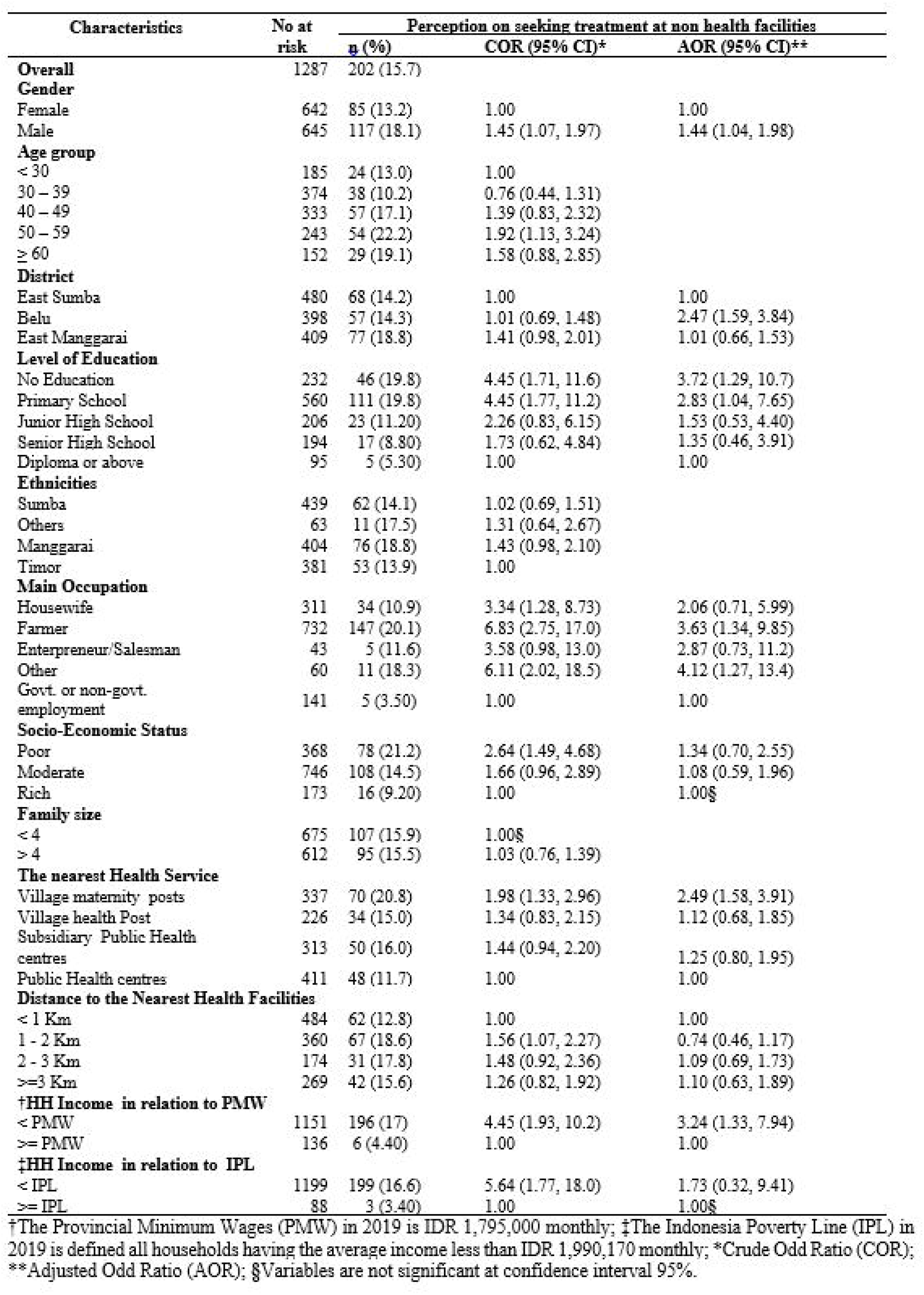
**The level of perception on seeking malaria treatment in non-health facilities by different sociodemographic and environmental characteristics in the East Nusa Tenggara Province, Indonesia (n=202)**.

After adjustment for gender and age group, the following variables were significantly associated with the perception of seeking malaria treatment at non-health facilities: male compared to female (Adjusted Odds ratio (AOR) 1.44, 95% confidence interval (CI) 1.04 – 1.98); Belu district compared to East Sumba district (AOR: 2.47, 95% CI: 1.59 – 3.84); no education level compared to those with a diploma or above education level (AOR: 3.72, 95% CI: 1.29 – 10.72); farmer compared to those with government or non-government workers (AOR: 3.63, 95% CI: 1.34 – 9.85); village maternity post compared to those were living close to the public health centre (AOR: 2.49, 95% CI: 1.58 – 3.91); household income less than provincial minimum wages (PMW) compared to those were from the household with income more than PMW (AOR: 3.24, 95%CI: 1.33 – 7.94).

### The poor understanding of appropriate malaria treatment-seeking behaviour (AMTSB) and its factors associated

The distribution of the poor understanding of AMTSB and its factors associated was presented in Fig 4. Overall, 60.4% of participants hearing malaria term had a poor understanding of AMTSB. The proportion of poor understanding was the highest (73.7%) for those having no education compared to those with a diploma or above education level (31.6%). 70.7% of participants with poor SES compared to 42.8% of participants with rich SES were found to have a poor understanding of AMTSB. More than half of participants in all ethnicities in the ENTP had a poor understanding of AMTSB with the highest in Sumba ethnicity (69.9%) and the lowest in other ethnicities (50.8%).

**Fig 4.**
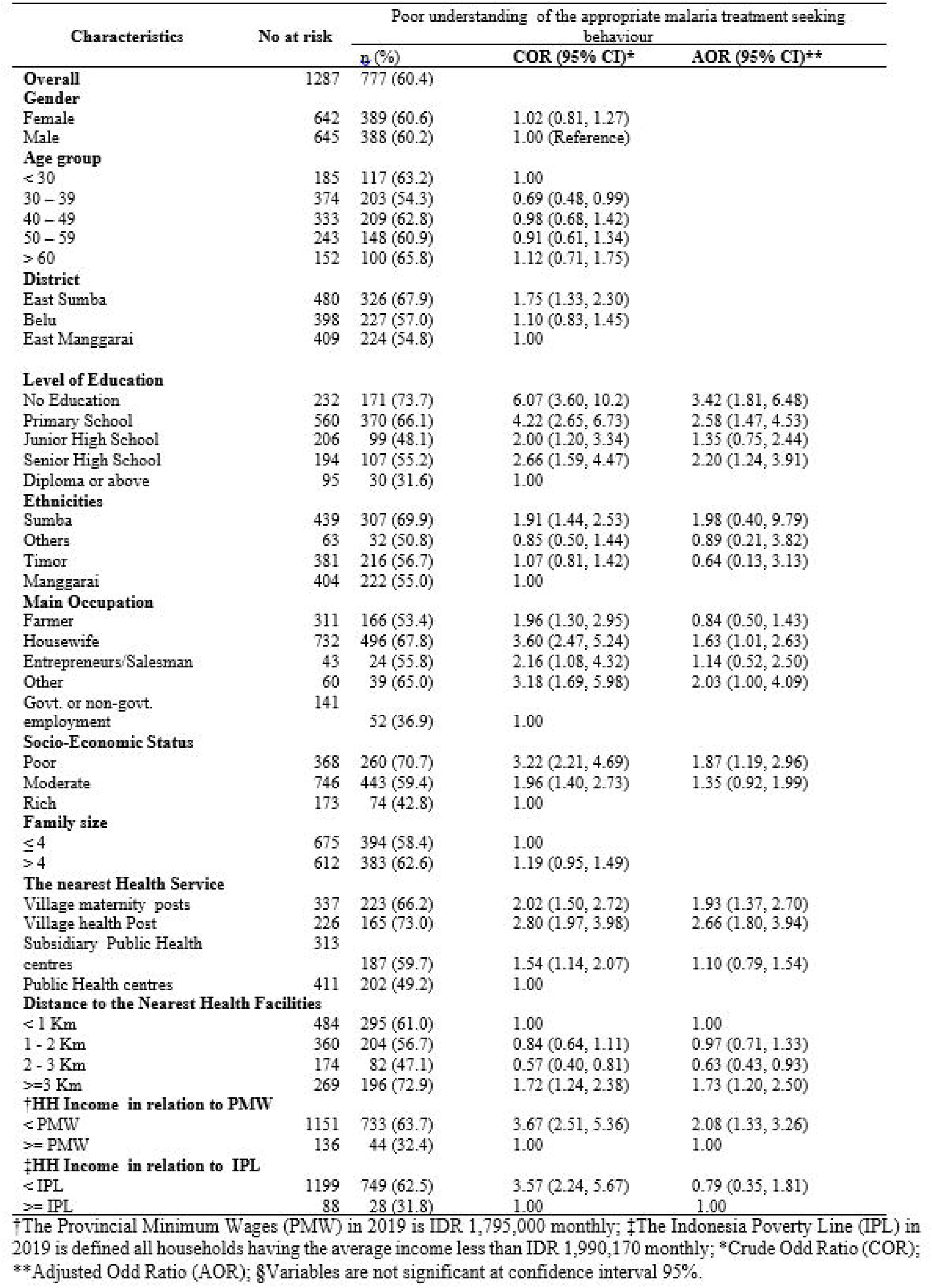
**The level of poor understanding of appropriate treatment-seeking behaviour of malaria by different sociodemographic and environmental characteristics in the East Nusa Tenggara Province, Indonesia (n=777)**.

After adjustment for gender and age group, the following variables were significantly associated with the poor understanding of AMTSB for rural adults in ENTP: no education level compared to those with a diploma or above education level (Adjusted Odds ratio (AOR) 3.42, 95% confidence interval (CI) 1.81 – 6.48); household income less than provincial minimum wages (PMW) compared to those were from the household with income more than PMW (AOR: 2.08, 95%CI: 1.33 – 3.26); poor SES compared to rich SES (AOR: 1.87, 95% CI: 1.19 – 2.96); village health post compared to those were living close to the public health centre (AOR: 2.66, 95% CI: 1.80 – 3.94); participants living at least three kilometres away from the nearest health facilities compared to those living within one kilometre from health facilities (AOR: 1.73, 95% CI: 1.2 – 2.5); occupation as housewife compared to those working at the government or non-government sector (AOR : 1.63, 95%CI: 1.01 – 2.63).

## Discussion

This study addresses a research gap around understanding of AMTSB and its associated factors in rural communities in Indonesia. The main finding of this study shows that more than half of rural communities in ENTP have a perception to delay in malaria treatment-seeking behaviour and more than half of them have a poor understanding of AMTSB which leads to a significant obstacle to address malaria. The main factors associated with the poor understanding of the AMTSB were the low level of education, low social-economic status, working as housewife, and distance to the nearest health facilities.

This study shows that more than half of participants delay in seeking treatment for their malaria. This finding is consistent with studies in Myanmar [15], Nigeria [43], Uganda [52], and South-Eastern Nigeria [53]. However, our finding contrasted with studies in Cabo Verde [54] and South Africa [55], revealing that the high awareness of the community to seek treatment within 24 hours. The low awareness to seek treatment within 24 hours leads to ineffective implementation of ACTs as the first line of malaria treatment, even though the coverage of ACTs in ENTP has increased from 55% in 2013 [21] to 83% in 2018 [22]. The effectiveness of ACTs as the first-line treatment for malaria will occur when the treatment has been initiated within 24 hours of malaria [2]. This finding indicates that it is critical to improve the awareness of the rural population in this province to seek treatment promptly since delay in seeking treatment leads to adverse effects for both participants and the community. For an individual, seeking treatment after 24 hours increases their possibility for severe malaria anemia, blood transfusion [12], hospital inpatients [16], and mortality rate [56]. At the community level, delay in seeking treatment increased the chance of onward transmission of the diseases to the community [11,57].

The study also shows that a high proportion of participant has a perception in seeking malaria treatment at non-health facilities. This finding was consistent with a study in India [51] Bangladesh [58], Ghana [59], and Turkey [60], however, it was contrasted with finding in Saudi Arabia [61] and Zambia [62] indicating that a high proportion of community to seek treatment at health facilities. Raising awareness at the community level to seek treatment at health facilities is crucial to ensure all clinical cases of malaria could be examined accurately under microscopies for allowing health practitioners to identify the types of malaria to provide treatment accordingly [63]. The study shows that participants with no education level were almost four times higher to seek malaria treatment at non-health facilities compared to those having a diploma or above. This is more likely that non educated people was associated with low health literacy [64], therefore they do not know the right place to diagnosis their disease leads to the low rate in the use of professional health service as a highlight in other studies [65].

The present study demonstrated that the low level of education was significantly associated with the poor understanding of AMTSB. This study shows that adults with no education were three times higher to have poor understanding compared to adults with a diploma or above education level. These results were consistent with other studies in other countries such as Myanmar [42], India [46], and Northern Ethiopia [47]. This is more likely adults with a low level of education have poor knowledge on when and where they have to seek treatment for malaria since they have no ability to understand various written health information to improve their health behaviour. This situation was worst with another finding of this study indicating that 34.2% of participants who never hear malaria term were from a low level of education. Therefore, it could be assumed that most rural adults in this province have a poor understanding of AMTSB regardless of whether they hear or never heard of malaria term. This situation leads to worst the management of malaria treatment which causes by *Plasmodium* vivax which in turn hinders the effort to achieve malaria elimination in this province.

This study shows further that distance to the nearest health facilities is one of the determinant factors associated with a poor understanding of AMTSB. The study has shown that the poor understanding of adults residing at least 3 kilometres from the nearest health services was almost two times higher compared to those living less than one kilometres. Our finding corroborates with the studies in Myanmar [49], Laos [66], India [16], Ethiopia [67], Northwest Ethiopia [68], Equatorial Guinea [69], and Tanzania [50]. This might be because people living far from health facilities were less exposure with malaria information since they were struggle to access to health facilities. Literature indicated public transports in village were limited and people fully rely on foot for accessing health facilities in rural area [70].

The study has identified the poor understanding of malaria treatment-seeking behaviour of rural communities in ENTP. Improving the awareness of this community to seek treatment effectively is fundamental to be addressed by the local authority to ensure universal access to malaria diagnosis and effective treatment, which is one of the global strategies recommended by WHO to achieve malaria elimination by 2030 [71]. Furthermore, this study has provided strong evidence that rural adults with a low level of education are the most vulnerable groups having a poor understanding of AMTSB. Considering of poor malaria awareness and the low level of education of the rural population in the ENTP [28,34,72] special attention of the local authority should be focused on this vulnerable group to improve their awareness of AMTSB.

The strength of the study include that it was conducted based on a large sample size representing the rural population of three main ethnicities from three main islands of ENTP and three different malaria-endemic areas allowing authors to capture a different perspective of the local community on how their understanding of AMTSB. Furthermore, data collection supported by local nurses was conducted by face-to-face interview to accommodate the low literacy of the rural population in the province. Despite these, the study has a number of limitations. The treatment-seeking behaviour was assessed based on the hypothetical scenario. The participants were asked their opinion on when and where they would seek treatment if they or their family members experienced malaria. The hypothetical behaviour might be different from actual behaviour. However, this kind of treatment-seeking understanding is usually used to present variation of malaria treatment-seeking behaviour amongst demographic groups in the rural population.

## Conclusion

More than half of rural adults in this study have a poor understanding of appropriate malaria treatment-seeking behaviour. This situation leads to poor implementation of ACTs to treat malaria and the completeness of malaria treatment. Rural adults having low-level education, low SES, working as a housewife, and living more than three kilometres from the nearest public health facilities were the most vulnerable groups that should be prioritized by policy health markers. The improvement of AMTSB for these groups will support the efficacy of malaria treatment in ENTP, as well as work to eliminate malaria globally.

## Data Availability

All data supporting this article have been provided with this article as supporting information

## Acknowledgments

We would like to thank the Australia Awards Scholarship for supporting this research and the Faculty of Health, Arts, and Design of the Swinburne University Technology for providing funding for primary data collection for this study. The funders had no role in the designing of the study, data collection, analysis, or interpretation of data, or writing the paper. We further would like to express our gratitude to the Health Ministry of Indonesia, the governor of ENTP, head of East Sumba, Belu, and East Manggarai district, nine head of sub-districts, and forty-nine village leaders for allowing conducted this research in their region.

## Supporting Information

S1 Dataset: Database for study malaria treatment-seeking behaviour in rural East Nusa Tenggara Province, Indonesia.

## References

1. World Health Organization. World Malaria Report 2020: 20 years of global progress and challenges. Geneva World Health Organization; 2020.

2. World Health Organization. Guidelines For The Treatment of Malaria. Third ed. Geneva: World Health Organization; 2015.

3. Björkman A, Shakely D, Ali AS, Morris U, Mkali H, Abbas AK, et al. From high to low malaria transmission in Zanzibar—challenges and opportunities to achieve elimination. BMC Med. 2019;17:14. doi: https://doi.org/10.1186/s12916-018-1243-z.

4. Goldlust SM, Thuan PD, Giang DDH, Thang ND, Thwaites GE, Farrar J, et al. The decline of malaria in Vietnam, 1991-2014. Malar J. 2018;17:226. doi: 10.1186/s12936-018-2372-8. PubMed PMID: 29880051.

5. Bhatt S, Weiss DJ, Cameron E, Bisanzio D, Mappin B, Dalrymple U, et al. The effect of malaria control on Plasmodium falciparum in Africa between 2000 and 2015. Nature. 2015;526(7572):207–211. doi: 10.1038/nature15535. PubMed PMID: 26375008.

6. The malERA Consultative Group on Health Systems and Operational Research. A Research Agenda for Malaria Eradication: Health Systems and Operational Research. PLoS Med. 2011;8(1):e1000397. doi: 10.1371/journal.pmed.1000397.

7. Rao VB, Schellenberg D, Ghani AC. Overcoming health systems barriers to successful malaria treatment. Trends Parasitol. 2013;29(4):164-180. doi: https://doi.org/10.1016/j.pt.2013.01.005.

8. Littrell M, Miller JM, Ndhlovu M, Hamainza B, Hawela M, Kamuliwo M, et al. Documenting malaria case management coverage in Zambia: a systems effectiveness approach. Malar J. 2013;12(1):371. doi: 10.1186/1475-2875-12-371.

9. World Health Organization. World malaria report 2014. Geneva: World Health Organization; 2014.

10. Battle KE, Bisanzio D, Gibson HS, Bhatt S, Cameron E, Weiss DJ, et al. Treatment-seeking rates in malaria endemic countries. Malar J. 2016;15:20. doi: 10.1186/s12936-015-1048-x. PubMed PMID: PMC4709965.

11. Challenger JD, Gonçalves BP, Bradley J, Bruxvoort K, Tiono AB, Drakeley C, et al. How delayed and non-adherent treatment contribute to onward transmission of malaria: a modelling study. BMJ Glob Health. 2019;4(6):e001856. doi: 10.1136/bmjgh-2019-001856.

12. Mousa A, Al-Taiar A, Anstey NM, Badaut C, Barber BE, Bassat Q, et al. The impact of delayed treatment of uncomplicated P. falciparum malaria on progression to severe malaria: A systematic review and a pooled multicentre individual-patient meta-analysis. PLoS Med. 2020;17(10):e1003359. doi: 10.1371/journal.pmed.1003359.

13. Yadav SP, Yadav S, Kuma P, Yadav S. Knowledge, treatment-seeking behaviour and socio-economic impact of malaria in the desert of Rajasthan, India. Southern African Journal of Epidemiology and Infection. 2013;28(1):41–47. doi: 10.1080/10158782.2013.11441518.

14. Yasuoka J, Kikuchi K, Nanishi K, Ly P, Thavrin B, Omatsu T, et al. Malaria knowledge, preventive actions, and treatment-seeking behavior among ethnic minorities in Ratanakiri Province, Cambodia: a community-based cross-sectional survey. BMC Public Health. 2018;18(1):1206–1206. doi: 10.1186/s12889-018-6123-0. PubMed PMID: 30367615.

15. Than MM, Myo M, Pyae LA. The Determinants of Delayed Diagnosis and Treatment Among Malaria Patients in Myanmar: A Cross-Sectional Study. The Open Public Health Journal. 2019;12. doi: 10.2174/1874944501912010078.

16. Chaturvedi HK, Bajpai RC, Tiwari P. Determination of cut-off and correlates of delay in treatment-seeking of febrile illness: a retrospective analysis. BMC Public Health. 2020;20(1):572. doi: 10.1186/s12889-020-08660-2.

17. Suswardany DL, Sibbritt DW, Supardi S, Chang S, Adams J. A critical review of traditional medicine and traditional healer use for malaria and among people in malaria-endemic areas: contemporary research in low to middle-income Asia-Pacific countries. Malar J. 2015;14:98. doi: 10.1186/s12936-015-0593-7. PubMed PMID: 25889412.

18. Rajendran J. Artemisinin-Based Therapy for Malaria. American Journal of Medicine and Medical Sciences 2013;3(4): 81–90. doi: 10.5923/j.ajmms.20130304.05.

19. Sitohang V, Sariwati E, Fajariyani SB, Hwang D, Kurnia B, Hapsari RK, et al. Malaria elimination in Indonesia: halfway there. Lancet Glob Health. 2018;6(6):e604–e606. doi: 10.1016/S2214-109X(18)30198-0.

20. Kusriastuti R, Surya A. New treatment policy of malaria as a part of malaria control program in Indonesia. Acta Medica Indonesiana - The Indonesian Journal of Internal Medicine. 2012;44(3):265–269.

21. Indonesia Health Ministry. Basic Health Research Riskesdas 2013 Jakarta: Agency of Health Research and Development Ministry of Health of Republic of Indonesia; 2013.

22. Indonesia Ministry of Health. National Report of Basic Health Research 2018. Jakarta: Ministry of Health of The Republic of Indonesia; 2019.

23. Indonesia Ministry of Health. The current development situation of malaria control program in Indonesia 2019. Jakarta: Ministry of Health of The Republic of Indonesia; 2020.

24. Suswardany DL, Sibbritt DW, Supardi S, Pardosi JF, Chang S, Adams J. A cross-sectional analysis of traditional medicine use for malaria alongside free antimalarial drugs treatment amongst adults in high-risk malaria endemic provinces of Indonesia. PLoS One. 2017;12(3):e0173522–e0173522. doi: 10.1371/journal.pone.0173522. PubMed PMID: 28329019.

25. Widayanti AW, Green JA, Heydon S, Norris P. Health-Seeking Behavior of People in Indonesia: A Narrative Review. J Epidemiol Glob Health. 2020;10(1):6–15. doi: 10.2991/jegh.k.200102.001. PubMed PMID: 32175705.

26. Subanpulo OSW. Pengaruh Budaya Lamaholot Dalam Ruang Kota Larantuka. Jurnal Pembangunan Wilayah dan Kota. 2012;8:247–256.

27. Yuliawati S. Measurement of social cultural gatra in East Nusa Tenggara Province Indonesia [in Indonesian]. Journal of Educational Research and Evaluation. 2011;15:139–154.

28. The Central Bureau of Statistics East Nusa Tenggara Province. East Nusa Tenggara Province (ENTP) in Figures 2020 Kupang: The Central Bureau of Statistics, ENTP; 2020.

29. Kosasih A, Koepfli C, Dahlan MS, Hawley WA, Baird JK, Mueller I, et al. Gametocyte carriage of Plasmodium falciparum (pfs25) and Plasmodium vivax (pvs25) during mass screening and treatment in West Timor, Indonesia: a longitudinal prospective study. Malar J. 2021;20(1):177. doi: 10.1186/s12936-021-03709-y.

30. Hutagalung J, Kusnanto H, Supargiyono S, Sadewa A, Aw S, Novianti R, et al. Malaria pre-elimination assessment in The Eastern Indonesia. Outbreak Surveill Investig Rep. 2016;9:1–7.

31. Karolus N, Oktafianus S. Environment and Public Behaviour Factor about Malaria in East Kupang Subdistrict Kupang District ENTP Indonesia. National Public Health Journal 2013;7(6):271 –278.

32. Taek MM, Banilodu L, Neonbasu G, Watu YV E W BP, Agil M. Ethnomedicine of Tetun ethnic people in West Timor Indonesia; philosophy and practice in the treatment of malaria. Integr Med Res. 2019;8(3):139–144. doi: 10.1016/j.imr.2019.05.005. PubMed PMID: 31304086.

33. Ipa M, Dhewantara PW. Treatment variation of malaria at household-level in six endemic regions in Indonesia. J Aspirator Journal of Vector-Borne Diseases. 2015;7(1):13–22.

34. Guntur RD, Kingsley J, Islam FMA. Malaria awareness of adults in high, moderate and low transmission settings: A cross-sectional study in rural East Nusa Tenggara Province, Indonesia. medRxiv. 2021:2020.2012.2004.20243675. doi: 10.1101/2020.12.04.20243675.

35. Shaikh B. Understanding social determinants of health seeking behaviours, providing a rational framework for health policy and systems development. J Pak Med Assoc. 2008;58:33–36.

36. Abuduxike G, Aşut Ö, Vaizoğlu SA, Cali S. Health-Seeking Behaviors and its Determinants: A Facility-Based Cross-Sectional Study in the Turkish Republic of Northern Cyprus. Int J Health Policy Manag. 2020;9(6):240–249. doi: 10.15171/ijhpm.2019.106.

37. Health Department of the ENTP. The East Nusa Tenggara Province (ENTP) Health Profile in 2018 Indonesia Kupang: Health Department of the ENTP, Indonesia; 2019.

38. Indonesia Health Ministry. Basic database of Public Health Centre in East Nusa Tenggara Province Indonesia, Condition at 31 December 2018. Jakarta: Indonesia Health Ministry; 2019.

39. Guntur RD, Kingsley J, Islam FMA. Epidemiology of Malaria in East Nusa Tenggara Province in Indonesia: Protocol for a Cross-sectional Study. JMIR Res Protoc. 2021;10(4):e23545. doi: 10.2196/23545.

40. Indonesia Health Ministry. Indonesia’s Health Profile 2018. Jakarta: Ministry of Health of the Republic of Indonesia; 2019.

41. Thandar MM, Kyaw MP, Jimba M, Yasuoka J. Caregivers’ treatment-seeking behaviour for children under age five in malaria-endemic areas of rural Myanmar: a cross-sectional study. Malar J. 2015;14(1):1. doi: 10.1186/1475-2875-14-1.

42. Naing PA, Maung TM, Tripathy JP, Oo T, Wai KT, Thi A. Awareness of malaria and treatment-seeking behaviour among persons with acute undifferentiated fever in the endemic regions of Myanmar. Trop Med Health. 2017;45:31. doi: 10.1186/s41182-017-0070-9. PubMed PMID: 29213208.

43. Babalola OJ, Ajumobi O, Ajayi IO. Rural-urban disparities and factors associated with delayed care-seeking and testing for malaria before medication use by mothers of under-five children, Igabi LGA, Kaduna Nigeria. Malar J. 2020;19(1):294–294. doi: 10.1186/s12936-020-03371-w. PubMed PMID: 32811529.

44. Adinan J, Damian DJ, Mosha NR, Mboya IB, Mamseri R, Msuya SE. Individual and contextual factors associated with appropriate healthcare seeking behavior among febrile children in Tanzania. PLoS One. 2017;12(4):e0175446. doi: 10.1371/journal.pone.0175446.

45. Mitiku I, Assefa A. Caregivers’ perception of malaria and treatment-seeking behaviour for under five children in Mandura District, West Ethiopia: a cross-sectional study. Malar J. 2017;16(1):144. doi: 10.1186/s12936-017-1798-8.

46. Singh MP, Saha KB, Chand SK, Savargaonkar D. Socioeconomic determinants of community knowledge and practice in relation to malaria in high-and low-transmission areas of central India. J Biosoc Sci. 2020;52(3):317–329. doi: 10.1017/S0021932019000440.

47. Tesfahunegn A, Zenebe D, Addisu A. Determinants of malaria treatment delay in northwestern zone of Tigray region, Northern Ethiopia, 2018. Malar J. 2019;18(1):358. doi: 10.1186/s12936-019-2992-7.

48. Karyana M, Devine A, Kenangalem E, Burdarm L, Poespoprodjo JR, Vemuri R, et al. Treatment-seeking behaviour and associated costs for malaria in Papua, Indonesia. Malar J. 2016;15:536. doi: 10.1186/s12936-016-1588-8.

49. Xu JW, Xu QZ, Liu H, Zeng YR. Malaria treatment-seeking behaviour and related factors of Wa ethnic minority in Myanmar: a cross-sectional study. Malar J. 2012;11:417. doi: 10.1186/1475-2875-11-417. PubMed PMID: 23237576.

50. Kassile T, Lokina R, Mujinja P, Mmbando BP. Determinants of delay in care seeking among children under five with fever in Dodoma region, central Tanzania: a cross-sectional study. Malar J. 2014;13:348–348. doi: 10.1186/1475-2875-13-348. PubMed PMID: 25182432.

51. Singh MP, Saha KB, Chand SK, Anvikar A. Factors associated with treatment seeking for malaria in Madhya Pradesh, India. Trop Med Int Health. 2017;22(11):1377–1384. doi: 10.1111/tmi.12973.

52. Rutebemberwa E, Kallander K, Tomson G, Peterson S, Pariyo G. Determinants of delay in care-seeking for febrile children in eastern Uganda. Trop Med Int Health. 2009;14(4):472–479. doi: 10.1111/j.1365-3156.2009.02237.x. PubMed PMID: 19222823.

53. Chukwuocha UM, Okpanma AC, Nwakwuo GC, Dozie INS. Determinants of Delay in Seeking Malaria Treatment for Children Under-Five Years in Parts of South Eastern Nigeria. Journal of Community Health. 2014;39(6):1171–1178. doi: 10.1007/s10900-014-9872-4.

54. DePina AJ, Dia AK, de Ascenção Soares Martins A, Ferreira MC, Moreira AL, Leal SV, et al. Knowledge, attitudes and practices about malaria in Cabo Verde: a country in the pre-elimination context. BMC Public Health. 2019;19(1):850. doi: 10.1186/s12889-019-7130-5.

55. Manana PN, Kuonza L, Musekiwa A, Mpangane HD, Koekemoer LL. Knowledge, attitudes and practices on malaria transmission in Mamfene, KwaZulu-Natal Province, South Africa 2015. BMC Public Health. 2018;18(1):41–41. doi: 10.1186/s12889-017-4583-2. PubMed PMID: 28728572.

56. Giao PT, de Vries PJ, Binh TQ, Nam NV, Kager PA. Early diagnosis and treatment of uncomplicated malaria and patterns of health seeking in Vietnam. Trop Med Int Health. 2005;10(9):919-925. doi: https://doi.org/10.1111/j.1365-3156.2005.01472.x.

57. Chaturvedi HK, Mahanta J, Bajpai RC, Pandey A. Risk of malaria among febrile patients: retrospective analysis of a hospital-based study in an endemic area of northeast India. International Health. 2014;6(2):144–151.

58. Ahmed SM, Haque R, Haque U, Hossain A. Knowledge on the transmission, prevention and treatment of malaria among two endemic populations of Bangladesh and their health-seeking behaviour. Malar J. 2009;8(1):173. doi: 10.1186/1475-2875-8-173.

59. Awuah RB, Asante PY, Sakyi L, Biney AAE, Kushitor MK, Agyei F, et al. Factors associated with treatment-seeking for malaria in urban poor communities in Accra, Ghana. Malar J. 2018;17(1):168. doi: 10.1186/s12936-018-2311-8.

60. Simsek Z, Kurcer MA. Malaria: knowledge and behaviour in an endemic rural area of Turkey. Public Health. 2005;119(3):202-208. doi: https://doi.org/10.1016/j.puhe.2004.03.011.

61. Khairy S, Al-Surimi K, Ali A, Shubily HM, Al Walaan N, Househ M, et al. Knowledge, attitude and practice about malaria in south-western Saudi Arabia: A household-based cross-sectional survey. J Infect Public Health. 2017;10(5):499-506. doi: https://doi.org/10.1016/j.jiph.2016.09.021.

62. Nzooma SM, Tembo-Mwase E, Gebreslasie M, Samson M. Knowledge, attitudes and practices in the control and prevention of malaria in four endemic provinces of Zambia. S Afr J Infect Dis. 2017;32(1). doi: http://dx.doi.org/10.1080/23120053.2016.1205330. PubMed PMID: 2200892527.

63. Mbanefo A, Kumar N. Evaluation of Malaria Diagnostic Methods as a Key for Successful Control and Elimination Programs. 2020;5(2):102. PubMed PMID: doi:10.3390/tropicalmed5020102.

64. Van der Heide I, Wang J, Droomers M, Spreeuwenberg P, Rademakers J, Uiters E. The Relationship Between Health, Education, and Health Literacy: Results From the Dutch Adult Literacy and Life Skills Survey. J Health Commun. 2013;18(sup1):172–184. doi: 10.1080/10810730.2013.825668.

65. Berkman ND, Sheridan SL, Donahue KE, Halpern DJ, Crotty K. Low health literacy and health outcomes: an updated systematic review. Ann Intern Med. 2011;155(2):97–107.

66. Adhikari B, Phommasone K, Pongvongsa T, Koummarasy P, Soundala X, Henriques G, et al. Treatment-seeking behaviour for febrile illnesses and its implications for malaria control and elimination in Savannakhet Province, Lao PDR (Laos): a mixed method study. BMC Health Serv Res. 2019;19(1):252. doi: 10.1186/s12913-019-4070-9.

67. Regassa H, Taffere GR, Gebregergs GB. Delay in malaria diagnosis and treatment and its determinants among rural communities of the Oromia special zone, Ethiopia: facility-based cross-sectional study. Journal of Public Health. 2018;26(3):339–344. doi: 10.1007/s10389-017-0863-7.

68. Workineh B, Mekonnen FA. Early treatment-seeking behaviour for malaria in febrile patients in northwest Ethiopia. Malar J. 2018;17(1):406. doi: 10.1186/s12936-018-2556-2.

69. Romay-Barja M, Cano J, Ncogo P, Nseng G, Santana-Morales MA, Valladares B, et al. Determinants of delay in malaria care-seeking behaviour for children 15 years and under in Bata district, Equatorial Guinea. Malar J. 2016;15(1):187. doi: 10.1186/s12936-016-1239-0.

70. Yendris KS, Kuntoro H. The distance, alternative modes of transportation and the decision-making time to health facilities for the poor mother in a remote area: a case study in Central Sumba District, East Nusa Tenggara Province, Indonesia. 50TH APACPH Conference; Kota Kinabalu, Malaysia. 2018.

71. World Health Organization. Global technical strategy for malaria 2016–2030. Geneva: World Health Organization; 2015.

72. Guntur RD, Maria L. An Analysis of Factors that Affect Out of School Junior High Aged Children Using Logistic Regression Method. Asia-Pacific Collaborative Education Journal. 2015;11(2):23–36.

