## Supplemental Table for "Malaria treatment-seeking behaviour and its associated factors: A cross-sectional study in rural East Nusa Tenggara Province, Indonesia"

| RID | G | AG | D | Edu | Eth | Job | SES | FZ | HF | DHF | Inc1 | Inc2 | Q1 | Q2 | Q3 | Q4 | O1 | O2 | O3 |
|---|---|---|---|---|---|---|---|---|---|---|---|---|---|---|---|---|---|---|---|
| 10111 | 1 | 4 | 2 | 3 | 3 | 2 | 3 | 2 | 1 | 2 | 1 | 1 | 1 | 1 | 1 | 1 | 0 | 0 | 0 |
| 10112 | 1 | 2 | 2 | 5 | 3 | 5 | 3 | 1 | 1 | 2 | 0 | 0 | 1 | 1 | 1 | 1 | 0 | 0 | 0 |
| 10113 | 1 | 3 | 2 | 2 | 3 | 2 | 2 | 1 | 1 | 2 | 0 | 0 | 1 | 1 | 1 | 1 | 0 | 0 | 0 |
| 10114 | 1 | 1 | 2 | 5 | 3 | 5 | 2 | 1 | 1 | 2 | 0 | 0 | 1 | 1 | 1 | 1 | 0 | 0 | 0 |
| 10115 | 1 | 4 | 2 | 3 | 3 | 5 | 2 | 1 | 1 | 2 | 1 | 1 | 1 | 1 | 1 | 1 | 0 | 0 | 0 |
| 10116 | 1 | 3 | 2 | 3 | 3 | 2 | 2 | 1 | 1 | 2 | 0 | 0 | 1 | 1 | 1 | 1 | 0 | 0 | 0 |
| 10117 | 1 | 1 | 2 | 5 | 3 | 2 | 2 | 1 | 1 | 2 | 0 | 0 | 1 | 1 | 1 | 1 | 0 | 0 | 0 |
| 10118 | 1 | 3 | 2 | 4 | 3 | 5 | 2 | 1 | 1 | 2 | 1 | 0 | 1 | 1 | 1 | 1 | 0 | 0 | 0 |
| 10119 | 1 | 3 | 2 | 3 | 3 | 2 | 2 | 1 | 1 | 2 | 0 | 0 | 1 | 1 | 1 | 1 | 0 | 0 | 0 |
| 101110 | 1 | 4 | 2 | 2 | 3 | 2 | 2 | 1 | 1 | 2 | 0 | 0 | 1 | 1 | 1 | 1 | 0 | 0 | 0 |
| 101111 | 1 | 3 | 2 | 2 | 3 | 2 | 2 | 2 | 1 | 2 | 0 | 0 | 1 | 1 | 1 | 1 | 0 | 0 | 0 |
| 101112 | 1 | 1 | 2 | 5 | 3 | 5 | 2 | 1 | 1 | 2 | 1 | 0 | 1 | 1 | 1 | 1 | 0 | 0 | 0 |
| 101113 | 1 | 4 | 2 | 4 | 3 | 5 | 2 | 1 | 2 | 2 | 1 | 1 | 1 | 1 | 1 | 1 | 0 | 0 | 0 |
| 101114 | 1 | 5 | 2 | 5 | 3 | 5 | 3 | 1 | 1 | 2 | 1 | 1 | 1 | 1 | 1 | 1 | 0 | 0 | 0 |
| 101115 | 0 | 5 | 2 | 1 | 3 | 2 | 1 | 1 | 1 | 2 | 0 | 0 | 0 | 99 | 99 | 99 | 99 | 99 | 99 |
| 101116 | 0 | 2 | 2 | 3 | 3 | 1 | 2 | 2 | 1 | 1 | 0 | 0 | 1 | 1 | 1 | 1 | 0 | 0 | 0 |
| 101117 | 1 | 3 | 2 | 5 | 3 | 5 | 3 | 1 | 1 | 1 | 1 | 1 | 1 | 1 | 2 | 1 | 1 | 0 | 1 |
| 101118 | 1 | 4 | 2 | 3 | 3 | 2 | 2 | 1 | 1 | 1 | 0 | 0 | 1 | 1 | 1 | 1 | 0 | 0 | 0 |
| 101119 | 1 | 2 | 2 | 3 | 3 | 2 | 2 | 2 | 1 | 1 | 0 | 0 | 1 | 1 | 1 | 1 | 0 | 0 | 0 |
| 101120 | 0 | 4 | 2 | 2 | 3 | 1 | 1 | 1 | 1 | 1 | 0 | 0 | 1 | 1 | 1 | 1 | 0 | 0 | 0 |
| 101121 | 1 | 3 | 2 | 3 | 3 | 2 | 1 | 1 | 1 | 1 | 0 | 0 | 1 | 1 | 1 | 1 | 0 | 0 | 0 |
| 101122 | 0 | 4 | 2 | 1 | 3 | 1 | 1 | 1 | 1 | 1 | 0 | 0 | 1 | 1 | 1 | 1 | 0 | 0 | 0 |
| 101123 | 0 | 4 | 2 | 3 | 3 | 1 | 2 | 1 | 1 | 1 | 0 | 0 | 1 | 1 | 1 | 1 | 0 | 0 | 0 |
| 101124 | 0 | 4 | 2 | 1 | 3 | 1 | 2 | 1 | 1 | 1 | 0 | 0 | 1 | 1 | 1 | 1 | 0 | 0 | 0 |
| 101125 | 1 | 5 | 2 | 1 | 3 | 2 | 2 | 1 | 1 | 1 | 0 | 0 | 1 | 1 | 1 | 1 | 0 | 0 | 0 |
| 101126 | 0 | 3 | 2 | 3 | 3 | 1 | 2 | 1 | 1 | 1 | 0 | 0 | 1 | 1 | 1 | 1 | 0 | 0 | 0 |
| 101127 | 1 | 3 | 2 | 3 | 3 | 2 | 2 | 1 | 1 | 1 | 0 | 0 | 1 | 1 | 1 | 1 | 0 | 0 | 0 |
| 101128 | 1 | 4 | 2 | 3 | 3 | 2 | 2 | 1 | 1 | 1 | 0 | 0 | 1 | 1 | 1 | 1 | 0 | 0 | 0 |
| 101129 | 0 | 2 | 2 | 4 | 3 | 1 | 1 | 1 | 1 | 1 | 0 | 0 | 1 | 1 | 1 | 1 | 0 | 0 | 0 |
| 101130 | 1 | 4 | 2 | 3 | 3 | 2 | 2 | 1 | 1 | 1 | 0 | 0 | 1 | 1 | 1 | 1 | 0 | 0 | 0 |
| 10121 | 0 | 1 | 2 | 3 | 3 | 1 | 2 | 1 | 3 | 3 | 0 | 0 | 1 | 1 | 1 | 1 | 0 | 0 | 0 |
| 10122 | 1 | 2 | 2 | 4 | 3 | 2 | 2 | 2 | 3 | 3 | 0 | 0 | 1 | 1 | 1 | 1 | 0 | 0 | 0 |
| 10123 | 0 | 2 | 2 | 2 | 3 | 1 | 2 | 2 | 3 | 3 | 0 | 0 | 1 | 1 | 1 | 1 | 0 | 0 | 0 |
| 10124 | 0 | 1 | 2 | 4 | 3 | 1 | 2 | 1 | 3 | 3 | 0 | 0 | 1 | 1 | 1 | 1 | 0 | 0 | 0 |
| 10125 | 0 | 1 | 2 | 4 | 3 | 1 | 2 | 1 | 3 | 3 | 0 | 0 | 1 | 1 | 1 | 1 | 0 | 0 | 0 |
| 10126 | 0 | 5 | 2 | 1 | 3 | 2 | 1 | 1 | 3 | 3 | 0 | 0 | 0 | 99 | 99 | 99 | 99 | 99 | 99 |
| 10127 | 1 | 5 | 2 | 1 | 3 | 2 | 1 | 1 | 3 | 3 | 0 | 0 | 1 | 1 | 3 | 1 | 1 | 0 | 1 |
| 10128 | 0 | 3 | 2 | 2 | 3 | 1 | 2 | 1 | 3 | 2 | 0 | 0 | 1 | 1 | 1 | 1 | 0 | 0 | 0 |
| 10129 | 1 | 3 | 2 | 1 | 3 | 2 | 2 | 1 | 3 | 2 | 0 | 0 | 1 | 1 | 3 | 1 | 1 | 0 | 1 |
| 101210 | 0 | 2 | 2 | 1 | 3 | 1 | 2 | 2 | 3 | 2 | 0 | 0 | 1 | 1 | 1 | 1 | 0 | 0 | 0 |
| 101211 | 0 | 4 | 2 | 1 | 3 | 1 | 2 | 1 | 3 | 2 | 0 | 0 | 1 | 1 | 1 | 1 | 0 | 0 | 0 |
| 101212 | 0 | 2 | 2 | 4 | 3 | 1 | 3 | 1 | 3 | 2 | 0 | 0 | 1 | 1 | 3 | 1 | 1 | 0 | 1 |
| 101213 | 1 | 5 | 2 | 2 | 3 | 2 | 2 | 1 | 3 | 2 | 0 | 0 | 1 | 1 | 3 | 1 | 1 | 0 | 1 |
| 101214 | 1 | 4 | 2 | 1 | 3 | 2 | 2 | 1 | 3 | 2 | 0 | 0 | 1 | 1 | 1 | 1 | 0 | 0 | 0 |
| 101215 | 0 | 1 | 2 | 1 | 3 | 1 | 2 | 1 | 3 | 2 | 0 | 0 | 0 | 99 | 99 | 99 | 99 | 99 | 99 |
| 101216 | 0 | 5 | 2 | 1 | 3 | 1 | 1 | 1 | 3 | 2 | 0 | 0 | 1 | 1 | 1 | 1 | 0 | 0 | 0 |
| 101217 | 1 | 2 | 2 | 2 | 3 | 5 | 2 | 1 | 3 | 2 | 0 | 0 | 1 | 1 | 1 | 1 | 0 | 0 | 0 |
| 101218 | 0 | 1 | 2 | 2 | 3 | 1 | 2 | 1 | 3 | 2 | 0 | 0 | 1 | 1 | 1 | 1 | 0 | 0 | 0 |
| 101219 | 1 | 2 | 2 | 3 | 3 | 4 | 2 | 1 | 3 | 3 | 0 | 0 | 1 | 1 | 1 | 1 | 0 | 0 | 0 |
| 101220 | 0 | 2 | 2 | 2 | 3 | 1 | 2 | 1 | 3 | 3 | 0 | 0 | 1 | 1 | 1 | 1 | 0 | 0 | 0 |
| 101221 | 1 | 2 | 2 | 3 | 3 | 4 | 2 | 1 | 3 | 3 | 0 | 0 | 1 | 1 | 1 | 1 | 0 | 0 | 0 |
| 101222 | 1 | 2 | 2 | 4 | 3 | 4 | 3 | 1 | 3 | 3 | 1 | 0 | 1 | 1 | 1 | 1 | 0 | 0 | 0 |
| 101223 | 1 | 4 | 2 | 1 | 3 | 4 | 2 | 1 | 3 | 3 | 0 | 0 | 1 | 1 | 1 | 1 | 0 | 0 | 0 |
| 101224 | 0 | 3 | 2 | 2 | 3 | 1 | 2 | 1 | 3 | 2 | 0 | 0 | 1 | 1 | 1 | 1 | 0 | 0 | 0 |
| 101225 | 1 | 1 | 2 | 3 | 3 | 4 | 2 | 1 | 3 | 3 | 0 | 0 | 1 | 1 | 1 | 1 | 0 | 0 | 0 |

| | | | | | | | | | | | | | | | | | | | |
|---|---|---|---|---|---|---|---|---|---|---|---|---|---|---|---|---|---|---|---|
| 101226 | 0 | 3 | 2 | 2 | 2 | 1 | 1 | 1 | 4 | 1 | 0 | 0 | 1 | 1 | 3 | 1 | 1 | 0 | 1 |
| 101227 | 1 | 2 | 2 | 3 | 2 | 4 | 2 | 1 | 3 | 1 | 0 | 0 | 1 | 1 | 1 | 1 | 0 | 0 | 0 |
| 101228 | 0 | 3 | 2 | 3 | 3 | 1 | 2 | 1 | 3 | 1 | 0 | 0 | 1 | 1 | 1 | 1 | 0 | 0 | 0 |
| 101229 | 1 | 2 | 2 | 4 | 3 | 1 | 2 | 1 | 3 | 1 | 1 | 0 | 1 | 1 | 1 | 1 | 0 | 0 | 0 |
| 101230 | 1 | 3 | 2 | 3 | 3 | 2 | 3 | 1 | 3 | 1 | 0 | 0 | 1 | 1 | 1 | 1 | 0 | 0 | 0 |
| 101231 | 1 | 5 | 2 | 1 | 3 | 2 | 1 | 1 | 3 | 1 | 0 | 0 | 0 | 99 | 99 | 99 | 99 | 99 | 99 |
| 101232 | 1 | 5 | 2 | 1 | 3 | 2 | 1 | 1 | 3 | 2 | 0 | 0 | 1 | 1 | 1 | 1 | 0 | 0 | 0 |
| 101233 | 1 | 2 | 2 | 1 | 3 | 2 | 1 | 1 | 3 | 1 | 0 | 0 | 1 | 1 | 1 | 1 | 0 | 0 | 0 |
| 101234 | 1 | 3 | 2 | 2 | 3 | 2 | 1 | 2 | 3 | 3 | 0 | 0 | 1 | 1 | 1 | 1 | 0 | 0 | 0 |
| 101235 | 1 | 5 | 2 | 1 | 3 | 2 | 1 | 1 | 3 | 3 | 0 | 0 | 1 | 1 | 1 | 1 | 0 | 0 | 0 |
| 10131 | 0 | 3 | 2 | 3 | 3 | 1 | 2 | 1 | 4 | 1 | 0 | 0 | 1 | 1 | 1 | 1 | 0 | 0 | 0 |
| 10132 | 1 | 1 | 2 | 4 | 3 | 5 | 2 | 1 | 4 | 1 | 0 | 0 | 1 | 1 | 1 | 1 | 0 | 0 | 0 |
| 10133 | 1 | 3 | 2 | 4 | 3 | 5 | 2 | 1 | 4 | 1 | 0 | 0 | 1 | 1 | 1 | 1 | 0 | 0 | 0 |
| 10134 | 0 | 2 | 2 | 4 | 3 | 1 | 2 | 1 | 4 | 1 | 1 | 0 | 1 | 1 | 1 | 1 | 0 | 0 | 0 |
| 10135 | 0 | 1 | 2 | 4 | 3 | 1 | 2 | 1 | 4 | 1 | 0 | 0 | 1 | 1 | 1 | 1 | 0 | 0 | 0 |
| 10136 | 0 | 3 | 2 | 4 | 3 | 1 | 2 | 1 | 4 | 1 | 0 | 0 | 1 | 1 | 1 | 1 | 0 | 0 | 0 |
| 10137 | 1 | 2 | 2 | 4 | 3 | 5 | 3 | 2 | 4 | 2 | 0 | 0 | 1 | 1 | 1 | 1 | 0 | 0 | 0 |
| 10138 | 0 | 2 | 2 | 4 | 3 | 1 | 2 | 1 | 4 | 2 | 1 | 0 | 1 | 1 | 1 | 1 | 0 | 0 | 0 |
| 10139 | 0 | 2 | 2 | 3 | 3 | 1 | 2 | 1 | 4 | 2 | 1 | 0 | 1 | 1 | 1 | 1 | 0 | 0 | 0 |
| 101310 | 1 | 2 | 2 | 4 | 3 | 5 | 2 | 1 | 4 | 2 | 0 | 0 | 1 | 1 | 1 | 1 | 0 | 0 | 0 |
| 101311 | 1 | 1 | 2 | 4 | 3 | 5 | 2 | 1 | 4 | 2 | 0 | 0 | 1 | 1 | 1 | 1 | 0 | 0 | 0 |
| 101312 | 0 | 2 | 2 | 4 | 3 | 1 | 2 | 1 | 4 | 2 | 0 | 0 | 1 | 1 | 1 | 1 | 0 | 0 | 0 |
| 101313 | 0 | 3 | 2 | 1 | 3 | 1 | 1 | 1 | 4 | 2 | 0 | 0 | 1 | 1 | 1 | 1 | 0 | 0 | 0 |
| 101314 | 1 | 2 | 2 | 4 | 3 | 2 | 2 | 1 | 4 | 2 | 0 | 0 | 1 | 1 | 1 | 1 | 0 | 0 | 0 |
| 101315 | 1 | 1 | 2 | 3 | 3 | 2 | 1 | 1 | 4 | 2 | 0 | 0 | 1 | 1 | 1 | 1 | 0 | 0 | 0 |
| 101316 | 0 | 3 | 2 | 3 | 3 | 1 | 2 | 1 | 4 | 2 | 0 | 0 | 1 | 1 | 1 | 1 | 0 | 0 | 0 |
| 101317 | 0 | 2 | 2 | 3 | 3 | 1 | 2 | 1 | 4 | 2 | 0 | 0 | 1 | 1 | 1 | 1 | 0 | 0 | 0 |
| 101318 | 0 | 4 | 2 | 4 | 3 | 1 | 2 | 2 | 4 | 2 | 0 | 0 | 1 | 1 | 1 | 1 | 0 | 0 | 0 |
| 101319 | 1 | 3 | 2 | 3 | 3 | 3 | 3 | 1 | 4 | 2 | 0 | 0 | 1 | 1 | 1 | 1 | 0 | 0 | 0 |
| 101320 | 1 | 5 | 2 | 2 | 3 | 2 | 2 | 1 | 4 | 2 | 0 | 0 | 1 | 1 | 1 | 1 | 0 | 0 | 0 |
| 101321 | 0 | 4 | 2 | 2 | 3 | 1 | 2 | 1 | 4 | 2 | 0 | 0 | 1 | 1 | 1 | 1 | 0 | 0 | 0 |
| 101322 | 1 | 2 | 2 | 3 | 3 | 2 | 2 | 1 | 4 | 2 | 0 | 0 | 1 | 1 | 1 | 1 | 0 | 0 | 0 |
| 101323 | 0 | 2 | 2 | 1 | 3 | 2 | 2 | 2 | 4 | 2 | 0 | 0 | 1 | 1 | 1 | 1 | 0 | 0 | 0 |
| 101324 | 1 | 4 | 2 | 2 | 3 | 2 | 2 | 1 | 4 | 2 | 0 | 0 | 1 | 1 | 1 | 1 | 0 | 0 | 0 |
| 101325 | 1 | 3 | 2 | 3 | 3 | 1 | 2 | 1 | 4 | 2 | 0 | 0 | 1 | 1 | 1 | 1 | 0 | 0 | 0 |
| 101326 | 0 | 1 | 2 | 1 | 3 | 1 | 2 | 1 | 4 | 2 | 0 | 0 | 1 | 1 | 1 | 1 | 0 | 0 | 0 |
| 101327 | 1 | 1 | 2 | 3 | 3 | 2 | 2 | 1 | 4 | 2 | 0 | 0 | 1 | 1 | 1 | 1 | 0 | 0 | 0 |
| 101328 | 1 | 5 | 2 | 1 | 3 | 2 | 2 | 1 | 4 | 2 | 0 | 0 | 1 | 1 | 1 | 1 | 0 | 0 | 0 |
| 101329 | 0 | 1 | 2 | 4 | 3 | 1 | 2 | 1 | 4 | 2 | 0 | 0 | 1 | 1 | 1 | 1 | 0 | 0 | 0 |
| 101330 | 1 | 5 | 2 | 3 | 3 | 2 | 2 | 1 | 4 | 2 | 0 | 0 | 1 | 1 | 1 | 1 | 0 | 0 | 0 |
| 10141 | 1 | 2 | 2 | 5 | 3 | 5 | 3 | 1 | 4 | 1 | 0 | 0 | 1 | 1 | 1 | 1 | 0 | 0 | 0 |
| 10142 | 0 | 2 | 2 | 4 | 3 | 1 | 2 | 1 | 4 | 1 | 0 | 0 | 1 | 1 | 3 | 1 | 1 | 0 | 1 |
| 10143 | 1 | 5 | 2 | 2 | 3 | 5 | 2 | 1 | 4 | 1 | 1 | 1 | 1 | 1 | 3 | 2 | 1 | 0 | 1 |
| 10144 | 0 | 1 | 2 | 4 | 3 | 5 | 2 | 1 | 4 | 1 | 0 | 0 | 1 | 1 | 3 | 1 | 1 | 0 | 1 |
| 10145 | 1 | 4 | 2 | 1 | 3 | 2 | 1 | 2 | 4 | 1 | 0 | 0 | 0 | 99 | 99 | 99 | 99 | 99 | 99 |
| 10146 | 0 | 5 | 2 | 2 | 2 | 1 | 1 | 2 | 4 | 1 | 0 | 0 | 0 | 99 | 99 | 99 | 99 | 99 | 99 |
| 10147 | 1 | 2 | 2 | 4 | 3 | 5 | 2 | 2 | 4 | 1 | 0 | 0 | 1 | 1 | 2 | 1 | 1 | 0 | 1 |
| 10148 | 1 | 4 | 2 | 1 | 3 | 2 | 2 | 2 | 4 | 1 | 0 | 0 | 1 | 1 | 3 | 6 | 1 | 1 | 1 |
| 10149 | 0 | 3 | 2 | 2 | 3 | 1 | 3 | 2 | 4 | 2 | 0 | 0 | 1 | 1 | 1 | 1 | 0 | 0 | 0 |
| 101410 | 0 | 3 | 2 | 2 | 2 | 1 | 2 | 2 | 4 | 2 | 0 | 0 | 1 | 0 | 5 | 5 | 1 | 1 | 1 |
| 101411 | 1 | 5 | 2 | 1 | 3 | 5 | 1 | 1 | 4 | 2 | 0 | 0 | 0 | 99 | 99 | 99 | 99 | 99 | 99 |
| 101412 | 1 | 3 | 2 | 2 | 3 | 2 | 2 | 1 | 4 | 2 | 0 | 0 | 1 | 1 | 5 | 5 | 1 | 1 | 1 |
| 101413 | 1 | 2 | 2 | 4 | 3 | 2 | 2 | 1 | 4 | 2 | 0 | 0 | 1 | 1 | 1 | 1 | 0 | 0 | 0 |
| 101414 | 1 | 3 | 2 | 3 | 3 | 2 | 1 | 1 | 4 | 2 | 0 | 0 | 0 | 99 | 99 | 99 | 99 | 99 | 99 |
| 101415 | 1 | 4 | 2 | 1 | 3 | 2 | 1 | 1 | 4 | 2 | 0 | 0 | 0 | 99 | 99 | 99 | 99 | 99 | 99 |
| 101416 | 0 | 4 | 2 | 1 | 3 | 1 | 1 | 1 | 4 | 2 | 0 | 0 | 0 | 99 | 99 | 99 | 99 | 99 | 99 |

| | | | | | | | | | | | | | | | | | | | |
|---|---|---|---|---|---|---|---|---|---|---|---|---|---|---|---|---|---|---|---|
| 101417 | 1 | 3 | 2 | 2 | 3 | 2 | 2 | 1 | 4 | 2 | 0 | 0 | 1 | 1 | 2 | 1 | 1 | 0 | 1 |
| 101418 | 1 | 4 | 2 | 2 | 3 | 2 | 1 | 1 | 4 | 2 | 0 | 0 | 1 | 1 | 3 | 1 | 1 | 0 | 1 |
| 101419 | 1 | 3 | 2 | 2 | 3 | 2 | 2 | 1 | 4 | 2 | 0 | 0 | 1 | 1 | 2 | 2 | 1 | 0 | 1 |
| 101420 | 0 | 2 | 2 | 3 | 3 | 1 | 2 | 2 | 4 | 2 | 0 | 0 | 1 | 1 | 3 | 1 | 1 | 0 | 1 |
| 101421 | 1 | 4 | 2 | 2 | 3 | 2 | 2 | 1 | 4 | 2 | 0 | 0 | 0 | 99 | 99 | 99 | 99 | 99 | 99 |
| 101422 | 0 | 3 | 2 | 1 | 3 | 1 | 1 | 1 | 4 | 2 | 0 | 0 | 0 | 99 | 99 | 99 | 99 | 99 | 99 |
| 101423 | 1 | 4 | 2 | 2 | 3 | 2 | 2 | 1 | 4 | 2 | 0 | 0 | 1 | 1 | 2 | 1 | 1 | 0 | 1 |
| 101424 | 0 | 1 | 2 | 4 | 2 | 1 | 2 | 1 | 4 | 1 | 0 | 0 | 1 | 1 | 2 | 1 | 1 | 0 | 1 |
| 101425 | 1 | 1 | 2 | 4 | 3 | 4 | 2 | 1 | 4 | 1 | 0 | 0 | 1 | 1 | 3 | 1 | 1 | 0 | 1 |
| 101426 | 0 | 1 | 2 | 3 | 3 | 1 | 1 | 1 | 4 | 2 | 0 | 0 | 1 | 1 | 1 | 1 | 0 | 0 | 0 |
| 101427 | 0 | 3 | 2 | 3 | 3 | 1 | 2 | 1 | 4 | 2 | 0 | 0 | 1 | 1 | 2 | 1 | 1 | 0 | 1 |
| 101428 | 0 | 4 | 2 | 1 | 3 | 1 | 2 | 2 | 4 | 2 | 0 | 0 | 1 | 1 | 2 | 1 | 1 | 0 | 1 |
| 101429 | 1 | 2 | 2 | 3 | 3 | 2 | 2 | 1 | 4 | 2 | 0 | 0 | 1 | 1 | 3 | 1 | 1 | 0 | 1 |
| 101430 | 0 | 3 | 2 | 2 | 3 | 1 | 2 | 1 | 4 | 2 | 0 | 0 | 1 | 1 | 2 | 1 | 1 | 0 | 1 |
| 101431 | 1 | 4 | 2 | 2 | 3 | 2 | 2 | 2 | 4 | 2 | 0 | 0 | 1 | 1 | 2 | 1 | 1 | 0 | 1 |
| 101432 | 1 | 4 | 2 | 1 | 3 | 1 | 2 | 1 | 4 | 2 | 0 | 0 | 1 | 1 | 3 | 1 | 1 | 0 | 1 |
| 101433 | 0 | 1 | 2 | 3 | 3 | 1 | 2 | 1 | 4 | 2 | 0 | 0 | 1 | 1 | 2 | 1 | 1 | 0 | 1 |
| 101434 | 1 | 2 | 2 | 3 | 4 | 2 | 1 | 1 | 4 | 2 | 0 | 0 | 1 | 1 | 1 | 1 | 0 | 0 | 0 |
| 101435 | 1 | 2 | 2 | 3 | 3 | 4 | 2 | 1 | 4 | 2 | 0 | 0 | 1 | 1 | 2 | 1 | 1 | 0 | 1 |
| 10151 | 0 | 2 | 2 | 4 | 3 | 1 | 2 | 2 | 4 | 2 | 0 | 0 | 1 | 1 | 2 | 1 | 1 | 0 | 1 |
| 10152 | 1 | 4 | 2 | 4 | 3 | 4 | 2 | 1 | 4 | 2 | 0 | 0 | 1 | 1 | 3 | 2 | 1 | 0 | 1 |
| 10153 | 0 | 2 | 2 | 3 | 3 | 1 | 2 | 1 | 4 | 2 | 0 | 0 | 1 | 1 | 2 | 1 | 1 | 0 | 1 |
| 10154 | 0 | 1 | 2 | 4 | 3 | 5 | 2 | 2 | 4 | 2 | 1 | 0 | 1 | 1 | 2 | 2 | 1 | 0 | 1 |
| 10155 | 1 | 3 | 2 | 3 | 3 | 4 | 2 | 1 | 4 | 2 | 0 | 0 | 1 | 1 | 1 | 1 | 0 | 0 | 0 |
| 10156 | 1 | 3 | 2 | 3 | 3 | 5 | 2 | 1 | 4 | 2 | 0 | 0 | 1 | 1 | 2 | 1 | 1 | 0 | 1 |
| 10157 | 0 | 1 | 2 | 4 | 3 | 1 | 2 | 1 | 1 | 1 | 0 | 0 | 1 | 1 | 2 | 1 | 1 | 0 | 1 |
| 10158 | 0 | 2 | 2 | 4 | 3 | 1 | 2 | 1 | 1 | 1 | 0 | 0 | 1 | 1 | 3 | 1 | 1 | 0 | 1 |
| 10159 | 1 | 5 | 2 | 4 | 3 | 5 | 3 | 1 | 4 | 3 | 1 | 1 | 1 | 1 | 2 | 1 | 1 | 0 | 1 |
| 101510 | 1 | 1 | 2 | 4 | 3 | 4 | 2 | 1 | 1 | 1 | 0 | 0 | 1 | 1 | 2 | 1 | 1 | 0 | 1 |
| 101511 | 0 | 4 | 2 | 4 | 3 | 5 | 3 | 2 | 1 | 1 | 1 | 1 | 1 | 1 | 3 | 1 | 1 | 0 | 1 |
| 101512 | 0 | 2 | 2 | 5 | 3 | 1 | 1 | 1 | 1 | 1 | 0 | 0 | 1 | 1 | 2 | 4 | 1 | 1 | 1 |
| 101513 | 1 | 3 | 2 | 4 | 3 | 4 | 2 | 2 | 1 | 2 | 1 | 0 | 1 | 1 | 2 | 1 | 1 | 0 | 1 |
| 101514 | 1 | 5 | 2 | 3 | 3 | 4 | 2 | 1 | 1 | 2 | 0 | 0 | 1 | 1 | 1 | 1 | 0 | 0 | 0 |
| 101515 | 0 | 1 | 2 | 4 | 3 | 1 | 2 | 1 | 1 | 1 | 0 | 0 | 1 | 1 | 2 | 1 | 1 | 0 | 1 |
| 101516 | 1 | 2 | 2 | 2 | 3 | 4 | 2 | 1 | 1 | 1 | 0 | 0 | 1 | 1 | 2 | 2 | 1 | 0 | 1 |
| 101517 | 0 | 2 | 2 | 3 | 2 | 1 | 2 | 2 | 1 | 1 | 0 | 0 | 1 | 1 | 3 | 1 | 1 | 0 | 1 |
| 101518 | 1 | 3 | 2 | 4 | 3 | 4 | 2 | 1 | 1 | 1 | 0 | 0 | 1 | 1 | 2 | 1 | 1 | 0 | 1 |
| 101519 | 0 | 2 | 2 | 4 | 3 | 1 | 2 | 2 | 1 | 1 | 0 | 0 | 1 | 1 | 2 | 1 | 1 | 0 | 1 |
| 101520 | 1 | 4 | 2 | 4 | 3 | 4 | 3 | 2 | 1 | 2 | 1 | 1 | 1 | 1 | 2 | 1 | 1 | 0 | 1 |
| 101521 | 0 | 5 | 2 | 2 | 3 | 1 | 2 | 1 | 1 | 2 | 0 | 0 | 1 | 1 | 3 | 1 | 1 | 0 | 1 |
| 101522 | 1 | 5 | 2 | 3 | 3 | 5 | 2 | 1 | 1 | 2 | 1 | 1 | 1 | 1 | 2 | 1 | 1 | 0 | 1 |
| 101523 | 1 | 4 | 2 | 4 | 3 | 5 | 2 | 1 | 1 | 1 | 1 | 1 | 1 | 1 | 2 | 2 | 1 | 0 | 1 |
| 101524 | 0 | 3 | 2 | 4 | 3 | 5 | 3 | 1 | 1 | 1 | 1 | 1 | 1 | 1 | 3 | 2 | 1 | 0 | 1 |
| 101525 | 1 | 2 | 2 | 4 | 3 | 5 | 2 | 2 | 1 | 1 | 1 | 1 | 1 | 1 | 2 | 1 | 1 | 0 | 1 |
| 101526 | 1 | 2 | 2 | 4 | 3 | 5 | 2 | 1 | 1 | 1 | 1 | 1 | 1 | 1 | 2 | 1 | 1 | 0 | 1 |
| 101527 | 1 | 3 | 2 | 4 | 3 | 5 | 2 | 1 | 1 | 1 | 0 | 0 | 1 | 1 | 2 | 1 | 1 | 0 | 1 |
| 101528 | 1 | 4 | 2 | 2 | 3 | 2 | 2 | 1 | 1 | 1 | 1 | 1 | 1 | 1 | 2 | 1 | 1 | 0 | 1 |
| 101529 | 0 | 3 | 2 | 3 | 3 | 1 | 2 | 1 | 1 | 1 | 0 | 0 | 1 | 1 | 3 | 1 | 1 | 0 | 1 |
| 101530 | 0 | 3 | 2 | 4 | 3 | 1 | 2 | 1 | 1 | 1 | 0 | 0 | 1 | 1 | 2 | 1 | 1 | 0 | 1 |
| 10161 | 0 | 1 | 2 | 4 | 3 | 1 | 2 | 1 | 1 | 1 | 0 | 0 | 1 | 1 | 2 | 1 | 1 | 0 | 1 |
| 10162 | 0 | 1 | 2 | 2 | 3 | 1 | 2 | 1 | 2 | 2 | 1 | 0 | 1 | 1 | 2 | 1 | 1 | 0 | 1 |
| 10163 | 0 | 2 | 2 | 4 | 3 | 1 | 3 | 1 | 1 | 2 | 0 | 0 | 1 | 1 | 3 | 1 | 1 | 0 | 1 |
| 10164 | 1 | 4 | 2 | 5 | 3 | 5 | 3 | 2 | 1 | 2 | 1 | 1 | 1 | 1 | 1 | 6 | 0 | 1 | 1 |
| 10165 | 1 | 2 | 2 | 2 | 3 | 2 | 2 | 1 | 1 | 2 | 0 | 0 | 1 | 1 | 2 | 1 | 1 | 0 | 1 |
| 10166 | 1 | 5 | 2 | 2 | 3 | 2 | 2 | 1 | 1 | 2 | 1 | 1 | 1 | 1 | 2 | 1 | 1 | 0 | 1 |
| 10167 | 1 | 4 | 2 | 2 | 3 | 4 | 2 | 2 | 1 | 1 | 0 | 0 | 1 | 1 | 2 | 1 | 1 | 0 | 1 |

| | | | | | | | | | | | | | | | | | | | |
|---|---|---|---|---|---|---|---|---|---|---|---|---|---|---|---|---|---|---|---|
| 10168 | 1 | 1 | 2 | 2 | 3 | 2 | 2 | 1 | 1 | 1 | 0 | 0 | 1 | 1 | 3 | 1 | 1 | 0 | 1 |
| 10169 | 0 | 3 | 2 | 2 | 3 | 1 | 2 | 1 | 1 | 1 | 0 | 0 | 1 | 1 | 3 | 2 | 1 | 0 | 1 |
| 101610 | 0 | 3 | 2 | 1 | 3 | 1 | 2 | 2 | 1 | 1 | 0 | 0 | 1 | 1 | 2 | 1 | 1 | 0 | 1 |
| 101611 | 0 | 1 | 2 | 2 | 3 | 1 | 2 | 2 | 1 | 1 | 0 | 0 | 1 | 1 | 2 | 6 | 1 | 1 | 1 |
| 101612 | 0 | 4 | 2 | 2 | 3 | 1 | 2 | 1 | 1 | 1 | 0 | 0 | 1 | 1 | 4 | 6 | 1 | 1 | 1 |
| 101613 | 1 | 3 | 2 | 2 | 3 | 4 | 2 | 1 | 1 | 1 | 0 | 0 | 1 | 1 | 2 | 1 | 1 | 0 | 1 |
| 101614 | 0 | 3 | 2 | 2 | 3 | 1 | 3 | 2 | 1 | 1 | 0 | 0 | 1 | 1 | 2 | 1 | 1 | 0 | 1 |
| 101615 | 0 | 4 | 2 | 2 | 3 | 1 | 2 | 1 | 1 | 1 | 0 | 0 | 1 | 1 | 3 | 1 | 1 | 0 | 1 |
| 101616 | 0 | 1 | 2 | 4 | 3 | 1 | 2 | 1 | 1 | 1 | 0 | 0 | 1 | 1 | 2 | 1 | 1 | 0 | 1 |
| 101617 | 0 | 3 | 2 | 1 | 3 | 1 | 2 | 2 | 1 | 1 | 0 | 0 | 1 | 1 | 2 | 1 | 1 | 0 | 1 |
| 101618 | 1 | 5 | 2 | 2 | 3 | 2 | 1 | 1 | 1 | 1 | 0 | 0 | 1 | 1 | 3 | 2 | 1 | 0 | 1 |
| 101619 | 1 | 4 | 2 | 2 | 3 | 4 | 2 | 2 | 1 | 1 | 0 | 0 | 1 | 1 | 3 | 6 | 1 | 1 | 1 |
| 101620 | 0 | 1 | 2 | 3 | 3 | 1 | 2 | 2 | 1 | 1 | 0 | 0 | 1 | 1 | 3 | 1 | 1 | 0 | 1 |
| 101621 | 1 | 4 | 2 | 2 | 3 | 4 | 2 | 1 | 1 | 1 | 0 | 0 | 1 | 1 | 2 | 1 | 1 | 0 | 1 |
| 101622 | 0 | 2 | 2 | 3 | 3 | 1 | 2 | 1 | 1 | 1 | 0 | 0 | 1 | 1 | 2 | 1 | 1 | 0 | 1 |
| 101623 | 1 | 1 | 2 | 2 | 3 | 4 | 2 | 1 | 1 | 1 | 0 | 0 | 1 | 1 | 3 | 1 | 1 | 0 | 1 |
| 101624 | 0 | 5 | 2 | 2 | 3 | 1 | 1 | 1 | 1 | 1 | 0 | 0 | 1 | 1 | 2 | 6 | 1 | 1 | 1 |
| 101625 | 1 | 2 | 2 | 3 | 1 | 2 | 2 | 1 | 1 | 1 | 0 | 0 | 1 | 1 | 3 | 1 | 1 | 0 | 1 |
| 101626 | 1 | 4 | 2 | 2 | 3 | 4 | 2 | 2 | 1 | 1 | 0 | 0 | 1 | 1 | 2 | 1 | 1 | 0 | 1 |
| 101627 | 1 | 3 | 2 | 2 | 3 | 4 | 2 | 1 | 1 | 1 | 0 | 0 | 1 | 1 | 2 | 1 | 1 | 0 | 1 |
| 101628 | 1 | 2 | 2 | 2 | 3 | 4 | 2 | 2 | 1 | 1 | 0 | 0 | 1 | 1 | 2 | 6 | 1 | 1 | 1 |
| 101629 | 1 | 3 | 2 | 3 | 3 | 4 | 2 | 1 | 1 | 1 | 0 | 0 | 1 | 1 | 2 | 1 | 1 | 0 | 1 |
| 101630 | 0 | 4 | 2 | 2 | 3 | 1 | 2 | 1 | 1 | 1 | 0 | 0 | 1 | 1 | 2 | 1 | 1 | 0 | 1 |
| 10271 | 0 | 3 | 2 | 1 | 3 | 1 | 2 | 2 | 1 | 1 | 0 | 0 | 1 | 1 | 3 | 4 | 1 | 1 | 1 |
| 10272 | 0 | 1 | 2 | 4 | 3 | 1 | 2 | 2 | 1 | 1 | 0 | 0 | 1 | 1 | 2 | 1 | 1 | 0 | 1 |
| 10273 | 0 | 2 | 2 | 2 | 3 | 1 | 2 | 2 | 1 | 3 | 0 | 0 | 1 | 1 | 3 | 1 | 1 | 0 | 1 |
| 10274 | 1 | 2 | 2 | 2 | 3 | 4 | 2 | 2 | 1 | 3 | 0 | 0 | 1 | 1 | 2 | 1 | 1 | 0 | 1 |
| 10275 | 0 | 4 | 2 | 2 | 3 | 1 | 1 | 1 | 1 | 3 | 0 | 0 | 1 | 1 | 1 | 5 | 0 | 1 | 1 |
| 10276 | 1 | 2 | 2 | 1 | 3 | 2 | 2 | 1 | 1 | 3 | 0 | 0 | 1 | 0 | 5 | 5 | 1 | 1 | 1 |
| 10277 | 0 | 1 | 2 | 3 | 3 | 1 | 2 | 2 | 1 | 3 | 0 | 0 | 1 | 1 | 3 | 6 | 1 | 1 | 1 |
| 10278 | 0 | 5 | 2 | 1 | 3 | 1 | 1 | 1 | 1 | 1 | 0 | 0 | 0 | 99 | 99 | 99 | 99 | 99 | 99 |
| 10279 | 1 | 4 | 2 | 2 | 3 | 2 | 2 | 2 | 1 | 3 | 0 | 0 | 1 | 1 | 1 | 1 | 0 | 0 | 0 |
| 102710 | 1 | 3 | 2 | 4 | 3 | 2 | 2 | 1 | 1 | 3 | 0 | 0 | 1 | 1 | 2 | 1 | 1 | 0 | 1 |
| 102711 | 1 | 4 | 2 | 1 | 3 | 2 | 2 | 1 | 1 | 3 | 0 | 0 | 1 | 1 | 4 | 4 | 1 | 1 | 1 |
| 102712 | 0 | 2 | 2 | 4 | 3 | 1 | 2 | 1 | 1 | 3 | 0 | 0 | 1 | 1 | 1 | 1 | 0 | 0 | 0 |
| 102713 | 0 | 4 | 2 | 2 | 3 | 1 | 2 | 1 | 1 | 3 | 0 | 0 | 0 | 99 | 99 | 99 | 99 | 99 | 99 |
| 102714 | 0 | 3 | 2 | 3 | 3 | 1 | 1 | 1 | 1 | 3 | 0 | 0 | 1 | 1 | 3 | 4 | 1 | 1 | 1 |
| 102715 | 1 | 5 | 2 | 1 | 3 | 2 | 1 | 1 | 1 | 3 | 0 | 0 | 0 | 99 | 99 | 99 | 99 | 99 | 99 |
| 102716 | 1 | 4 | 2 | 2 | 3 | 2 | 1 | 1 | 1 | 3 | 0 | 0 | 1 | 1 | 3 | 6 | 1 | 1 | 1 |
| 102717 | 0 | 4 | 2 | 2 | 3 | 1 | 1 | 1 | 1 | 3 | 0 | 0 | 0 | 99 | 99 | 99 | 99 | 99 | 99 |
| 102718 | 1 | 3 | 2 | 2 | 3 | 2 | 1 | 1 | 1 | 3 | 0 | 0 | 1 | 1 | 1 | 1 | 0 | 0 | 0 |
| 102719 | 0 | 4 | 2 | 1 | 3 | 2 | 1 | 1 | 1 | 3 | 0 | 0 | 1 | 1 | 2 | 5 | 1 | 1 | 1 |
| 102720 | 1 | 3 | 2 | 3 | 3 | 4 | 2 | 1 | 1 | 3 | 0 | 0 | 1 | 1 | 2 | 6 | 1 | 1 | 1 |
| 102721 | 1 | 4 | 2 | 2 | 3 | 2 | 2 | 1 | 1 | 3 | 0 | 0 | 1 | 1 | 1 | 1 | 0 | 0 | 0 |
| 102722 | 0 | 4 | 2 | 1 | 3 | 1 | 1 | 1 | 1 | 3 | 0 | 0 | 0 | 99 | 99 | 99 | 99 | 99 | 99 |
| 102723 | 0 | 2 | 2 | 2 | 3 | 1 | 1 | 1 | 1 | 3 | 0 | 0 | 1 | 1 | 3 | 5 | 1 | 1 | 1 |
| 102724 | 1 | 3 | 2 | 1 | 3 | 2 | 2 | 1 | 1 | 3 | 0 | 0 | 1 | 1 | 2 | 1 | 1 | 0 | 1 |
| 102725 | 0 | 2 | 2 | 2 | 3 | 1 | 1 | 2 | 1 | 3 | 0 | 0 | 1 | 1 | 4 | 1 | 1 | 0 | 1 |
| 102726 | 1 | 3 | 2 | 2 | 3 | 2 | 2 | 1 | 1 | 3 | 0 | 0 | 1 | 1 | 3 | 7 | 1 | 1 | 1 |
| 102727 | 1 | 2 | 2 | 2 | 3 | 2 | 2 | 1 | 1 | 2 | 0 | 0 | 1 | 1 | 1 | 1 | 0 | 0 | 0 |
| 102728 | 1 | 2 | 2 | 4 | 3 | 2 | 2 | 1 | 1 | 2 | 0 | 0 | 1 | 1 | 1 | 1 | 0 | 0 | 0 |
| 102729 | 1 | 2 | 2 | 3 | 3 | 4 | 2 | 2 | 1 | 3 | 0 | 0 | 1 | 1 | 2 | 6 | 1 | 1 | 1 |
| 102730 | 1 | 3 | 2 | 2 | 3 | 4 | 2 | 1 | 1 | 3 | 0 | 0 | 1 | 1 | 2 | 6 | 1 | 1 | 1 |
| 10281 | 1 | 3 | 2 | 2 | 3 | 2 | 2 | 1 | 1 | 4 | 0 | 0 | 1 | 1 | 2 | 6 | 1 | 1 | 1 |
| 10282 | 1 | 4 | 2 | 2 | 3 | 2 | 1 | 1 | 1 | 1 | 0 | 0 | 0 | 99 | 99 | 99 | 99 | 99 | 99 |
| 10283 | 0 | 4 | 2 | 1 | 3 | 1 | 1 | 1 | 1 | 4 | 0 | 0 | 1 | 1 | 3 | 5 | 1 | 1 | 1 |

| | | | | | | | | | | | | | | | | | | | |
|---|---|---|---|---|---|---|---|---|---|---|---|---|---|---|---|---|---|---|---|
| 10284 | 1 | 3 | 2 | 1 | 3 | 1 | 2 | 1 | 1 | 4 | 0 | 0 | 1 | 1 | 2 | 5 | 1 | 1 | 1 |
| 10285 | 1 | 4 | 2 | 2 | 3 | 2 | 2 | 1 | 1 | 4 | 0 | 0 | 1 | 1 | 1 | 4 | 0 | 1 | 1 |
| 10286 | 0 | 3 | 2 | 2 | 3 | 1 | 1 | 1 | 1 | 4 | 0 | 0 | 1 | 1 | 2 | 4 | 1 | 1 | 1 |
| 10287 | 0 | 5 | 2 | 1 | 3 | 1 | 1 | 1 | 1 | 4 | 0 | 0 | 0 | 99 | 99 | 99 | 99 | 99 | 99 |
| 10288 | 1 | 5 | 2 | 1 | 3 | 2 | 1 | 1 | 1 | 4 | 0 | 0 | 0 | 99 | 99 | 99 | 99 | 99 | 99 |
| 10289 | 1 | 1 | 2 | 3 | 3 | 2 | 1 | 1 | 1 | 4 | 0 | 0 | 1 | 1 | 3 | 1 | 1 | 0 | 1 |
| 102810 | 0 | 3 | 2 | 2 | 3 | 1 | 1 | 1 | 1 | 4 | 0 | 0 | 1 | 1 | 2 | 1 | 1 | 0 | 1 |
| 102811 | 1 | 3 | 2 | 3 | 3 | 2 | 2 | 1 | 1 | 4 | 0 | 0 | 1 | 1 | 1 | 2 | 0 | 0 | 0 |
| 102812 | 1 | 3 | 2 | 3 | 3 | 2 | 2 | 1 | 1 | 4 | 0 | 0 | 1 | 1 | 1 | 1 | 0 | 0 | 0 |
| 102813 | 1 | 4 | 2 | 2 | 3 | 2 | 1 | 1 | 1 | 4 | 0 | 0 | 1 | 1 | 5 | 4 | 1 | 1 | 1 |
| 102814 | 0 | 3 | 2 | 2 | 3 | 1 | 1 | 1 | 1 | 4 | 0 | 0 | 0 | 99 | 99 | 99 | 99 | 99 | 99 |
| 102815 | 1 | 5 | 2 | 1 | 3 | 2 | 1 | 1 | 1 | 4 | 0 | 0 | 0 | 99 | 99 | 99 | 99 | 99 | 99 |
| 102816 | 1 | 3 | 2 | 2 | 3 | 2 | 1 | 1 | 1 | 4 | 0 | 0 | 1 | 1 | 1 | 1 | 0 | 0 | 0 |
| 102817 | 0 | 4 | 2 | 2 | 3 | 1 | 1 | 1 | 1 | 4 | 0 | 0 | 0 | 99 | 99 | 99 | 99 | 99 | 99 |
| 102818 | 0 | 4 | 2 | 2 | 3 | 1 | 1 | 1 | 1 | 4 | 0 | 0 | 1 | 1 | 2 | 1 | 1 | 0 | 1 |
| 102819 | 1 | 3 | 2 | 2 | 3 | 2 | 1 | 1 | 1 | 4 | 0 | 0 | 1 | 1 | 3 | 6 | 1 | 1 | 1 |
| 102820 | 0 | 2 | 2 | 3 | 3 | 1 | 1 | 1 | 1 | 4 | 0 | 0 | 1 | 1 | 3 | 1 | 1 | 0 | 1 |
| 102821 | 1 | 2 | 2 | 3 | 3 | 2 | 3 | 1 | 1 | 1 | 0 | 0 | 1 | 1 | 2 | 1 | 1 | 0 | 1 |
| 102822 | 0 | 3 | 2 | 2 | 3 | 1 | 2 | 2 | 1 | 1 | 0 | 0 | 1 | 1 | 2 | 5 | 1 | 1 | 1 |
| 102823 | 0 | 3 | 2 | 2 | 3 | 1 | 3 | 1 | 1 | 1 | 0 | 0 | 1 | 1 | 1 | 1 | 0 | 0 | 0 |
| 102824 | 1 | 4 | 2 | 1 | 3 | 2 | 2 | 1 | 1 | 1 | 0 | 0 | 0 | 99 | 99 | 99 | 99 | 99 | 99 |
| 102825 | 1 | 4 | 2 | 2 | 3 | 2 | 2 | 1 | 1 | 1 | 0 | 0 | 1 | 1 | 1 | 1 | 0 | 0 | 0 |
| 102826 | 0 | 4 | 2 | 2 | 3 | 1 | 2 | 1 | 1 | 1 | 0 | 0 | 1 | 1 | 1 | 1 | 0 | 0 | 0 |
| 102827 | 0 | 5 | 2 | 2 | 3 | 1 | 3 | 1 | 1 | 1 | 0 | 0 | 1 | 1 | 1 | 1 | 0 | 0 | 0 |
| 102828 | 0 | 2 | 2 | 2 | 3 | 1 | 3 | 1 | 1 | 1 | 0 | 0 | 1 | 1 | 1 | 1 | 0 | 0 | 0 |
| 102829 | 1 | 5 | 2 | 1 | 3 | 2 | 3 | 1 | 1 | 1 | 0 | 0 | 1 | 1 | 1 | 1 | 0 | 0 | 0 |
| 102830 | 0 | 1 | 2 | 3 | 3 | 1 | 2 | 1 | 1 | 1 | 0 | 0 | 1 | 1 | 1 | 1 | 0 | 0 | 0 |
| 10291 | 1 | 5 | 2 | 2 | 3 | 2 | 2 | 1 | 1 | 2 | 0 | 0 | 1 | 1 | 1 | 6 | 0 | 1 | 1 |
| 10292 | 0 | 4 | 2 | 2 | 3 | 1 | 2 | 1 | 1 | 2 | 0 | 0 | 0 | 99 | 99 | 99 | 99 | 99 | 99 |
| 10293 | 0 | 5 | 2 | 1 | 3 | 1 | 2 | 1 | 1 | 2 | 0 | 0 | 0 | 99 | 99 | 99 | 99 | 99 | 99 |
| 10294 | 1 | 1 | 2 | 3 | 3 | 2 | 2 | 1 | 1 | 2 | 0 | 0 | 1 | 1 | 1 | 1 | 0 | 0 | 0 |
| 10295 | 1 | 2 | 2 | 3 | 3 | 2 | 2 | 1 | 1 | 2 | 0 | 0 | 1 | 1 | 1 | 5 | 0 | 1 | 1 |
| 10296 | 1 | 5 | 2 | 2 | 3 | 2 | 3 | 2 | 1 | 2 | 0 | 0 | 1 | 1 | 3 | 1 | 1 | 0 | 1 |
| 10297 | 1 | 3 | 2 | 2 | 3 | 2 | 2 | 1 | 1 | 2 | 0 | 0 | 0 | 99 | 99 | 99 | 99 | 99 | 99 |
| 10298 | 0 | 1 | 2 | 4 | 3 | 1 | 3 | 1 | 1 | 2 | 0 | 0 | 1 | 1 | 2 | 1 | 1 | 0 | 1 |
| 10299 | 1 | 4 | 2 | 2 | 3 | 2 | 2 | 1 | 1 | 2 | 0 | 0 | 1 | 1 | 3 | 5 | 1 | 1 | 1 |
| 102910 | 0 | 3 | 2 | 2 | 3 | 1 | 2 | 2 | 1 | 2 | 0 | 0 | 1 | 1 | 1 | 1 | 0 | 0 | 0 |
| 102911 | 1 | 3 | 2 | 3 | 3 | 2 | 2 | 1 | 1 | 2 | 0 | 0 | 1 | 1 | 3 | 1 | 1 | 0 | 1 |
| 102912 | 0 | 1 | 2 | 3 | 3 | 1 | 2 | 1 | 1 | 2 | 0 | 0 | 1 | 1 | 2 | 5 | 1 | 1 | 1 |
| 102913 | 1 | 3 | 2 | 2 | 3 | 2 | 2 | 1 | 1 | 2 | 0 | 0 | 1 | 1 | 3 | 1 | 1 | 0 | 1 |
| 102914 | 0 | 4 | 2 | 2 | 3 | 1 | 2 | 1 | 1 | 2 | 0 | 0 | 1 | 1 | 1 | 1 | 0 | 0 | 0 |
| 102915 | 0 | 4 | 2 | 1 | 3 | 1 | 2 | 1 | 1 | 2 | 0 | 0 | 1 | 1 | 2 | 1 | 1 | 0 | 1 |
| 102916 | 1 | 4 | 2 | 2 | 3 | 2 | 2 | 1 | 1 | 2 | 0 | 0 | 0 | 99 | 99 | 99 | 99 | 99 | 99 |
| 102917 | 1 | 3 | 2 | 2 | 3 | 2 | 2 | 1 | 1 | 2 | 0 | 0 | 1 | 1 | 1 | 5 | 0 | 1 | 1 |
| 102918 | 0 | 4 | 2 | 2 | 3 | 1 | 2 | 1 | 1 | 2 | 0 | 0 | 0 | 99 | 99 | 99 | 99 | 99 | 99 |
| 102919 | 0 | 4 | 2 | 2 | 3 | 1 | 2 | 1 | 1 | 2 | 0 | 0 | 0 | 99 | 99 | 99 | 99 | 99 | 99 |
| 102920 | 0 | 1 | 2 | 3 | 3 | 1 | 2 | 1 | 1 | 2 | 0 | 0 | 1 | 1 | 3 | 1 | 1 | 0 | 1 |
| 102921 | 0 | 1 | 2 | 2 | 3 | 1 | 2 | 1 | 1 | 2 | 0 | 0 | 1 | 1 | 2 | 1 | 1 | 0 | 1 |
| 102922 | 1 | 3 | 2 | 2 | 3 | 2 | 2 | 1 | 1 | 2 | 0 | 0 | 1 | 1 | 3 | 5 | 1 | 1 | 1 |
| 102923 | 1 | 3 | 2 | 2 | 3 | 2 | 2 | 1 | 1 | 2 | 0 | 0 | 1 | 1 | 1 | 1 | 0 | 0 | 0 |
| 102924 | 1 | 2 | 2 | 3 | 3 | 2 | 2 | 1 | 1 | 2 | 0 | 0 | 0 | 99 | 99 | 99 | 99 | 99 | 99 |
| 102925 | 0 | 2 | 2 | 4 | 3 | 1 | 3 | 1 | 1 | 2 | 0 | 0 | 1 | 1 | 2 | 1 | 1 | 0 | 1 |
| 102926 | 1 | 3 | 2 | 2 | 3 | 2 | 2 | 1 | 1 | 2 | 0 | 0 | 1 | 1 | 3 | 1 | 1 | 0 | 1 |
| 102927 | 1 | 3 | 2 | 2 | 3 | 2 | 2 | 1 | 1 | 2 | 0 | 0 | 0 | 99 | 99 | 99 | 99 | 99 | 99 |
| 102928 | 0 | 4 | 2 | 3 | 3 | 1 | 2 | 1 | 1 | 2 | 0 | 0 | 1 | 1 | 2 | 5 | 1 | 1 | 1 |
| 102929 | 0 | 4 | 2 | 2 | 3 | 1 | 2 | 1 | 1 | 2 | 0 | 0 | 1 | 1 | 2 | 6 | 1 | 1 | 1 |

| | | | | | | | | | | | | | | | | | | | |
|---|---|---|---|---|---|---|---|---|---|---|---|---|---|---|---|---|---|---|---|
| 102930 | 1 | 4 | 2 | 2 | 3 | 2 | 2 | 1 | 1 | 2 | 0 | 0 | 1 | 1 | 2 | 5 | 1 | 1 | 1 |
| 102101 | 0 | 1 | 2 | 4 | 3 | 1 | 2 | 2 | 1 | 1 | 0 | 0 | 1 | 1 | 1 | 1 | 0 | 0 | 0 |
| 102102 | 0 | 2 | 2 | 3 | 3 | 1 | 2 | 1 | 1 | 1 | 0 | 0 | 1 | 1 | 3 | 1 | 1 | 0 | 1 |
| 102103 | 0 | 2 | 2 | 2 | 3 | 1 | 2 | 2 | 2 | 1 | 0 | 0 | 1 | 1 | 3 | 1 | 1 | 0 | 1 |
| 102104 | 0 | 5 | 2 | 1 | 3 | 2 | 2 | 2 | 1 | 1 | 0 | 0 | 1 | 1 | 3 | 1 | 1 | 0 | 1 |
| 102105 | 1 | 5 | 2 | 3 | 3 | 2 | 3 | 2 | 1 | 1 | 0 | 0 | 1 | 1 | 1 | 1 | 0 | 0 | 0 |
| 102106 | 1 | 4 | 2 | 1 | 3 | 2 | 2 | 2 | 4 | 2 | 0 | 0 | 1 | 1 | 3 | 1 | 1 | 0 | 1 |
| 102107 | 0 | 1 | 2 | 4 | 3 | 1 | 2 | 1 | 4 | 1 | 0 | 0 | 1 | 1 | 3 | 1 | 1 | 0 | 1 |
| 102108 | 0 | 2 | 2 | 2 | 3 | 1 | 2 | 2 | 1 | 1 | 0 | 0 | 1 | 1 | 3 | 1 | 1 | 0 | 1 |
| 102109 | 1 | 3 | 2 | 2 | 3 | 2 | 3 | 1 | 1 | 1 | 0 | 0 | 1 | 1 | 3 | 1 | 1 | 0 | 1 |
| 1021010 | 0 | 3 | 2 | 2 | 3 | 1 | 2 | 1 | 1 | 1 | 0 | 0 | 1 | 1 | 3 | 1 | 1 | 0 | 1 |
| 1021011 | 1 | 4 | 2 | 1 | 2 | 2 | 2 | 1 | 1 | 1 | 0 | 0 | 1 | 1 | 3 | 1 | 1 | 0 | 1 |
| 1021012 | 0 | 1 | 2 | 2 | 3 | 1 | 3 | 1 | 1 | 1 | 0 | 0 | 1 | 1 | 3 | 1 | 1 | 0 | 1 |
| 1021013 | 0 | 5 | 2 | 1 | 3 | 2 | 2 | 1 | 4 | 1 | 0 | 0 | 1 | 1 | 4 | 5 | 1 | 1 | 1 |
| 1021014 | 0 | 2 | 2 | 2 | 3 | 1 | 2 | 1 | 1 | 1 | 0 | 0 | 1 | 1 | 3 | 1 | 1 | 0 | 1 |
| 1021015 | 1 | 5 | 2 | 1 | 3 | 2 | 2 | 1 | 1 | 1 | 0 | 0 | 1 | 1 | 1 | 1 | 0 | 0 | 0 |
| 1021016 | 1 | 4 | 2 | 2 | 3 | 2 | 2 | 2 | 4 | 1 | 0 | 0 | 1 | 1 | 3 | 1 | 1 | 0 | 1 |
| 1021017 | 1 | 2 | 2 | 3 | 3 | 4 | 2 | 2 | 2 | 4 | 0 | 0 | 1 | 1 | 3 | 1 | 1 | 0 | 1 |
| 1021018 | 1 | 2 | 2 | 1 | 3 | 2 | 2 | 2 | 1 | 1 | 0 | 0 | 1 | 1 | 3 | 1 | 1 | 0 | 1 |
| 1021019 | 1 | 4 | 2 | 1 | 3 | 2 | 2 | 1 | 1 | 1 | 0 | 0 | 1 | 1 | 3 | 1 | 1 | 0 | 1 |
| 1021020 | 1 | 5 | 2 | 2 | 2 | 2 | 2 | 1 | 1 | 1 | 0 | 0 | 1 | 0 | 5 | 5 | 1 | 1 | 1 |
| 1021021 | 1 | 4 | 2 | 1 | 3 | 2 | 2 | 1 | 1 | 1 | 0 | 0 | 1 | 1 | 3 | 2 | 1 | 0 | 1 |
| 1021022 | 1 | 3 | 2 | 1 | 3 | 2 | 2 | 1 | 1 | 1 | 0 | 0 | 0 | 99 | 99 | 99 | 99 | 99 | 99 |
| 1021023 | 1 | 4 | 2 | 2 | 3 | 5 | 3 | 2 | 4 | 1 | 1 | 0 | 1 | 1 | 4 | 1 | 1 | 0 | 1 |
| 1021024 | 1 | 3 | 2 | 1 | 3 | 2 | 2 | 1 | 1 | 1 | 0 | 0 | 1 | 1 | 3 | 1 | 1 | 0 | 1 |
| 1021025 | 1 | 4 | 2 | 2 | 3 | 5 | 2 | 2 | 4 | 2 | 0 | 0 | 1 | 1 | 3 | 1 | 1 | 0 | 1 |
| 1021026 | 1 | 4 | 2 | 2 | 3 | 2 | 2 | 1 | 1 | 1 | 0 | 0 | 1 | 1 | 3 | 1 | 1 | 0 | 1 |
| 1021027 | 0 | 5 | 2 | 1 | 3 | 2 | 1 | 1 | 1 | 1 | 0 | 0 | 1 | 0 | 5 | 5 | 1 | 1 | 1 |
| 1021028 | 0 | 5 | 2 | 1 | 3 | 2 | 2 | 2 | 4 | 2 | 0 | 0 | 1 | 1 | 1 | 1 | 0 | 0 | 0 |
| 1021029 | 0 | 5 | 2 | 1 | 3 | 2 | 2 | 1 | 1 | 1 | 0 | 0 | 1 | 1 | 1 | 1 | 0 | 0 | 0 |
| 1021030 | 0 | 5 | 2 | 2 | 3 | 2 | 3 | 2 | 1 | 1 | 0 | 0 | 1 | 1 | 1 | 1 | 0 | 0 | 0 |
| 1021031 | 0 | 2 | 2 | 3 | 3 | 1 | 2 | 1 | 2 | 4 | 0 | 0 | 1 | 1 | 3 | 1 | 1 | 0 | 1 |
| 1021032 | 0 | 2 | 2 | 4 | 3 | 1 | 3 | 2 | 4 | 1 | 0 | 0 | 1 | 1 | 3 | 1 | 1 | 0 | 1 |
| 1021033 | 0 | 2 | 2 | 3 | 3 | 1 | 2 | 1 | 1 | 1 | 0 | 0 | 1 | 1 | 1 | 1 | 0 | 0 | 0 |
| 102111 | 0 | 3 | 2 | 2 | 3 | 2 | 1 | 1 | 2 | 1 | 0 | 0 | 0 | 99 | 99 | 99 | 99 | 99 | 99 |
| 102112 | 1 | 3 | 2 | 2 | 3 | 3 | 2 | 1 | 1 | 1 | 0 | 0 | 1 | 1 | 3 | 1 | 1 | 0 | 1 |
| 102113 | 0 | 4 | 2 | 1 | 3 | 2 | 2 | 1 | 2 | 1 | 0 | 0 | 1 | 1 | 2 | 1 | 1 | 0 | 1 |
| 102114 | 0 | 3 | 2 | 3 | 3 | 1 | 2 | 1 | 4 | 3 | 0 | 0 | 1 | 1 | 4 | 1 | 1 | 0 | 1 |
| 102115 | 1 | 2 | 2 | 2 | 3 | 4 | 2 | 1 | 4 | 3 | 0 | 0 | 1 | 1 | 1 | 1 | 0 | 0 | 0 |
| 102116 | 0 | 2 | 2 | 2 | 2 | 1 | 2 | 2 | 4 | 3 | 0 | 0 | 1 | 1 | 3 | 1 | 1 | 0 | 1 |
| 102117 | 1 | 2 | 2 | 5 | 3 | 5 | 3 | 1 | 4 | 1 | 1 | 0 | 1 | 1 | 1 | 1 | 0 | 0 | 0 |
| 102118 | 1 | 1 | 2 | 3 | 3 | 5 | 2 | 2 | 4 | 4 | 0 | 0 | 1 | 1 | 2 | 5 | 1 | 1 | 1 |
| 102119 | 1 | 5 | 2 | 4 | 3 | 4 | 2 | 1 | 2 | 1 | 0 | 0 | 1 | 1 | 1 | 1 | 0 | 0 | 0 |
| 1021110 | 0 | 4 | 2 | 1 | 3 | 2 | 2 | 1 | 4 | 2 | 0 | 0 | 0 | 99 | 99 | 99 | 99 | 99 | 99 |
| 1021111 | 1 | 2 | 2 | 4 | 3 | 4 | 2 | 1 | 4 | 2 | 0 | 0 | 1 | 0 | 5 | 5 | 1 | 1 | 1 |
| 1021112 | 1 | 2 | 2 | 3 | 3 | 2 | 2 | 1 | 4 | 2 | 0 | 0 | 1 | 1 | 1 | 1 | 0 | 0 | 0 |
| 1021113 | 1 | 1 | 2 | 1 | 2 | 4 | 2 | 1 | 2 | 1 | 0 | 0 | 1 | 0 | 5 | 5 | 1 | 1 | 1 |
| 1021114 | 0 | 3 | 2 | 1 | 3 | 2 | 1 | 1 | 2 | 1 | 0 | 0 | 1 | 0 | 5 | 5 | 1 | 1 | 1 |
| 1021115 | 0 | 3 | 2 | 2 | 3 | 1 | 2 | 1 | 2 | 1 | 0 | 0 | 1 | 1 | 3 | 1 | 1 | 0 | 1 |
| 1021116 | 0 | 3 | 2 | 2 | 3 | 1 | 2 | 2 | 1 | 1 | 0 | 0 | 1 | 0 | 5 | 5 | 1 | 1 | 1 |
| 1021117 | 1 | 5 | 2 | 2 | 2 | 5 | 2 | 1 | 4 | 3 | 1 | 1 | 1 | 1 | 1 | 1 | 0 | 0 | 0 |
| 1021118 | 0 | 4 | 2 | 2 | 3 | 1 | 2 | 1 | 4 | 3 | 0 | 0 | 1 | 1 | 1 | 1 | 0 | 0 | 0 |
| 1021119 | 0 | 4 | 2 | 2 | 3 | 1 | 2 | 2 | 4 | 3 | 0 | 0 | 1 | 1 | 1 | 1 | 0 | 0 | 0 |
| 1021120 | 1 | 3 | 2 | 1 | 3 | 2 | 2 | 2 | 4 | 2 | 0 | 0 | 1 | 1 | 4 | 1 | 1 | 0 | 1 |
| 1021121 | 1 | 5 | 2 | 1 | 3 | 2 | 1 | 1 | 4 | 1 | 0 | 0 | 0 | 99 | 99 | 99 | 99 | 99 | 99 |
| 1021122 | 1 | 5 | 2 | 1 | 3 | 2 | 3 | 2 | 4 | 2 | 0 | 0 | 1 | 1 | 3 | 1 | 1 | 0 | 1 |

| | | | | | | | | | | | | | | | | | | |
|---|---|---|---|---|---|---|---|---|---|---|---|---|---|---|---|---|---|---|
| 1021123 | 0 | 5 | 2 | 2 | 3 | 2 | 1 | 1 | 4 | 3 | 0 | 0 | 1 | 0 | 5 | 5 | 1 | 1 | 1 |
| 1021124 | 1 | 4 | 2 | 1 | 3 | 4 | 2 | 2 | 4 | 4 | 0 | 0 | 1 | 1 | 2 | 1 | 1 | 0 | 1 |
| 1021125 | 1 | 2 | 2 | 3 | 3 | 4 | 3 | 1 | 4 | 2 | 0 | 0 | 1 | 1 | 3 | 1 | 1 | 0 | 1 |
| 1021126 | 0 | 4 | 2 | 2 | 3 | 5 | 3 | 2 | 4 | 3 | 0 | 0 | 1 | 1 | 3 | 1 | 1 | 0 | 1 |
| 1021127 | 1 | 3 | 2 | 1 | 3 | 4 | 2 | 1 | 4 | 2 | 0 | 0 | 1 | 1 | 1 | 1 | 0 | 0 | 0 |
| 1021128 | 1 | 3 | 2 | 4 | 3 | 5 | 3 | 2 | 4 | 1 | 0 | 0 | 1 | 1 | 1 | 1 | 0 | 0 | 0 |
| 1021129 | 1 | 3 | 2 | 4 | 3 | 5 | 3 | 2 | 4 | 1 | 0 | 0 | 1 | 1 | 3 | 1 | 1 | 0 | 1 |
| 1021130 | 1 | 3 | 2 | 4 | 3 | 5 | 3 | 1 | 4 | 3 | 0 | 0 | 1 | 1 | 1 | 1 | 0 | 0 | 0 |
| 1021131 | 0 | 2 | 2 | 4 | 3 | 5 | 2 | 1 | 4 | 3 | 0 | 0 | 1 | 1 | 1 | 1 | 0 | 0 | 0 |
| 1021132 | 0 | 3 | 2 | 4 | 3 | 1 | 3 | 2 | 4 | 2 | 0 | 0 | 1 | 1 | 3 | 1 | 1 | 0 | 1 |
| 1021133 | 0 | 4 | 2 | 2 | 3 | 5 | 3 | 1 | 4 | 3 | 0 | 0 | 1 | 1 | 1 | 1 | 0 | 0 | 0 |
| 1021134 | 1 | 4 | 2 | 4 | 3 | 5 | 3 | 2 | 4 | 2 | 0 | 0 | 1 | 1 | 1 | 1 | 0 | 0 | 0 |
| 1021135 | 1 | 2 | 2 | 5 | 2 | 5 | 3 | 1 | 4 | 2 | 0 | 0 | 1 | 1 | 1 | 1 | 0 | 0 | 0 |
| 1021136 | 0 | 2 | 2 | 4 | 3 | 1 | 2 | 2 | 4 | 4 | 0 | 0 | 1 | 1 | 1 | 1 | 0 | 0 | 0 |
| 1021137 | 0 | 5 | 2 | 2 | 3 | 2 | 2 | 2 | 4 | 3 | 0 | 0 | 1 | 1 | 1 | 1 | 0 | 0 | 0 |
| 1021138 | 1 | 2 | 2 | 5 | 2 | 5 | 3 | 1 | 4 | 2 | 0 | 0 | 1 | 1 | 1 | 1 | 0 | 0 | 0 |
| 1021139 | 0 | 4 | 2 | 2 | 3 | 1 | 2 | 1 | 4 | 2 | 0 | 0 | 1 | 1 | 1 | 1 | 0 | 0 | 0 |
| 102121 | 0 | 5 | 2 | 1 | 3 | 2 | 2 | 1 | 2 | 1 | 0 | 0 | 0 | 99 | 99 | 99 | 99 | 99 | 99 |
| 102122 | 0 | 4 | 2 | 2 | 3 | 2 | 3 | 1 | 1 | 1 | 0 | 0 | 1 | 1 | 1 | 1 | 0 | 0 | 0 |
| 102123 | 1 | 5 | 2 | 5 | 2 | 5 | 3 | 1 | 2 | 1 | 0 | 0 | 1 | 1 | 1 | 1 | 0 | 0 | 0 |
| 102124 | 1 | 4 | 2 | 1 | 3 | 2 | 2 | 1 | 1 | 1 | 0 | 0 | 0 | 99 | 99 | 99 | 99 | 99 | 99 |
| 102125 | 0 | 3 | 2 | 3 | 3 | 1 | 2 | 2 | 4 | 1 | 0 | 0 | 1 | 1 | 3 | 1 | 1 | 0 | 1 |
| 102126 | 1 | 3 | 2 | 2 | 3 | 2 | 2 | 1 | 2 | 1 | 0 | 0 | 1 | 1 | 3 | 6 | 1 | 1 | 1 |
| 102127 | 0 | 3 | 2 | 1 | 3 | 1 | 2 | 2 | 1 | 1 | 0 | 0 | 1 | 1 | 3 | 1 | 1 | 0 | 1 |
| 102128 | 1 | 3 | 2 | 3 | 3 | 2 | 3 | 1 | 4 | 1 | 0 | 0 | 1 | 1 | 3 | 1 | 1 | 0 | 1 |
| 102129 | 0 | 1 | 2 | 4 | 3 | 1 | 3 | 1 | 4 | 1 | 0 | 0 | 1 | 1 | 3 | 1 | 1 | 0 | 1 |
| 1021210 | 1 | 3 | 2 | 2 | 3 | 2 | 3 | 2 | 2 | 1 | 0 | 0 | 1 | 1 | 1 | 1 | 0 | 0 | 0 |
| 1021211 | 0 | 1 | 2 | 5 | 3 | 5 | 3 | 1 | 2 | 3 | 0 | 0 | 1 | 1 | 1 | 1 | 0 | 0 | 0 |
| 1021212 | 1 | 2 | 2 | 2 | 3 | 2 | 3 | 2 | 2 | 3 | 0 | 0 | 1 | 1 | 2 | 1 | 1 | 0 | 1 |
| 1021213 | 0 | 5 | 2 | 2 | 3 | 1 | 2 | 2 | 2 | 2 | 0 | 0 | 1 | 1 | 1 | 1 | 0 | 0 | 0 |
| 1021214 | 0 | 4 | 2 | 2 | 3 | 1 | 2 | 2 | 2 | 2 | 0 | 0 | 1 | 1 | 4 | 1 | 1 | 0 | 1 |
| 1021215 | 0 | 4 | 2 | 2 | 3 | 1 | 2 | 2 | 2 | 2 | 0 | 0 | 1 | 1 | 4 | 1 | 1 | 0 | 1 |
| 1021216 | 1 | 5 | 2 | 1 | 3 | 2 | 2 | 2 | 2 | 2 | 0 | 0 | 1 | 1 | 3 | 5 | 1 | 1 | 1 |
| 1021217 | 0 | 4 | 2 | 2 | 3 | 1 | 2 | 1 | 2 | 1 | 0 | 0 | 1 | 1 | 4 | 1 | 1 | 0 | 1 |
| 1021218 | 1 | 1 | 2 | 3 | 3 | 4 | 3 | 1 | 2 | 1 | 0 | 0 | 1 | 1 | 3 | 5 | 1 | 1 | 1 |
| 1021219 | 1 | 3 | 2 | 3 | 3 | 2 | 2 | 2 | 2 | 1 | 0 | 0 | 1 | 1 | 1 | 5 | 0 | 1 | 1 |
| 1021220 | 1 | 4 | 2 | 2 | 3 | 2 | 2 | 1 | 2 | 1 | 0 | 0 | 1 | 0 | 5 | 5 | 1 | 1 | 1 |
| 1021221 | 1 | 5 | 2 | 2 | 3 | 2 | 3 | 1 | 1 | 1 | 0 | 0 | 1 | 1 | 1 | 1 | 0 | 0 | 0 |
| 1021222 | 0 | 4 | 2 | 1 | 3 | 1 | 2 | 2 | 2 | 2 | 0 | 0 | 1 | 1 | 1 | 1 | 0 | 0 | 0 |
| 1021223 | 1 | 2 | 2 | 5 | 3 | 5 | 3 | 1 | 4 | 1 | 0 | 0 | 1 | 1 | 2 | 1 | 1 | 0 | 1 |
| 1021224 | 0 | 4 | 2 | 5 | 3 | 5 | 3 | 2 | 4 | 1 | 1 | 1 | 1 | 1 | 2 | 1 | 1 | 0 | 1 |
| 1021225 | 0 | 5 | 2 | 2 | 3 | 1 | 2 | 2 | 2 | 1 | 0 | 0 | 1 | 1 | 2 | 1 | 1 | 0 | 1 |
| 1021226 | 0 | 4 | 2 | 1 | 3 | 1 | 2 | 1 | 2 | 2 | 0 | 0 | 1 | 0 | 5 | 5 | 1 | 1 | 1 |
| 1021227 | 1 | 2 | 2 | 1 | 3 | 4 | 2 | 1 | 2 | 2 | 0 | 0 | 1 | 1 | 1 | 1 | 0 | 0 | 0 |
| 1021228 | 1 | 5 | 2 | 1 | 3 | 2 | 2 | 2 | 2 | 1 | 0 | 0 | 1 | 0 | 5 | 5 | 1 | 1 | 1 |
| 1021229 | 0 | 2 | 2 | 2 | 4 | 1 | 2 | 2 | 4 | 1 | 0 | 0 | 1 | 1 | 3 | 1 | 1 | 0 | 1 |
| 1021230 | 1 | 2 | 2 | 2 | 3 | 2 | 2 | 2 | 2 | 1 | 0 | 0 | 1 | 1 | 4 | 1 | 1 | 0 | 1 |
| 1021231 | 1 | 3 | 2 | 1 | 3 | 4 | 2 | 2 | 2 | 1 | 0 | 0 | 1 | 1 | 3 | 1 | 1 | 0 | 1 |
| 1021232 | 1 | 4 | 2 | 1 | 2 | 4 | 2 | 1 | 2 | 1 | 0 | 0 | 1 | 0 | 5 | 5 | 1 | 1 | 1 |
| 1021233 | 1 | 1 | 2 | 5 | 3 | 3 | 3 | 1 | 2 | 1 | 1 | 1 | 1 | 1 | 2 | 1 | 1 | 0 | 1 |
| 1021234 | 0 | 5 | 2 | 1 | 2 | 1 | 2 | 2 | 4 | 2 | 0 | 0 | 1 | 1 | 1 | 1 | 0 | 0 | 0 |
| 1021235 | 0 | 2 | 2 | 5 | 3 | 5 | 2 | 2 | 4 | 2 | 0 | 0 | 1 | 1 | 1 | 1 | 0 | 0 | 0 |
| 103141 | 1 | 4 | 2 | 3 | 3 | 2 | 2 | 1 | 2 | 1 | 1 | 1 | 0 | 99 | 99 | 99 | 99 | 99 | 99 |
| 103142 | 1 | 2 | 2 | 3 | 3 | 2 | 2 | 2 | 2 | 1 | 0 | 0 | 0 | 99 | 99 | 99 | 99 | 99 | 99 |
| 103143 | 1 | 5 | 2 | 3 | 3 | 2 | 2 | 2 | 2 | 1 | 1 | 0 | 0 | 99 | 99 | 99 | 99 | 99 | 99 |
| 103144 | 1 | 3 | 2 | 3 | 3 | 2 | 2 | 2 | 2 | 1 | 0 | 0 | 1 | 1 | 1 | 1 | 0 | 0 | 0 |

| | | | | | | | | | | | | | | | | | | |
|---|---|---|---|---|---|---|---|---|---|---|---|---|---|---|---|---|---|---|
| 103145 | 1 | 3 | 2 | 3 | 3 | 2 | 2 | 1 | 2 | 1 | 0 | 0 | 0 | 99 | 99 | 99 | 99 | 99 | 99 |
| 103146 | 1 | 5 | 2 | 1 | 3 | 2 | 2 | 1 | 2 | 1 | 0 | 0 | 0 | 99 | 99 | 99 | 99 | 99 | 99 |
| 103147 | 1 | 5 | 2 | 1 | 3 | 2 | 1 | 2 | 2 | 1 | 0 | 0 | 0 | 99 | 99 | 99 | 99 | 99 | 99 |
| 103148 | 1 | 4 | 2 | 3 | 3 | 2 | 2 | 2 | 2 | 1 | 0 | 0 | 0 | 99 | 99 | 99 | 99 | 99 | 99 |
| 103149 | 1 | 3 | 2 | 2 | 3 | 2 | 2 | 2 | 2 | 1 | 1 | 0 | 0 | 99 | 99 | 99 | 99 | 99 | 99 |
| 1031410 | 1 | 4 | 2 | 4 | 3 | 2 | 2 | 1 | 2 | 1 | 0 | 0 | 1 | 1 | 1 | 1 | 0 | 0 | 0 |
| 1031411 | 0 | 5 | 2 | 2 | 3 | 1 | 2 | 1 | 3 | 1 | 1 | 1 | 0 | 99 | 99 | 99 | 99 | 99 | 99 |
| 1031412 | 0 | 3 | 2 | 2 | 3 | 1 | 1 | 1 | 3 | 1 | 0 | 0 | 0 | 99 | 99 | 99 | 99 | 99 | 99 |
| 1031413 | 0 | 4 | 2 | 1 | 3 | 1 | 2 | 1 | 3 | 1 | 0 | 0 | 0 | 99 | 99 | 99 | 99 | 99 | 99 |
| 1031414 | 0 | 3 | 2 | 2 | 3 | 1 | 2 | 1 | 3 | 1 | 0 | 0 | 0 | 99 | 99 | 99 | 99 | 99 | 99 |
| 1031415 | 0 | 3 | 2 | 1 | 3 | 1 | 2 | 1 | 3 | 1 | 0 | 0 | 1 | 1 | 1 | 1 | 0 | 0 | 0 |
| 1031416 | 0 | 3 | 2 | 2 | 3 | 1 | 2 | 1 | 3 | 1 | 0 | 0 | 0 | 99 | 99 | 99 | 99 | 99 | 99 |
| 1031417 | 0 | 5 | 2 | 2 | 3 | 1 | 1 | 1 | 3 | 1 | 0 | 0 | 0 | 99 | 99 | 99 | 99 | 99 | 99 |
| 1031418 | 0 | 5 | 2 | 2 | 3 | 1 | 2 | 2 | 3 | 1 | 0 | 0 | 0 | 99 | 99 | 99 | 99 | 99 | 99 |
| 1031419 | 0 | 4 | 2 | 2 | 3 | 1 | 1 | 1 | 3 | 1 | 0 | 0 | 0 | 99 | 99 | 99 | 99 | 99 | 99 |
| 1031420 | 0 | 5 | 2 | 2 | 3 | 1 | 2 | 2 | 3 | 1 | 0 | 0 | 0 | 99 | 99 | 99 | 99 | 99 | 99 |
| 1031421 | 0 | 4 | 2 | 2 | 3 | 1 | 1 | 1 | 3 | 1 | 0 | 0 | 0 | 99 | 99 | 99 | 99 | 99 | 99 |
| 1031422 | 0 | 4 | 2 | 2 | 3 | 1 | 1 | 1 | 3 | 1 | 0 | 0 | 0 | 99 | 99 | 99 | 99 | 99 | 99 |
| 1031423 | 0 | 1 | 2 | 2 | 3 | 1 | 1 | 1 | 3 | 1 | 0 | 0 | 1 | 1 | 1 | 1 | 0 | 0 | 0 |
| 1031424 | 0 | 4 | 2 | 1 | 3 | 1 | 2 | 2 | 3 | 1 | 0 | 0 | 0 | 99 | 99 | 99 | 99 | 99 | 99 |
| 1031425 | 1 | 4 | 2 | 1 | 3 | 2 | 1 | 1 | 3 | 1 | 0 | 0 | 0 | 99 | 99 | 99 | 99 | 99 | 99 |
| 1031426 | 0 | 2 | 2 | 2 | 3 | 1 | 1 | 1 | 3 | 1 | 0 | 0 | 0 | 99 | 99 | 99 | 99 | 99 | 99 |
| 1031427 | 0 | 3 | 2 | 2 | 3 | 1 | 1 | 1 | 3 | 1 | 0 | 0 | 1 | 1 | 1 | 1 | 0 | 0 | 0 |
| 1031428 | 0 | 1 | 2 | 4 | 3 | 1 | 1 | 1 | 3 | 1 | 0 | 0 | 0 | 99 | 99 | 99 | 99 | 99 | 99 |
| 1031429 | 0 | 4 | 2 | 3 | 3 | 1 | 2 | 1 | 3 | 1 | 0 | 0 | 1 | 1 | 3 | 1 | 1 | 0 | 1 |
| 1031430 | 0 | 3 | 2 | 2 | 3 | 1 | 1 | 1 | 3 | 1 | 0 | 0 | 0 | 99 | 99 | 99 | 99 | 99 | 99 |
| 103131 | 0 | 3 | 2 | 2 | 3 | 1 | 2 | 1 | 2 | 1 | 0 | 0 | 1 | 1 | 1 | 1 | 0 | 0 | 0 |
| 103132 | 0 | 2 | 2 | 5 | 3 | 5 | 2 | 2 | 2 | 4 | 0 | 0 | 1 | 1 | 1 | 1 | 0 | 0 | 0 |
| 103133 | 0 | 2 | 2 | 2 | 3 | 1 | 2 | 2 | 2 | 1 | 0 | 0 | 1 | 1 | 4 | 1 | 1 | 0 | 1 |
| 103134 | 0 | 2 | 2 | 3 | 3 | 1 | 3 | 2 | 2 | 1 | 0 | 0 | 1 | 1 | 1 | 1 | 0 | 0 | 0 |
| 103135 | 0 | 2 | 2 | 5 | 3 | 5 | 3 | 2 | 2 | 1 | 0 | 0 | 1 | 1 | 1 | 1 | 0 | 0 | 0 |
| 103136 | 1 | 4 | 2 | 2 | 3 | 2 | 1 | 1 | 2 | 1 | 0 | 0 | 1 | 1 | 4 | 1 | 1 | 0 | 1 |
| 103137 | 0 | 3 | 2 | 2 | 3 | 1 | 1 | 2 | 2 | 1 | 0 | 0 | 1 | 1 | 4 | 1 | 1 | 0 | 1 |
| 103138 | 0 | 2 | 2 | 2 | 3 | 1 | 1 | 2 | 2 | 1 | 0 | 0 | 0 | 99 | 99 | 99 | 99 | 99 | 99 |
| 103139 | 0 | 3 | 2 | 4 | 3 | 1 | 1 | 1 | 2 | 1 | 0 | 0 | 1 | 1 | 1 | 1 | 0 | 0 | 0 |
| 1031310 | 1 | 5 | 2 | 2 | 3 | 2 | 1 | 1 | 2 | 1 | 0 | 0 | 1 | 1 | 3 | 1 | 1 | 0 | 1 |
| 1031311 | 0 | 4 | 2 | 2 | 3 | 1 | 2 | 1 | 2 | 1 | 0 | 0 | 1 | 1 | 3 | 1 | 1 | 0 | 1 |
| 1031312 | 1 | 4 | 2 | 2 | 3 | 5 | 1 | 1 | 2 | 1 | 0 | 0 | 0 | 99 | 99 | 99 | 99 | 99 | 99 |
| 1031313 | 1 | 3 | 2 | 4 | 3 | 2 | 2 | 2 | 2 | 1 | 0 | 0 | 1 | 1 | 4 | 1 | 1 | 0 | 1 |
| 1031314 | 1 | 2 | 2 | 4 | 3 | 2 | 2 | 2 | 2 | 1 | 0 | 0 | 1 | 1 | 4 | 1 | 1 | 0 | 1 |
| 1031315 | 0 | 4 | 2 | 2 | 3 | 1 | 1 | 1 | 2 | 1 | 0 | 0 | 1 | 1 | 3 | 1 | 1 | 0 | 1 |
| 1031316 | 0 | 4 | 2 | 2 | 3 | 1 | 1 | 1 | 2 | 1 | 0 | 0 | 1 | 1 | 4 | 1 | 1 | 0 | 1 |
| 1031317 | 1 | 2 | 2 | 2 | 3 | 2 | 2 | 2 | 2 | 1 | 0 | 0 | 0 | 99 | 99 | 99 | 99 | 99 | 99 |
| 1031318 | 0 | 3 | 2 | 2 | 3 | 2 | 1 | 1 | 2 | 1 | 0 | 0 | 1 | 1 | 4 | 1 | 1 | 0 | 1 |
| 1031319 | 0 | 1 | 2 | 4 | 3 | 5 | 3 | 1 | 2 | 1 | 0 | 0 | 1 | 1 | 4 | 1 | 1 | 0 | 1 |
| 1031320 | 0 | 1 | 2 | 4 | 3 | 1 | 2 | 1 | 2 | 1 | 0 | 0 | 1 | 1 | 1 | 1 | 0 | 0 | 0 |
| 1031321 | 0 | 5 | 2 | 2 | 3 | 1 | 1 | 1 | 2 | 1 | 0 | 0 | 0 | 99 | 99 | 99 | 99 | 99 | 99 |
| 1031322 | 0 | 5 | 2 | 2 | 3 | 1 | 2 | 1 | 2 | 1 | 0 | 0 | 0 | 99 | 99 | 99 | 99 | 99 | 99 |
| 1031323 | 0 | 2 | 2 | 2 | 3 | 1 | 2 | 2 | 2 | 1 | 0 | 0 | 0 | 99 | 99 | 99 | 99 | 99 | 99 |
| 1031324 | 0 | 3 | 2 | 2 | 3 | 1 | 2 | 1 | 2 | 1 | 0 | 0 | 1 | 1 | 4 | 1 | 1 | 0 | 1 |
| 1031325 | 0 | 4 | 2 | 2 | 3 | 1 | 2 | 1 | 2 | 1 | 0 | 0 | 1 | 1 | 3 | 1 | 1 | 0 | 1 |
| 1031326 | 0 | 2 | 2 | 2 | 3 | 1 | 2 | 2 | 2 | 1 | 0 | 0 | 1 | 1 | 1 | 1 | 0 | 0 | 0 |
| 1031327 | 0 | 3 | 2 | 2 | 3 | 1 | 2 | 2 | 2 | 1 | 0 | 0 | 1 | 1 | 4 | 1 | 1 | 0 | 1 |
| 1031328 | 0 | 5 | 2 | 2 | 3 | 1 | 1 | 1 | 2 | 1 | 0 | 0 | 0 | 99 | 99 | 99 | 99 | 99 | 99 |
| 1031329 | 0 | 3 | 2 | 2 | 3 | 1 | 1 | 2 | 2 | 1 | 0 | 0 | 1 | 1 | 3 | 1 | 1 | 0 | 1 |
| 1031330 | 0 | 3 | 2 | 2 | 3 | 1 | 2 | 1 | 2 | 1 | 0 | 0 | 0 | 99 | 99 | 99 | 99 | 99 | 99 |

| | | | | | | | | | | | | | | | | | | | |
|---|---|---|---|---|---|---|---|---|---|---|---|---|---|---|---|---|---|---|---|
| 103151 | 0 | 4 | 2 | 1 | 3 | 1 | 1 | 1 | 2 | 1 | 0 | 0 | 0 | 99 | 99 | 99 | 99 | 99 | 99 |
| 103152 | 0 | 4 | 2 | 2 | 3 | 1 | 2 | 1 | 2 | 1 | 0 | 0 | 1 | 1 | 3 | 1 | 1 | 0 | 1 |
| 103153 | 0 | 4 | 2 | 2 | 3 | 1 | 2 | 2 | 2 | 1 | 0 | 0 | 0 | 99 | 99 | 99 | 99 | 99 | 99 |
| 103154 | 0 | 1 | 2 | 2 | 3 | 1 | 2 | 2 | 2 | 1 | 0 | 0 | 0 | 99 | 99 | 99 | 99 | 99 | 99 |
| 103155 | 0 | 2 | 2 | 2 | 3 | 1 | 2 | 1 | 2 | 1 | 0 | 0 | 1 | 1 | 3 | 1 | 1 | 0 | 1 |
| 103156 | 1 | 4 | 2 | 2 | 3 | 2 | 2 | 1 | 2 | 1 | 0 | 0 | 0 | 99 | 99 | 99 | 99 | 99 | 99 |
| 103157 | 0 | 1 | 2 | 4 | 3 | 1 | 1 | 1 | 2 | 1 | 0 | 0 | 0 | 99 | 99 | 99 | 99 | 99 | 99 |
| 103158 | 0 | 4 | 2 | 2 | 3 | 1 | 1 | 1 | 2 | 1 | 0 | 0 | 0 | 99 | 99 | 99 | 99 | 99 | 99 |
| 103159 | 0 | 5 | 2 | 2 | 3 | 1 | 2 | 1 | 2 | 1 | 1 | 1 | 0 | 99 | 99 | 99 | 99 | 99 | 99 |
| 1031510 | 0 | 3 | 2 | 2 | 3 | 1 | 2 | 2 | 2 | 1 | 0 | 0 | 0 | 99 | 99 | 99 | 99 | 99 | 99 |
| 1031511 | 0 | 2 | 2 | 3 | 3 | 1 | 2 | 2 | 2 | 1 | 0 | 0 | 0 | 99 | 99 | 99 | 99 | 99 | 99 |
| 1031512 | 0 | 2 | 2 | 2 | 3 | 1 | 1 | 2 | 2 | 1 | 0 | 0 | 0 | 99 | 99 | 99 | 99 | 99 | 99 |
| 1031513 | 0 | 1 | 2 | 4 | 3 | 1 | 1 | 2 | 2 | 1 | 0 | 0 | 0 | 99 | 99 | 99 | 99 | 99 | 99 |
| 1031514 | 0 | 1 | 2 | 3 | 3 | 1 | 1 | 1 | 2 | 1 | 0 | 0 | 0 | 99 | 99 | 99 | 99 | 99 | 99 |
| 1031515 | 0 | 1 | 2 | 4 | 3 | 1 | 1 | 1 | 2 | 1 | 0 | 0 | 0 | 99 | 99 | 99 | 99 | 99 | 99 |
| 1031516 | 0 | 2 | 2 | 3 | 3 | 1 | 1 | 2 | 2 | 1 | 0 | 0 | 1 | 1 | 2 | 1 | 1 | 0 | 1 |
| 1031517 | 0 | 4 | 2 | 2 | 3 | 1 | 1 | 1 | 2 | 1 | 0 | 0 | 0 | 99 | 99 | 99 | 99 | 99 | 99 |
| 1031518 | 0 | 4 | 2 | 3 | 3 | 1 | 2 | 2 | 2 | 1 | 0 | 0 | 1 | 1 | 1 | 1 | 0 | 0 | 0 |
| 1031519 | 0 | 1 | 2 | 4 | 3 | 1 | 1 | 1 | 2 | 1 | 0 | 0 | 1 | 1 | 1 | 1 | 0 | 0 | 0 |
| 1031520 | 1 | 4 | 2 | 2 | 3 | 2 | 2 | 1 | 2 | 1 | 0 | 0 | 0 | 99 | 99 | 99 | 99 | 99 | 99 |
| 1031521 | 0 | 4 | 2 | 1 | 3 | 1 | 1 | 1 | 2 | 1 | 0 | 0 | 0 | 99 | 99 | 99 | 99 | 99 | 99 |
| 1031522 | 1 | 5 | 2 | 2 | 3 | 2 | 2 | 2 | 2 | 1 | 0 | 0 | 0 | 99 | 99 | 99 | 99 | 99 | 99 |
| 1031523 | 1 | 5 | 2 | 2 | 3 | 2 | 3 | 2 | 2 | 1 | 1 | 0 | 1 | 1 | 1 | 1 | 0 | 0 | 0 |
| 1031524 | 0 | 5 | 2 | 1 | 3 | 1 | 2 | 1 | 2 | 1 | 0 | 0 | 0 | 99 | 99 | 99 | 99 | 99 | 99 |
| 1031525 | 1 | 4 | 2 | 1 | 3 | 2 | 2 | 1 | 2 | 1 | 0 | 0 | 0 | 99 | 99 | 99 | 99 | 99 | 99 |
| 1031526 | 0 | 5 | 2 | 1 | 3 | 1 | 1 | 1 | 2 | 1 | 0 | 0 | 0 | 99 | 99 | 99 | 99 | 99 | 99 |
| 1031527 | 0 | 4 | 2 | 2 | 3 | 1 | 2 | 1 | 2 | 1 | 0 | 0 | 0 | 99 | 99 | 99 | 99 | 99 | 99 |
| 1031528 | 0 | 1 | 2 | 3 | 3 | 1 | 2 | 2 | 2 | 1 | 0 | 0 | 1 | 1 | 1 | 1 | 0 | 0 | 0 |
| 1031529 | 0 | 3 | 2 | 2 | 3 | 1 | 2 | 1 | 2 | 1 | 0 | 0 | 1 | 1 | 2 | 1 | 1 | 0 | 1 |
| 1031530 | 0 | 2 | 2 | 5 | 3 | 5 | 3 | 1 | 2 | 1 | 0 | 0 | 1 | 1 | 1 | 1 | 0 | 0 | 0 |
| 103161 | 0 | 3 | 2 | 2 | 3 | 1 | 2 | 2 | 1 | 1 | 0 | 0 | 0 | 99 | 99 | 99 | 99 | 99 | 99 |
| 103162 | 0 | 3 | 2 | 3 | 3 | 1 | 2 | 2 | 1 | 1 | 0 | 0 | 0 | 99 | 99 | 99 | 99 | 99 | 99 |
| 103163 | 0 | 2 | 2 | 3 | 3 | 1 | 3 | 2 | 1 | 1 | 0 | 0 | 1 | 1 | 1 | 1 | 0 | 0 | 0 |
| 103164 | 0 | 2 | 2 | 3 | 3 | 1 | 2 | 1 | 1 | 1 | 0 | 0 | 1 | 1 | 1 | 1 | 0 | 0 | 0 |
| 103165 | 0 | 4 | 2 | 4 | 3 | 1 | 1 | 1 | 1 | 1 | 0 | 0 | 1 | 1 | 1 | 1 | 0 | 0 | 0 |
| 103166 | 0 | 4 | 2 | 2 | 3 | 1 | 1 | 1 | 1 | 1 | 0 | 0 | 1 | 1 | 1 | 1 | 0 | 0 | 0 |
| 103167 | 0 | 2 | 2 | 2 | 3 | 1 | 1 | 2 | 1 | 1 | 0 | 0 | 0 | 99 | 99 | 99 | 99 | 99 | 99 |
| 103168 | 0 | 5 | 2 | 2 | 3 | 1 | 2 | 1 | 1 | 1 | 0 | 0 | 1 | 1 | 1 | 1 | 0 | 0 | 0 |
| 103169 | 0 | 5 | 2 | 2 | 3 | 1 | 1 | 2 | 1 | 1 | 0 | 0 | 0 | 99 | 99 | 99 | 99 | 99 | 99 |
| 1031610 | 0 | 1 | 2 | 4 | 3 | 1 | 2 | 1 | 1 | 1 | 0 | 0 | 0 | 99 | 99 | 99 | 99 | 99 | 99 |
| 1031611 | 0 | 1 | 2 | 2 | 3 | 1 | 1 | 2 | 1 | 1 | 0 | 0 | 1 | 1 | 4 | 1 | 1 | 0 | 1 |
| 1031612 | 0 | 2 | 2 | 2 | 3 | 1 | 2 | 1 | 1 | 1 | 1 | 1 | 1 | 1 | 3 | 1 | 1 | 0 | 1 |
| 1031613 | 0 | 3 | 2 | 2 | 3 | 1 | 2 | 2 | 1 | 1 | 0 | 0 | 1 | 1 | 4 | 1 | 1 | 0 | 1 |
| 1031614 | 0 | 3 | 2 | 2 | 3 | 1 | 1 | 1 | 1 | 1 | 0 | 0 | 1 | 1 | 2 | 1 | 1 | 0 | 1 |
| 1031615 | 0 | 5 | 2 | 2 | 3 | 1 | 1 | 2 | 1 | 1 | 0 | 0 | 1 | 1 | 1 | 1 | 0 | 0 | 0 |
| 1031616 | 0 | 4 | 2 | 2 | 3 | 1 | 1 | 2 | 1 | 1 | 0 | 0 | 0 | 99 | 99 | 99 | 99 | 99 | 99 |
| 1031617 | 0 | 4 | 2 | 2 | 3 | 1 | 2 | 2 | 1 | 1 | 1 | 1 | 0 | 99 | 99 | 99 | 99 | 99 | 99 |
| 1031618 | 0 | 4 | 2 | 2 | 3 | 1 | 1 | 2 | 1 | 1 | 0 | 0 | 0 | 99 | 99 | 99 | 99 | 99 | 99 |
| 1031619 | 0 | 5 | 2 | 1 | 3 | 1 | 2 | 2 | 1 | 1 | 1 | 1 | 0 | 99 | 99 | 99 | 99 | 99 | 99 |
| 1031620 | 0 | 3 | 2 | 3 | 3 | 1 | 2 | 1 | 1 | 1 | 0 | 0 | 0 | 99 | 99 | 99 | 99 | 99 | 99 |
| 1031621 | 0 | 1 | 2 | 5 | 3 | 5 | 3 | 1 | 1 | 1 | 1 | 1 | 1 | 1 | 2 | 1 | 1 | 0 | 1 |
| 1031622 | 0 | 4 | 2 | 4 | 3 | 1 | 2 | 2 | 1 | 1 | 1 | 1 | 0 | 99 | 99 | 99 | 99 | 99 | 99 |
| 1031623 | 0 | 4 | 2 | 3 | 3 | 1 | 3 | 1 | 1 | 1 | 1 | 1 | 1 | 1 | 1 | 1 | 0 | 0 | 0 |
| 204171 | 0 | 1 | 1 | 4 | 1 | 1 | 2 | 2 | 4 | 4 | 0 | 0 | 1 | 1 | 1 | 1 | 0 | 0 | 0 |
| 204172 | 0 | 3 | 1 | 1 | 1 | 2 | 2 | 1 | 4 | 4 | 0 | 0 | 1 | 1 | 1 | 1 | 0 | 0 | 0 |
| 204173 | 0 | 1 | 1 | 2 | 1 | 1 | 2 | 1 | 4 | 4 | 0 | 0 | 1 | 1 | 2 | 1 | 1 | 0 | 1 |

| | | | | | | | | | | | | | | | | | | | |
|---|---|---|---|---|---|---|---|---|---|---|---|---|---|---|---|---|---|---|---|
| 204174 | 1 | 2 | 1 | 2 | 1 | 2 | 2 | 2 | 4 | 4 | 0 | 0 | 1 | 1 | 3 | 1 | 1 | 0 | 1 |
| 204175 | 0 | 1 | 1 | 4 | 1 | 2 | 2 | 1 | 4 | 4 | 0 | 0 | 1 | 1 | 2 | 1 | 1 | 0 | 1 |
| 204176 | 1 | 5 | 1 | 2 | 1 | 2 | 2 | 2 | 4 | 4 | 0 | 0 | 1 | 1 | 2 | 1 | 1 | 0 | 1 |
| 204177 | 1 | 4 | 1 | 2 | 1 | 2 | 1 | 2 | 4 | 1 | 0 | 0 | 1 | 1 | 1 | 4 | 0 | 1 | 1 |
| 204178 | 1 | 2 | 1 | 1 | 1 | 2 | 2 | 1 | 4 | 4 | 0 | 0 | 1 | 1 | 3 | 1 | 1 | 0 | 1 |
| 204179 | 0 | 1 | 1 | 2 | 1 | 1 | 2 | 2 | 4 | 2 | 0 | 0 | 1 | 1 | 3 | 1 | 1 | 0 | 1 |
| 2041710 | 0 | 1 | 1 | 5 | 1 | 1 | 2 | 2 | 4 | 1 | 0 | 0 | 1 | 1 | 1 | 4 | 0 | 1 | 1 |
| 2041711 | 1 | 4 | 1 | 2 | 1 | 2 | 2 | 2 | 4 | 4 | 0 | 0 | 1 | 1 | 3 | 4 | 1 | 1 | 1 |
| 2041712 | 1 | 4 | 1 | 2 | 1 | 4 | 2 | 2 | 4 | 4 | 0 | 0 | 1 | 1 | 3 | 4 | 1 | 1 | 1 |
| 2041713 | 0 | 4 | 1 | 2 | 1 | 1 | 2 | 1 | 4 | 4 | 0 | 0 | 1 | 1 | 1 | 4 | 0 | 1 | 1 |
| 2041714 | 1 | 5 | 1 | 2 | 1 | 2 | 1 | 1 | 4 | 3 | 0 | 0 | 1 | 1 | 4 | 4 | 1 | 1 | 1 |
| 2041715 | 1 | 5 | 1 | 2 | 1 | 2 | 2 | 1 | 4 | 1 | 0 | 0 | 1 | 1 | 4 | 4 | 1 | 1 | 1 |
| 2041716 | 1 | 5 | 1 | 2 | 1 | 2 | 2 | 2 | 4 | 1 | 0 | 0 | 1 | 1 | 4 | 4 | 1 | 1 | 1 |
| 2041717 | 1 | 4 | 1 | 3 | 1 | 5 | 3 | 2 | 4 | 1 | 1 | 1 | 1 | 1 | 1 | 1 | 0 | 0 | 0 |
| 2041718 | 0 | 4 | 1 | 1 | 1 | 1 | 2 | 2 | 4 | 2 | 0 | 0 | 1 | 1 | 4 | 4 | 1 | 1 | 1 |
| 2041719 | 0 | 3 | 1 | 2 | 1 | 1 | 2 | 2 | 4 | 1 | 0 | 0 | 1 | 1 | 3 | 1 | 1 | 0 | 1 |
| 2041720 | 1 | 5 | 1 | 2 | 1 | 2 | 2 | 2 | 4 | 1 | 0 | 0 | 1 | 1 | 4 | 1 | 1 | 0 | 1 |
| 2041721 | 1 | 4 | 1 | 1 | 1 | 2 | 2 | 2 | 4 | 1 | 0 | 0 | 1 | 1 | 3 | 4 | 1 | 1 | 1 |
| 2041722 | 1 | 2 | 1 | 1 | 1 | 2 | 2 | 2 | 4 | 1 | 0 | 0 | 1 | 1 | 3 | 4 | 1 | 1 | 1 |
| 2041723 | 0 | 2 | 1 | 3 | 1 | 1 | 2 | 2 | 4 | 1 | 0 | 0 | 1 | 1 | 1 | 1 | 0 | 0 | 0 |
| 2041724 | 0 | 5 | 1 | 1 | 1 | 1 | 2 | 1 | 4 | 1 | 0 | 0 | 1 | 1 | 3 | 4 | 1 | 1 | 1 |
| 2041725 | 1 | 5 | 1 | 1 | 1 | 2 | 2 | 2 | 4 | 2 | 0 | 0 | 1 | 1 | 1 | 4 | 0 | 1 | 1 |
| 2041726 | 1 | 2 | 1 | 2 | 1 | 2 | 2 | 1 | 4 | 1 | 0 | 0 | 1 | 1 | 3 | 1 | 1 | 0 | 1 |
| 2041727 | 1 | 5 | 1 | 1 | 1 | 2 | 2 | 2 | 4 | 2 | 0 | 0 | 1 | 1 | 3 | 1 | 1 | 0 | 1 |
| 2041728 | 1 | 3 | 1 | 2 | 1 | 2 | 3 | 2 | 4 | 1 | 0 | 0 | 1 | 1 | 1 | 1 | 0 | 0 | 0 |
| 2041729 | 1 | 2 | 1 | 2 | 1 | 2 | 2 | 2 | 4 | 3 | 0 | 0 | 1 | 1 | 3 | 4 | 1 | 1 | 1 |
| 2041730 | 0 | 4 | 1 | 2 | 1 | 2 | 1 | 1 | 4 | 3 | 0 | 0 | 1 | 1 | 4 | 4 | 1 | 1 | 1 |
| 2041731 | 1 | 1 | 1 | 2 | 1 | 2 | 2 | 1 | 4 | 3 | 0 | 0 | 1 | 1 | 1 | 1 | 0 | 0 | 0 |
| 2041732 | 1 | 2 | 1 | 5 | 1 | 2 | 2 | 1 | 4 | 4 | 0 | 0 | 1 | 1 | 2 | 1 | 1 | 0 | 1 |
| 2041733 | 1 | 2 | 1 | 3 | 1 | 2 | 2 | 1 | 4 | 4 | 0 | 0 | 1 | 1 | 2 | 1 | 1 | 0 | 1 |
| 2041734 | 1 | 4 | 1 | 2 | 1 | 2 | 1 | 1 | 4 | 4 | 0 | 0 | 1 | 1 | 3 | 1 | 1 | 0 | 1 |
| 2041735 | 0 | 3 | 1 | 2 | 1 | 1 | 2 | 2 | 4 | 1 | 0 | 0 | 1 | 1 | 2 | 4 | 1 | 1 | 1 |
| 204181 | 1 | 3 | 1 | 2 | 1 | 2 | 3 | 2 | 1 | 2 | 0 | 0 | 1 | 1 | 3 | 4 | 1 | 1 | 1 |
| 204182 | 1 | 4 | 1 | 2 | 1 | 2 | 1 | 2 | 1 | 1 | 0 | 0 | 1 | 1 | 3 | 1 | 1 | 0 | 1 |
| 204183 | 1 | 3 | 1 | 2 | 1 | 2 | 2 | 2 | 1 | 3 | 0 | 0 | 1 | 1 | 4 | 4 | 1 | 1 | 1 |
| 204184 | 1 | 4 | 1 | 2 | 1 | 2 | 2 | 1 | 1 | 2 | 0 | 0 | 1 | 1 | 4 | 4 | 1 | 1 | 1 |
| 204185 | 0 | 5 | 1 | 1 | 1 | 2 | 2 | 1 | 1 | 1 | 0 | 0 | 1 | 1 | 3 | 1 | 1 | 0 | 1 |
| 204186 | 0 | 5 | 1 | 1 | 1 | 2 | 2 | 1 | 2 | 1 | 0 | 0 | 1 | 1 | 3 | 4 | 1 | 1 | 1 |
| 204187 | 1 | 2 | 1 | 5 | 1 | 2 | 2 | 2 | 4 | 2 | 0 | 0 | 1 | 1 | 1 | 1 | 0 | 0 | 0 |
| 204188 | 1 | 1 | 1 | 4 | 1 | 2 | 2 | 1 | 1 | 2 | 0 | 0 | 1 | 1 | 3 | 1 | 1 | 0 | 1 |
| 204189 | 1 | 4 | 1 | 2 | 1 | 2 | 2 | 1 | 1 | 1 | 0 | 0 | 1 | 1 | 3 | 4 | 1 | 1 | 1 |
| 2041810 | 1 | 5 | 1 | 3 | 1 | 2 | 1 | 2 | 1 | 2 | 0 | 0 | 1 | 1 | 1 | 1 | 0 | 0 | 0 |
| 2041811 | 1 | 4 | 1 | 4 | 1 | 2 | 3 | 2 | 4 | 2 | 0 | 0 | 1 | 1 | 3 | 4 | 1 | 1 | 1 |
| 2041812 | 0 | 3 | 1 | 2 | 1 | 2 | 3 | 1 | 1 | 1 | 0 | 0 | 1 | 1 | 1 | 6 | 0 | 1 | 1 |
| 2041813 | 0 | 3 | 1 | 2 | 1 | 2 | 2 | 1 | 4 | 2 | 0 | 0 | 1 | 1 | 4 | 1 | 1 | 0 | 1 |
| 2041814 | 1 | 3 | 1 | 4 | 1 | 2 | 2 | 1 | 3 | 4 | 0 | 0 | 1 | 1 | 1 | 1 | 0 | 0 | 0 |
| 2041815 | 1 | 1 | 1 | 5 | 1 | 2 | 1 | 1 | 4 | 4 | 0 | 0 | 1 | 1 | 3 | 1 | 1 | 0 | 1 |
| 2041816 | 1 | 3 | 1 | 2 | 1 | 2 | 2 | 1 | 4 | 4 | 0 | 0 | 1 | 1 | 3 | 1 | 1 | 0 | 1 |
| 2041817 | 1 | 4 | 1 | 2 | 1 | 2 | 2 | 2 | 4 | 4 | 0 | 0 | 1 | 1 | 1 | 1 | 0 | 0 | 0 |
| 2041818 | 1 | 5 | 1 | 2 | 1 | 2 | 2 | 2 | 1 | 1 | 0 | 0 | 1 | 1 | 3 | 1 | 1 | 0 | 1 |
| 2041819 | 1 | 2 | 1 | 5 | 1 | 5 | 2 | 1 | 4 | 4 | 1 | 1 | 1 | 1 | 1 | 1 | 0 | 0 | 0 |
| 2041820 | 0 | 4 | 1 | 1 | 1 | 2 | 3 | 2 | 1 | 1 | 0 | 0 | 1 | 1 | 4 | 1 | 1 | 0 | 1 |
| 2041821 | 1 | 3 | 1 | 3 | 1 | 2 | 2 | 2 | 4 | 2 | 0 | 0 | 1 | 1 | 3 | 1 | 1 | 0 | 1 |
| 2041822 | 1 | 4 | 1 | 1 | 1 | 2 | 2 | 1 | 4 | 4 | 0 | 0 | 1 | 1 | 2 | 1 | 1 | 0 | 1 |
| 2041823 | 1 | 5 | 1 | 1 | 1 | 2 | 2 | 2 | 1 | 1 | 0 | 0 | 1 | 1 | 3 | 1 | 1 | 0 | 1 |
| 2041824 | 1 | 1 | 1 | 4 | 1 | 2 | 2 | 1 | 1 | 1 | 0 | 0 | 1 | 1 | 1 | 4 | 0 | 1 | 1 |

| | | | | | | | | | | | | | | | | | | | |
|---|---|---|---|---|---|---|---|---|---|---|---|---|---|---|---|---|---|---|---|
| 2041825 | 0 | 1 | 1 | 3 | 1 | 1 | 2 | 1 | 1 | 1 | 0 | 0 | 1 | 1 | 3 | 4 | 1 | 1 | 1 |
| 2041826 | 1 | 3 | 1 | 2 | 1 | 2 | 2 | 2 | 1 | 1 | 0 | 0 | 1 | 1 | 3 | 4 | 1 | 1 | 1 |
| 2041827 | 1 | 5 | 1 | 2 | 1 | 2 | 2 | 1 | 1 | 1 | 0 | 0 | 1 | 1 | 3 | 4 | 1 | 1 | 1 |
| 2041828 | 1 | 5 | 1 | 1 | 1 | 2 | 2 | 2 | 4 | 4 | 0 | 0 | 1 | 1 | 3 | 4 | 1 | 1 | 1 |
| 2041829 | 1 | 1 | 1 | 4 | 1 | 2 | 2 | 1 | 4 | 4 | 0 | 0 | 1 | 1 | 4 | 1 | 1 | 0 | 1 |
| 2041830 | 1 | 4 | 1 | 2 | 1 | 1 | 2 | 2 | 1 | 1 | 0 | 0 | 1 | 1 | 4 | 1 | 1 | 0 | 1 |
| 204191 | 1 | 2 | 1 | 4 | 1 | 2 | 2 | 2 | 3 | 4 | 0 | 0 | 1 | 1 | 1 | 5 | 0 | 1 | 1 |
| 204192 | 1 | 1 | 1 | 5 | 1 | 2 | 2 | 1 | 1 | 2 | 0 | 0 | 1 | 1 | 3 | 1 | 1 | 0 | 1 |
| 204193 | 0 | 2 | 1 | 2 | 1 | 2 | 1 | 1 | 3 | 4 | 0 | 0 | 1 | 1 | 3 | 1 | 1 | 0 | 1 |
| 204194 | 1 | 5 | 1 | 1 | 1 | 2 | 1 | 1 | 3 | 4 | 0 | 0 | 1 | 1 | 3 | 7 | 1 | 1 | 1 |
| 204195 | 0 | 3 | 1 | 2 | 1 | 2 | 2 | 2 | 1 | 3 | 0 | 0 | 1 | 1 | 1 | 5 | 0 | 1 | 1 |
| 204196 | 0 | 4 | 1 | 2 | 1 | 2 | 1 | 1 | 1 | 2 | 0 | 0 | 1 | 1 | 3 | 1 | 1 | 0 | 1 |
| 204197 | 0 | 5 | 1 | 1 | 1 | 2 | 1 | 1 | 1 | 2 | 0 | 0 | 1 | 1 | 2 | 1 | 1 | 0 | 1 |
| 204198 | 0 | 3 | 1 | 4 | 1 | 1 | 2 | 1 | 1 | 2 | 0 | 0 | 1 | 1 | 3 | 7 | 1 | 1 | 1 |
| 204199 | 0 | 5 | 1 | 1 | 1 | 2 | 1 | 1 | 1 | 4 | 0 | 0 | 0 | 99 | 99 | 99 | 99 | 99 | 99 |
| 2041910 | 0 | 3 | 1 | 2 | 1 | 2 | 1 | 2 | 3 | 4 | 0 | 0 | 1 | 1 | 3 | 5 | 1 | 1 | 1 |
| 2041911 | 0 | 3 | 1 | 2 | 1 | 2 | 2 | 2 | 1 | 2 | 0 | 0 | 1 | 1 | 2 | 7 | 1 | 1 | 1 |
| 2041912 | 1 | 2 | 1 | 3 | 1 | 2 | 2 | 1 | 3 | 4 | 0 | 0 | 1 | 1 | 3 | 1 | 1 | 0 | 1 |
| 2041913 | 1 | 5 | 1 | 1 | 1 | 2 | 1 | 1 | 3 | 4 | 0 | 0 | 1 | 1 | 3 | 6 | 1 | 1 | 1 |
| 2041914 | 1 | 5 | 1 | 1 | 1 | 2 | 1 | 1 | 3 | 4 | 0 | 0 | 1 | 1 | 3 | 1 | 1 | 0 | 1 |
| 2041915 | 0 | 3 | 1 | 2 | 1 | 2 | 1 | 1 | 3 | 2 | 0 | 0 | 1 | 1 | 3 | 1 | 1 | 0 | 1 |
| 2041916 | 1 | 2 | 1 | 2 | 1 | 2 | 2 | 1 | 3 | 4 | 0 | 0 | 1 | 1 | 3 | 1 | 1 | 0 | 1 |
| 2041917 | 1 | 2 | 1 | 2 | 1 | 2 | 1 | 2 | 3 | 4 | 0 | 0 | 1 | 1 | 2 | 1 | 1 | 0 | 1 |
| 2041918 | 1 | 3 | 1 | 2 | 1 | 2 | 2 | 2 | 3 | 3 | 0 | 0 | 1 | 1 | 3 | 1 | 1 | 0 | 1 |
| 2041919 | 1 | 3 | 1 | 2 | 1 | 2 | 1 | 1 | 3 | 4 | 0 | 0 | 1 | 1 | 3 | 1 | 1 | 0 | 1 |
| 2041920 | 0 | 5 | 1 | 1 | 1 | 2 | 1 | 1 | 3 | 4 | 0 | 0 | 1 | 1 | 2 | 1 | 1 | 0 | 1 |
| 2041921 | 0 | 3 | 1 | 2 | 1 | 2 | 2 | 2 | 3 | 4 | 0 | 0 | 1 | 1 | 1 | 1 | 0 | 0 | 0 |
| 2041922 | 1 | 4 | 1 | 2 | 1 | 2 | 1 | 1 | 3 | 3 | 0 | 0 | 1 | 1 | 2 | 1 | 1 | 0 | 1 |
| 2041923 | 1 | 5 | 1 | 1 | 1 | 2 | 1 | 1 | 3 | 4 | 0 | 0 | 1 | 1 | 2 | 6 | 1 | 1 | 1 |
| 2041924 | 1 | 2 | 1 | 2 | 1 | 2 | 2 | 2 | 3 | 4 | 0 | 0 | 1 | 1 | 3 | 1 | 1 | 0 | 1 |
| 2041925 | 1 | 2 | 1 | 2 | 1 | 2 | 1 | 1 | 3 | 4 | 0 | 0 | 1 | 1 | 1 | 1 | 0 | 0 | 0 |
| 2041926 | 0 | 2 | 1 | 3 | 1 | 2 | 1 | 1 | 3 | 4 | 0 | 0 | 1 | 1 | 1 | 1 | 0 | 0 | 0 |
| 2041927 | 1 | 4 | 1 | 1 | 1 | 2 | 2 | 2 | 3 | 4 | 0 | 0 | 1 | 1 | 2 | 1 | 1 | 0 | 1 |
| 2041928 | 0 | 4 | 1 | 1 | 1 | 2 | 1 | 1 | 3 | 4 | 0 | 0 | 1 | 1 | 1 | 1 | 0 | 0 | 0 |
| 2041929 | 0 | 1 | 1 | 4 | 1 | 2 | 1 | 1 | 3 | 4 | 0 | 0 | 1 | 1 | 2 | 1 | 1 | 0 | 1 |
| 2041930 | 1 | 2 | 1 | 2 | 1 | 2 | 1 | 1 | 1 | 4 | 0 | 0 | 1 | 1 | 2 | 1 | 1 | 0 | 1 |
| 204201 | 1 | 2 | 1 | 4 | 1 | 2 | 2 | 1 | 1 | 4 | 0 | 0 | 1 | 1 | 3 | 7 | 1 | 1 | 1 |
| 204202 | 0 | 1 | 1 | 4 | 1 | 2 | 2 | 1 | 1 | 4 | 0 | 0 | 1 | 1 | 3 | 1 | 1 | 0 | 1 |
| 204203 | 1 | 3 | 1 | 5 | 1 | 5 | 2 | 1 | 1 | 4 | 0 | 0 | 1 | 1 | 3 | 1 | 1 | 0 | 1 |
| 204204 | 1 | 2 | 1 | 2 | 1 | 2 | 1 | 1 | 1 | 4 | 0 | 0 | 1 | 1 | 3 | 7 | 1 | 1 | 1 |
| 204205 | 1 | 3 | 1 | 2 | 1 | 2 | 2 | 2 | 1 | 2 | 0 | 0 | 1 | 1 | 2 | 1 | 1 | 0 | 1 |
| 204206 | 1 | 5 | 1 | 1 | 1 | 2 | 1 | 1 | 1 | 2 | 0 | 0 | 0 | 99 | 99 | 99 | 99 | 99 | 99 |
| 204207 | 1 | 3 | 1 | 2 | 1 | 2 | 1 | 2 | 1 | 1 | 0 | 0 | 1 | 1 | 3 | 1 | 1 | 0 | 1 |
| 204208 | 1 | 1 | 1 | 3 | 1 | 2 | 2 | 1 | 1 | 2 | 0 | 0 | 1 | 1 | 3 | 1 | 1 | 0 | 1 |
| 204209 | 0 | 3 | 1 | 2 | 1 | 1 | 1 | 1 | 1 | 2 | 0 | 0 | 1 | 1 | 2 | 1 | 1 | 0 | 1 |
| 2042010 | 0 | 3 | 1 | 2 | 1 | 3 | 1 | 2 | 1 | 2 | 0 | 0 | 1 | 1 | 3 | 1 | 1 | 0 | 1 |
| 2042011 | 1 | 5 | 1 | 1 | 1 | 2 | 1 | 1 | 1 | 3 | 0 | 0 | 1 | 1 | 2 | 1 | 1 | 0 | 1 |
| 2042012 | 0 | 5 | 1 | 1 | 1 | 2 | 1 | 1 | 1 | 3 | 0 | 0 | 0 | 99 | 99 | 99 | 99 | 99 | 99 |
| 2042013 | 1 | 3 | 1 | 2 | 1 | 2 | 2 | 2 | 1 | 4 | 0 | 0 | 1 | 1 | 3 | 1 | 1 | 0 | 1 |
| 2042014 | 0 | 4 | 1 | 1 | 1 | 2 | 1 | 2 | 1 | 4 | 0 | 0 | 1 | 1 | 2 | 1 | 1 | 0 | 1 |
| 2042015 | 1 | 2 | 1 | 2 | 1 | 2 | 2 | 1 | 1 | 2 | 0 | 0 | 1 | 1 | 2 | 1 | 1 | 0 | 1 |
| 2042016 | 0 | 2 | 1 | 2 | 1 | 1 | 1 | 1 | 1 | 2 | 0 | 0 | 1 | 1 | 3 | 6 | 1 | 1 | 1 |
| 2042017 | 0 | 2 | 1 | 2 | 1 | 2 | 3 | 2 | 1 | 1 | 0 | 0 | 1 | 1 | 3 | 6 | 1 | 1 | 1 |
| 2042018 | 1 | 4 | 1 | 2 | 1 | 2 | 2 | 2 | 4 | 4 | 0 | 0 | 1 | 1 | 3 | 1 | 1 | 0 | 1 |
| 2042019 | 1 | 3 | 1 | 2 | 1 | 2 | 2 | 2 | 1 | 4 | 0 | 0 | 1 | 1 | 3 | 1 | 1 | 0 | 1 |
| 2042020 | 0 | 5 | 1 | 1 | 1 | 2 | 1 | 1 | 1 | 4 | 0 | 0 | 1 | 1 | 3 | 1 | 1 | 0 | 1 |

| | | | | | | | | | | | | | | | | | | |
|---|---|---|---|---|---|---|---|---|---|---|---|---|---|---|---|---|---|---|
| 2042021 | 1 | 3 | 1 | 2 | 1 | 2 | 2 | 1 | 1 | 4 | 0 | 0 | 1 | 1 | 4 | 1 | 1 | 0 | 1 |
| 2042022 | 1 | 5 | 1 | 1 | 1 | 2 | 1 | 1 | 1 | 4 | 0 | 0 | 1 | 1 | 2 | 6 | 1 | 1 | 1 |
| 2042023 | 1 | 2 | 1 | 2 | 1 | 2 | 2 | 2 | 1 | 4 | 0 | 0 | 1 | 1 | 1 | 1 | 0 | 0 | 0 |
| 2042024 | 1 | 1 | 1 | 4 | 1 | 2 | 2 | 1 | 1 | 4 | 0 | 0 | 1 | 1 | 4 | 1 | 1 | 0 | 1 |
| 2042025 | 1 | 3 | 1 | 2 | 1 | 2 | 1 | 2 | 1 | 4 | 0 | 0 | 1 | 1 | 3 | 1 | 1 | 0 | 1 |
| 205211 | 0 | 3 | 1 | 2 | 1 | 2 | 1 | 2 | 3 | 1 | 0 | 0 | 1 | 1 | 4 | 4 | 1 | 1 | 1 |
| 205212 | 1 | 3 | 1 | 2 | 1 | 2 | 3 | 2 | 3 | 1 | 0 | 0 | 1 | 1 | 1 | 1 | 0 | 0 | 0 |
| 205213 | 0 | 5 | 1 | 2 | 1 | 2 | 1 | 2 | 3 | 1 | 0 | 0 | 1 | 1 | 2 | 1 | 1 | 0 | 1 |
| 205214 | 0 | 3 | 1 | 2 | 1 | 2 | 1 | 1 | 3 | 2 | 0 | 0 | 1 | 1 | 2 | 1 | 1 | 0 | 1 |
| 205215 | 1 | 4 | 1 | 4 | 1 | 2 | 1 | 1 | 3 | 3 | 0 | 0 | 1 | 1 | 2 | 1 | 1 | 0 | 1 |
| 205216 | 0 | 4 | 1 | 1 | 1 | 2 | 1 | 1 | 3 | 3 | 0 | 0 | 1 | 1 | 3 | 1 | 1 | 0 | 1 |
| 205217 | 0 | 2 | 1 | 2 | 1 | 2 | 1 | 2 | 3 | 3 | 0 | 0 | 1 | 1 | 2 | 1 | 1 | 0 | 1 |
| 205218 | 1 | 2 | 1 | 4 | 1 | 2 | 1 | 2 | 3 | 3 | 0 | 0 | 1 | 1 | 1 | 1 | 0 | 0 | 0 |
| 205219 | 1 | 5 | 1 | 2 | 1 | 2 | 1 | 2 | 3 | 4 | 0 | 0 | 1 | 1 | 1 | 1 | 0 | 0 | 0 |
| 2052110 | 0 | 3 | 1 | 2 | 1 | 2 | 2 | 2 | 3 | 2 | 0 | 0 | 1 | 1 | 1 | 1 | 0 | 0 | 0 |
| 2052111 | 0 | 4 | 1 | 2 | 1 | 2 | 1 | 1 | 3 | 1 | 0 | 0 | 1 | 1 | 2 | 1 | 1 | 0 | 1 |
| 2052112 | 1 | 3 | 1 | 2 | 1 | 2 | 2 | 1 | 3 | 3 | 0 | 0 | 1 | 1 | 2 | 1 | 1 | 0 | 1 |
| 2052113 | 0 | 3 | 1 | 1 | 1 | 2 | 2 | 1 | 3 | 2 | 0 | 0 | 1 | 1 | 2 | 1 | 1 | 0 | 1 |
| 2052114 | 0 | 2 | 1 | 2 | 1 | 2 | 1 | 2 | 3 | 2 | 0 | 0 | 1 | 1 | 1 | 1 | 0 | 0 | 0 |
| 2052115 | 0 | 1 | 1 | 5 | 1 | 5 | 2 | 1 | 3 | 1 | 0 | 0 | 1 | 1 | 1 | 1 | 0 | 0 | 0 |
| 2052116 | 1 | 3 | 1 | 2 | 1 | 2 | 1 | 2 | 3 | 4 | 0 | 0 | 1 | 1 | 3 | 1 | 1 | 0 | 1 |
| 2052117 | 1 | 2 | 1 | 2 | 1 | 2 | 2 | 1 | 3 | 1 | 0 | 0 | 1 | 1 | 1 | 1 | 0 | 0 | 0 |
| 2052118 | 0 | 3 | 1 | 2 | 1 | 2 | 1 | 2 | 3 | 1 | 0 | 0 | 1 | 1 | 5 | 4 | 1 | 1 | 1 |
| 2052119 | 0 | 3 | 1 | 2 | 1 | 2 | 1 | 2 | 3 | 1 | 0 | 0 | 1 | 1 | 1 | 1 | 0 | 0 | 0 |
| 2052120 | 1 | 5 | 1 | 1 | 1 | 2 | 1 | 1 | 3 | 1 | 0 | 0 | 1 | 1 | 2 | 1 | 1 | 0 | 1 |
| 2052121 | 0 | 3 | 1 | 2 | 1 | 2 | 2 | 2 | 3 | 1 | 0 | 0 | 1 | 1 | 3 | 1 | 1 | 0 | 1 |
| 2052122 | 1 | 3 | 1 | 2 | 1 | 2 | 1 | 2 | 1 | 1 | 0 | 0 | 0 | 99 | 99 | 99 | 99 | 99 | 99 |
| 2052123 | 0 | 3 | 1 | 2 | 1 | 2 | 2 | 2 | 3 | 1 | 0 | 0 | 1 | 1 | 1 | 1 | 0 | 0 | 0 |
| 2052124 | 0 | 3 | 1 | 2 | 1 | 2 | 1 | 2 | 3 | 1 | 0 | 0 | 1 | 1 | 2 | 1 | 1 | 0 | 1 |
| 2052125 | 1 | 2 | 1 | 2 | 1 | 2 | 1 | 2 | 1 | 4 | 0 | 0 | 1 | 1 | 2 | 1 | 1 | 0 | 1 |
| 2052126 | 1 | 2 | 1 | 4 | 1 | 2 | 2 | 2 | 3 | 2 | 0 | 0 | 1 | 1 | 1 | 1 | 0 | 0 | 0 |
| 2052127 | 0 | 3 | 1 | 2 | 1 | 2 | 1 | 2 | 3 | 2 | 0 | 0 | 1 | 1 | 2 | 1 | 1 | 0 | 1 |
| 2052128 | 0 | 3 | 1 | 3 | 2 | 2 | 2 | 2 | 3 | 2 | 0 | 0 | 1 | 1 | 1 | 1 | 0 | 0 | 0 |
| 2052129 | 1 | 4 | 1 | 2 | 1 | 2 | 2 | 1 | 2 | 1 | 0 | 0 | 1 | 1 | 3 | 1 | 1 | 0 | 1 |
| 2052130 | 0 | 5 | 1 | 1 | 1 | 2 | 1 | 1 | 3 | 3 | 0 | 0 | 0 | 99 | 99 | 99 | 99 | 99 | 99 |
| 205221 | 0 | 2 | 1 | 1 | 1 | 2 | 2 | 2 | 4 | 4 | 0 | 0 | 1 | 1 | 1 | 1 | 0 | 0 | 0 |
| 205222 | 1 | 3 | 1 | 1 | 1 | 2 | 2 | 2 | 4 | 4 | 0 | 0 | 1 | 1 | 2 | 1 | 1 | 0 | 1 |
| 205223 | 1 | 1 | 1 | 1 | 1 | 2 | 1 | 2 | 4 | 4 | 0 | 0 | 1 | 1 | 3 | 1 | 1 | 0 | 1 |
| 205224 | 0 | 1 | 1 | 1 | 2 | 3 | 2 | 1 | 4 | 4 | 0 | 0 | 1 | 1 | 2 | 7 | 1 | 1 | 1 |
| 205225 | 1 | 2 | 1 | 3 | 1 | 2 | 1 | 2 | 4 | 4 | 0 | 0 | 1 | 1 | 3 | 1 | 1 | 0 | 1 |
| 205226 | 1 | 3 | 1 | 2 | 1 | 2 | 1 | 1 | 4 | 4 | 0 | 0 | 1 | 1 | 3 | 1 | 1 | 0 | 1 |
| 205227 | 0 | 4 | 1 | 1 | 1 | 2 | 2 | 2 | 4 | 4 | 0 | 0 | 1 | 1 | 3 | 1 | 1 | 0 | 1 |
| 205228 | 0 | 4 | 1 | 1 | 1 | 2 | 1 | 2 | 3 | 4 | 0 | 0 | 1 | 1 | 2 | 1 | 1 | 0 | 1 |
| 205229 | 0 | 1 | 1 | 4 | 1 | 2 | 2 | 2 | 3 | 2 | 0 | 0 | 1 | 1 | 2 | 1 | 1 | 0 | 1 |
| 2052210 | 1 | 5 | 1 | 1 | 1 | 4 | 2 | 2 | 3 | 1 | 0 | 0 | 1 | 1 | 2 | 1 | 1 | 0 | 1 |
| 2052211 | 0 | 3 | 1 | 2 | 1 | 2 | 2 | 2 | 3 | 1 | 0 | 0 | 1 | 1 | 1 | 1 | 0 | 0 | 0 |
| 2052212 | 0 | 1 | 1 | 4 | 1 | 2 | 2 | 1 | 3 | 1 | 0 | 0 | 1 | 1 | 1 | 1 | 0 | 0 | 0 |
| 2052213 | 0 | 4 | 1 | 3 | 2 | 1 | 2 | 2 | 3 | 2 | 0 | 0 | 1 | 1 | 2 | 1 | 1 | 0 | 1 |
| 2052214 | 0 | 4 | 1 | 2 | 2 | 1 | 2 | 2 | 3 | 2 | 0 | 0 | 1 | 1 | 2 | 1 | 1 | 0 | 1 |
| 2052215 | 0 | 3 | 1 | 3 | 1 | 2 | 2 | 2 | 3 | 1 | 0 | 0 | 1 | 1 | 1 | 1 | 0 | 0 | 0 |
| 2052216 | 0 | 2 | 1 | 1 | 1 | 2 | 1 | 2 | 3 | 2 | 0 | 0 | 1 | 1 | 1 | 2 | 0 | 0 | 0 |
| 2052217 | 0 | 2 | 1 | 2 | 1 | 2 | 1 | 2 | 3 | 3 | 0 | 0 | 1 | 1 | 1 | 1 | 0 | 0 | 0 |
| 2052218 | 0 | 1 | 1 | 2 | 1 | 2 | 1 | 1 | 3 | 4 | 0 | 0 | 1 | 1 | 2 | 1 | 1 | 0 | 1 |
| 2052219 | 0 | 5 | 1 | 1 | 1 | 2 | 1 | 2 | 3 | 4 | 0 | 0 | 1 | 1 | 2 | 1 | 1 | 0 | 1 |
| 2052220 | 0 | 2 | 1 | 2 | 1 | 2 | 2 | 2 | 3 | 4 | 0 | 0 | 1 | 1 | 3 | 1 | 1 | 0 | 1 |
| 2052221 | 0 | 3 | 1 | 2 | 1 | 2 | 2 | 2 | 3 | 4 | 0 | 0 | 1 | 1 | 3 | 1 | 1 | 0 | 1 |

| | | | | | | | | | | | | | | | | | | | |
|---|---|---|---|---|---|---|---|---|---|---|---|---|---|---|---|---|---|---|---|
| 2052222 | 0 | 2 | 1 | 1 | 1 | 3 | 3 | 2 | 2 | 4 | 0 | 0 | 1 | 1 | 2 | 1 | 1 | 0 | 1 |
| 2052223 | 0 | 2 | 1 | 1 | 1 | 2 | 1 | 2 | 3 | 4 | 0 | 0 | 1 | 1 | 2 | 1 | 1 | 0 | 1 |
| 2052224 | 1 | 5 | 1 | 2 | 1 | 2 | 2 | 1 | 3 | 4 | 0 | 0 | 1 | 1 | 2 | 1 | 1 | 0 | 1 |
| 2052225 | 0 | 3 | 1 | 1 | 1 | 2 | 2 | 1 | 3 | 1 | 0 | 0 | 1 | 1 | 2 | 1 | 1 | 0 | 1 |
| 2052226 | 0 | 4 | 1 | 1 | 1 | 1 | 2 | 2 | 3 | 1 | 0 | 0 | 1 | 1 | 1 | 1 | 0 | 0 | 0 |
| 2052227 | 0 | 2 | 1 | 1 | 1 | 1 | 1 | 2 | 3 | 1 | 0 | 0 | 1 | 1 | 1 | 1 | 0 | 0 | 0 |
| 2052228 | 0 | 1 | 1 | 1 | 1 | 2 | 1 | 2 | 4 | 2 | 0 | 0 | 1 | 1 | 2 | 1 | 1 | 0 | 1 |
| 2052229 | 0 | 5 | 1 | 3 | 2 | 2 | 2 | 1 | 4 | 2 | 0 | 0 | 1 | 1 | 1 | 1 | 0 | 0 | 0 |
| 2052230 | 0 | 2 | 1 | 1 | 1 | 2 | 1 | 2 | 3 | 1 | 0 | 0 | 1 | 1 | 2 | 1 | 1 | 0 | 1 |
| 205231 | 0 | 1 | 1 | 5 | 1 | 5 | 2 | 1 | 3 | 1 | 0 | 0 | 1 | 1 | 2 | 1 | 1 | 0 | 1 |
| 205232 | 0 | 3 | 1 | 1 | 1 | 1 | 2 | 2 | 3 | 1 | 0 | 0 | 1 | 1 | 3 | 1 | 1 | 0 | 1 |
| 205233 | 0 | 1 | 1 | 4 | 1 | 2 | 2 | 2 | 3 | 1 | 0 | 0 | 1 | 1 | 1 | 1 | 0 | 0 | 0 |
| 205234 | 0 | 3 | 1 | 4 | 1 | 2 | 2 | 1 | 3 | 1 | 0 | 0 | 1 | 1 | 2 | 1 | 1 | 0 | 1 |
| 205235 | 1 | 2 | 1 | 3 | 1 | 3 | 3 | 1 | 3 | 1 | 1 | 0 | 1 | 1 | 4 | 1 | 1 | 0 | 1 |
| 205236 | 0 | 3 | 1 | 2 | 1 | 2 | 2 | 1 | 3 | 1 | 0 | 0 | 1 | 1 | 2 | 1 | 1 | 0 | 1 |
| 205237 | 0 | 2 | 1 | 2 | 1 | 3 | 2 | 2 | 4 | 2 | 0 | 0 | 1 | 1 | 1 | 1 | 0 | 0 | 0 |
| 205238 | 1 | 3 | 1 | 1 | 1 | 2 | 2 | 2 | 4 | 3 | 0 | 0 | 1 | 1 | 2 | 1 | 1 | 0 | 1 |
| 205239 | 0 | 1 | 1 | 3 | 2 | 1 | 1 | 2 | 4 | 4 | 0 | 0 | 1 | 1 | 3 | 1 | 1 | 0 | 1 |
| 2052310 | 1 | 3 | 1 | 1 | 1 | 2 | 2 | 2 | 4 | 4 | 0 | 0 | 1 | 1 | 3 | 1 | 1 | 0 | 1 |
| 2052311 | 1 | 2 | 1 | 1 | 1 | 4 | 1 | 2 | 4 | 2 | 0 | 0 | 1 | 1 | 2 | 1 | 1 | 0 | 1 |
| 2052312 | 1 | 3 | 1 | 1 | 1 | 4 | 2 | 2 | 4 | 3 | 0 | 0 | 1 | 1 | 2 | 1 | 1 | 0 | 1 |
| 2052313 | 0 | 3 | 1 | 2 | 1 | 2 | 2 | 1 | 4 | 4 | 0 | 0 | 1 | 1 | 2 | 1 | 1 | 0 | 1 |
| 2052314 | 0 | 3 | 1 | 5 | 1 | 5 | 2 | 2 | 4 | 4 | 0 | 0 | 1 | 1 | 1 | 1 | 0 | 0 | 0 |
| 2052315 | 0 | 2 | 1 | 1 | 1 | 2 | 2 | 2 | 4 | 2 | 0 | 0 | 1 | 1 | 2 | 1 | 1 | 0 | 1 |
| 2052316 | 1 | 5 | 1 | 1 | 1 | 2 | 2 | 2 | 4 | 3 | 0 | 0 | 1 | 1 | 3 | 1 | 1 | 0 | 1 |
| 2052317 | 0 | 1 | 1 | 1 | 1 | 2 | 2 | 1 | 4 | 4 | 0 | 0 | 1 | 1 | 3 | 1 | 1 | 0 | 1 |
| 2052318 | 1 | 5 | 1 | 4 | 1 | 5 | 3 | 2 | 4 | 4 | 1 | 1 | 1 | 1 | 1 | 1 | 0 | 0 | 0 |
| 2052319 | 1 | 5 | 1 | 5 | 1 | 5 | 3 | 2 | 4 | 1 | 0 | 0 | 1 | 1 | 1 | 1 | 0 | 0 | 0 |
| 2052320 | 0 | 2 | 1 | 2 | 1 | 3 | 3 | 2 | 3 | 1 | 1 | 1 | 1 | 1 | 2 | 1 | 1 | 0 | 1 |
| 2052321 | 1 | 5 | 1 | 4 | 1 | 2 | 2 | 2 | 3 | 1 | 0 | 0 | 1 | 1 | 2 | 1 | 1 | 0 | 1 |
| 2052322 | 0 | 4 | 1 | 1 | 1 | 2 | 2 | 2 | 3 | 1 | 0 | 0 | 1 | 1 | 3 | 1 | 1 | 0 | 1 |
| 2052323 | 1 | 5 | 1 | 1 | 1 | 2 | 2 | 2 | 3 | 1 | 0 | 0 | 1 | 1 | 4 | 1 | 1 | 0 | 1 |
| 2052324 | 0 | 1 | 1 | 1 | 1 | 2 | 1 | 2 | 3 | 1 | 0 | 0 | 1 | 1 | 2 | 1 | 1 | 0 | 1 |
| 2052325 | 1 | 3 | 1 | 1 | 1 | 2 | 2 | 2 | 3 | 1 | 0 | 0 | 1 | 1 | 3 | 1 | 1 | 0 | 1 |
| 2052326 | 0 | 1 | 1 | 2 | 1 | 2 | 2 | 2 | 3 | 1 | 0 | 0 | 1 | 1 | 3 | 1 | 1 | 0 | 1 |
| 2052327 | 1 | 5 | 1 | 1 | 1 | 2 | 1 | 1 | 3 | 1 | 0 | 0 | 1 | 1 | 2 | 1 | 1 | 0 | 1 |
| 2052328 | 1 | 3 | 1 | 2 | 1 | 2 | 2 | 2 | 3 | 1 | 0 | 0 | 1 | 1 | 1 | 1 | 0 | 0 | 0 |
| 2052329 | 1 | 1 | 1 | 1 | 1 | 2 | 2 | 1 | 4 | 1 | 0 | 0 | 1 | 1 | 2 | 6 | 1 | 1 | 1 |
| 2052330 | 0 | 1 | 1 | 1 | 1 | 1 | 1 | 2 | 4 | 1 | 0 | 0 | 1 | 1 | 1 | 1 | 0 | 0 | 0 |
| 205241 | 0 | 1 | 1 | 4 | 2 | 1 | 2 | 1 | 4 | 4 | 0 | 0 | 1 | 1 | 1 | 1 | 0 | 0 | 0 |
| 205242 | 0 | 3 | 1 | 2 | 1 | 2 | 3 | 2 | 4 | 3 | 0 | 0 | 1 | 1 | 1 | 6 | 0 | 1 | 1 |
| 205243 | 0 | 3 | 1 | 4 | 2 | 1 | 3 | 1 | 4 | 4 | 1 | 1 | 1 | 1 | 1 | 1 | 0 | 0 | 0 |
| 205244 | 0 | 3 | 1 | 3 | 2 | 1 | 3 | 2 | 4 | 3 | 1 | 1 | 1 | 1 | 1 | 1 | 0 | 0 | 0 |
| 205245 | 1 | 3 | 1 | 4 | 2 | 2 | 2 | 2 | 4 | 4 | 1 | 1 | 1 | 1 | 1 | 1 | 0 | 0 | 0 |
| 205246 | 0 | 4 | 1 | 5 | 2 | 5 | 3 | 2 | 4 | 4 | 1 | 1 | 1 | 1 | 1 | 1 | 0 | 0 | 0 |
| 205247 | 1 | 4 | 1 | 3 | 1 | 2 | 3 | 2 | 4 | 4 | 0 | 0 | 1 | 1 | 1 | 1 | 0 | 0 | 0 |
| 205248 | 1 | 3 | 1 | 2 | 1 | 4 | 2 | 2 | 4 | 4 | 0 | 0 | 1 | 1 | 2 | 5 | 1 | 1 | 1 |
| 205249 | 0 | 2 | 1 | 2 | 2 | 1 | 2 | 2 | 4 | 4 | 0 | 0 | 1 | 1 | 3 | 1 | 1 | 0 | 1 |
| 2052410 | 0 | 2 | 1 | 2 | 1 | 1 | 3 | 2 | 4 | 4 | 0 | 0 | 1 | 1 | 3 | 1 | 1 | 0 | 1 |
| 2052411 | 1 | 2 | 1 | 1 | 1 | 4 | 3 | 1 | 4 | 4 | 1 | 0 | 1 | 1 | 1 | 1 | 0 | 0 | 0 |
| 2052412 | 0 | 4 | 1 | 3 | 2 | 3 | 3 | 1 | 4 | 4 | 1 | 1 | 1 | 1 | 1 | 1 | 0 | 0 | 0 |
| 2052413 | 1 | 5 | 1 | 3 | 1 | 4 | 2 | 2 | 4 | 4 | 0 | 0 | 1 | 1 | 1 | 1 | 0 | 0 | 0 |
| 2052414 | 1 | 4 | 1 | 2 | 2 | 2 | 3 | 2 | 4 | 4 | 0 | 0 | 1 | 1 | 3 | 1 | 1 | 0 | 1 |
| 2052415 | 0 | 3 | 1 | 4 | 1 | 5 | 3 | 2 | 4 | 2 | 1 | 1 | 1 | 1 | 1 | 1 | 0 | 0 | 0 |
| 2052416 | 1 | 4 | 1 | 1 | 1 | 2 | 2 | 2 | 4 | 3 | 0 | 0 | 1 | 1 | 1 | 6 | 0 | 1 | 1 |
| 2052417 | 0 | 3 | 1 | 5 | 2 | 5 | 3 | 1 | 4 | 3 | 0 | 0 | 1 | 1 | 1 | 1 | 0 | 0 | 0 |

| | | | | | | | | | | | | | | | | | | | |
|---|---|---|---|---|---|---|---|---|---|---|---|---|---|---|---|---|---|---|---|
| 2052418 | 1 | 3 | 1 | 1 | 1 | 5 | 2 | 2 | 4 | 4 | 0 | 0 | 1 | 1 | 3 | 1 | 1 | 0 | 1 |
| 2052419 | 0 | 1 | 1 | 1 | 1 | 1 | 2 | 2 | 4 | 2 | 0 | 0 | 1 | 1 | 2 | 1 | 1 | 0 | 1 |
| 2052420 | 0 | 2 | 1 | 1 | 1 | 1 | 3 | 2 | 4 | 3 | 1 | 1 | 1 | 1 | 3 | 1 | 1 | 0 | 1 |
| 2052421 | 1 | 5 | 1 | 1 | 1 | 2 | 2 | 2 | 4 | 4 | 0 | 0 | 1 | 1 | 2 | 1 | 1 | 0 | 1 |
| 2052422 | 1 | 1 | 1 | 3 | 2 | 5 | 3 | 2 | 4 | 4 | 0 | 0 | 1 | 1 | 1 | 1 | 0 | 0 | 0 |
| 2052423 | 0 | 1 | 1 | 4 | 2 | 1 | 2 | 1 | 4 | 4 | 0 | 0 | 1 | 1 | 3 | 6 | 1 | 1 | 1 |
| 2052424 | 0 | 2 | 1 | 2 | 2 | 1 | 2 | 1 | 4 | 4 | 0 | 0 | 1 | 1 | 1 | 1 | 0 | 0 | 0 |
| 2052425 | 1 | 3 | 1 | 2 | 1 | 4 | 3 | 2 | 4 | 4 | 0 | 0 | 1 | 1 | 2 | 1 | 1 | 0 | 1 |
| 2052426 | 1 | 3 | 1 | 2 | 1 | 4 | 3 | 2 | 4 | 4 | 0 | 0 | 1 | 1 | 1 | 1 | 0 | 0 | 0 |
| 2052427 | 0 | 2 | 1 | 3 | 1 | 1 | 3 | 1 | 4 | 4 | 1 | 1 | 1 | 1 | 2 | 1 | 1 | 0 | 1 |
| 2052428 | 1 | 3 | 1 | 4 | 2 | 4 | 3 | 1 | 4 | 4 | 1 | 1 | 1 | 1 | 3 | 1 | 1 | 0 | 1 |
| 2052429 | 0 | 4 | 1 | 1 | 1 | 1 | 3 | 2 | 4 | 2 | 0 | 0 | 1 | 1 | 3 | 1 | 1 | 0 | 1 |
| 2052430 | 0 | 5 | 1 | 1 | 2 | 3 | 2 | 1 | 4 | 4 | 0 | 0 | 1 | 1 | 4 | 1 | 1 | 0 | 1 |
| 205251 | 0 | 2 | 1 | 2 | 1 | 2 | 2 | 1 | 2 | 1 | 0 | 0 | 1 | 1 | 1 | 1 | 0 | 0 | 0 |
| 205252 | 0 | 3 | 1 | 1 | 1 | 2 | 1 | 1 | 2 | 1 | 0 | 0 | 1 | 1 | 3 | 1 | 1 | 0 | 1 |
| 205253 | 0 | 2 | 1 | 4 | 1 | 1 | 2 | 2 | 2 | 1 | 0 | 0 | 1 | 1 | 2 | 1 | 1 | 0 | 1 |
| 205254 | 1 | 5 | 1 | 2 | 1 | 2 | 2 | 1 | 2 | 1 | 0 | 0 | 1 | 1 | 3 | 1 | 1 | 0 | 1 |
| 205255 | 0 | 2 | 1 | 1 | 1 | 3 | 1 | 2 | 2 | 1 | 0 | 0 | 1 | 1 | 2 | 1 | 1 | 0 | 1 |
| 205256 | 0 | 2 | 1 | 2 | 1 | 1 | 2 | 2 | 2 | 1 | 0 | 0 | 1 | 1 | 2 | 1 | 1 | 0 | 1 |
| 205257 | 0 | 2 | 1 | 2 | 1 | 2 | 2 | 2 | 2 | 1 | 0 | 0 | 1 | 1 | 3 | 1 | 1 | 0 | 1 |
| 205258 | 1 | 2 | 1 | 1 | 1 | 2 | 1 | 1 | 2 | 1 | 0 | 0 | 1 | 1 | 3 | 1 | 1 | 0 | 1 |
| 205259 | 0 | 3 | 1 | 1 | 2 | 2 | 1 | 2 | 2 | 1 | 0 | 0 | 1 | 1 | 1 | 1 | 0 | 0 | 0 |
| 2052510 | 1 | 5 | 1 | 1 | 1 | 2 | 1 | 1 | 2 | 1 | 0 | 0 | 1 | 1 | 3 | 1 | 1 | 0 | 1 |
| 2052511 | 1 | 2 | 1 | 2 | 1 | 2 | 1 | 2 | 2 | 1 | 0 | 0 | 1 | 1 | 3 | 1 | 1 | 0 | 1 |
| 2052512 | 0 | 1 | 1 | 2 | 1 | 2 | 1 | 2 | 2 | 1 | 0 | 0 | 1 | 1 | 2 | 1 | 1 | 0 | 1 |
| 2052513 | 0 | 2 | 1 | 1 | 1 | 2 | 1 | 2 | 2 | 1 | 0 | 0 | 1 | 1 | 2 | 1 | 1 | 0 | 1 |
| 2052514 | 0 | 2 | 1 | 4 | 1 | 2 | 2 | 2 | 2 | 1 | 0 | 0 | 1 | 1 | 2 | 1 | 1 | 0 | 1 |
| 2052515 | 0 | 2 | 1 | 2 | 1 | 2 | 2 | 2 | 2 | 1 | 1 | 0 | 1 | 1 | 3 | 1 | 1 | 0 | 1 |
| 2052516 | 0 | 2 | 1 | 4 | 1 | 3 | 2 | 2 | 2 | 1 | 0 | 0 | 1 | 1 | 1 | 1 | 0 | 0 | 0 |
| 2052517 | 0 | 2 | 1 | 1 | 1 | 2 | 1 | 1 | 2 | 2 | 0 | 0 | 1 | 1 | 2 | 1 | 1 | 0 | 1 |
| 2052518 | 0 | 2 | 1 | 1 | 1 | 2 | 1 | 2 | 2 | 2 | 0 | 0 | 1 | 1 | 2 | 1 | 1 | 0 | 1 |
| 2052519 | 1 | 2 | 1 | 1 | 1 | 2 | 1 | 2 | 2 | 1 | 0 | 0 | 1 | 1 | 3 | 1 | 1 | 0 | 1 |
| 2052520 | 0 | 1 | 1 | 1 | 1 | 2 | 2 | 2 | 2 | 1 | 0 | 0 | 1 | 1 | 2 | 1 | 1 | 0 | 1 |
| 2052521 | 0 | 3 | 1 | 1 | 1 | 2 | 1 | 2 | 3 | 1 | 0 | 0 | 1 | 1 | 2 | 1 | 1 | 0 | 1 |
| 2052522 | 0 | 2 | 1 | 1 | 1 | 2 | 2 | 2 | 2 | 1 | 0 | 0 | 1 | 1 | 2 | 1 | 1 | 0 | 1 |
| 2052523 | 0 | 3 | 1 | 1 | 1 | 2 | 2 | 2 | 3 | 1 | 0 | 0 | 1 | 1 | 3 | 1 | 1 | 0 | 1 |
| 2052524 | 0 | 3 | 1 | 1 | 1 | 2 | 2 | 2 | 3 | 1 | 0 | 0 | 1 | 1 | 2 | 1 | 1 | 0 | 1 |
| 2052525 | 0 | 3 | 1 | 2 | 1 | 3 | 3 | 2 | 3 | 2 | 0 | 0 | 1 | 1 | 2 | 1 | 1 | 0 | 1 |
| 2052526 | 0 | 2 | 1 | 2 | 1 | 2 | 1 | 2 | 3 | 2 | 0 | 0 | 1 | 1 | 2 | 1 | 1 | 0 | 1 |
| 2052527 | 0 | 1 | 1 | 1 | 1 | 1 | 2 | 1 | 3 | 3 | 0 | 0 | 1 | 1 | 2 | 1 | 1 | 0 | 1 |
| 2052528 | 1 | 3 | 1 | 1 | 1 | 2 | 2 | 2 | 3 | 3 | 0 | 0 | 1 | 1 | 2 | 1 | 1 | 0 | 1 |
| 2052529 | 0 | 3 | 1 | 1 | 1 | 2 | 1 | 2 | 2 | 1 | 0 | 0 | 1 | 1 | 1 | 1 | 0 | 0 | 0 |
| 2052530 | 1 | 3 | 1 | 3 | 1 | 3 | 2 | 2 | 2 | 2 | 0 | 0 | 1 | 1 | 2 | 1 | 1 | 0 | 1 |
| 206261 | 1 | 3 | 1 | 2 | 1 | 2 | 2 | 1 | 3 | 2 | 0 | 0 | 1 | 1 | 1 | 1 | 0 | 0 | 0 |
| 206262 | 1 | 1 | 1 | 2 | 1 | 2 | 1 | 1 | 3 | 2 | 0 | 0 | 1 | 1 | 3 | 1 | 1 | 0 | 1 |
| 206263 | 1 | 3 | 1 | 3 | 1 | 2 | 2 | 2 | 3 | 1 | 0 | 0 | 1 | 1 | 1 | 1 | 0 | 0 | 0 |
| 206264 | 1 | 2 | 1 | 4 | 1 | 2 | 2 | 2 | 3 | 1 | 0 | 0 | 1 | 1 | 1 | 1 | 0 | 0 | 0 |
| 206265 | 1 | 4 | 1 | 2 | 1 | 2 | 2 | 2 | 3 | 1 | 0 | 0 | 1 | 1 | 1 | 1 | 0 | 0 | 0 |
| 206266 | 0 | 1 | 1 | 2 | 1 | 2 | 1 | 1 | 3 | 3 | 0 | 0 | 1 | 1 | 3 | 1 | 1 | 0 | 1 |
| 206267 | 1 | 2 | 1 | 3 | 1 | 2 | 2 | 1 | 3 | 2 | 0 | 0 | 1 | 1 | 1 | 1 | 0 | 0 | 0 |
| 206268 | 0 | 1 | 1 | 4 | 1 | 1 | 2 | 2 | 3 | 3 | 0 | 0 | 1 | 1 | 1 | 1 | 0 | 0 | 0 |
| 206269 | 0 | 4 | 1 | 2 | 1 | 2 | 2 | 2 | 3 | 4 | 0 | 0 | 1 | 1 | 1 | 1 | 0 | 0 | 0 |
| 2062610 | 0 | 1 | 1 | 2 | 1 | 2 | 2 | 2 | 3 | 4 | 0 | 0 | 1 | 1 | 3 | 1 | 1 | 0 | 1 |
| 2062611 | 0 | 2 | 1 | 1 | 1 | 2 | 1 | 1 | 3 | 4 | 0 | 0 | 1 | 1 | 1 | 1 | 0 | 0 | 0 |
| 2062612 | 0 | 1 | 1 | 4 | 1 | 2 | 1 | 2 | 3 | 4 | 0 | 0 | 1 | 1 | 1 | 1 | 0 | 0 | 0 |
| 2062613 | 0 | 3 | 1 | 1 | 1 | 2 | 2 | 2 | 3 | 4 | 0 | 0 | 1 | 1 | 3 | 1 | 1 | 0 | 1 |

| | | | | | | | | | | | | | | | | | | | |
|---|---|---|---|---|---|---|---|---|---|---|---|---|---|---|---|---|---|---|---|
| 2062614 | 0 | 3 | 1 | 2 | 1 | 2 | 2 | 1 | 3 | 4 | 1 | 1 | 1 | 1 | 1 | 1 | 0 | 0 | 0 |
| 2062615 | 1 | 4 | 1 | 1 | 1 | 2 | 1 | 1 | 3 | 3 | 0 | 0 | 1 | 1 | 3 | 1 | 1 | 0 | 1 |
| 2062616 | 1 | 3 | 1 | 1 | 1 | 2 | 1 | 1 | 3 | 4 | 0 | 0 | 1 | 1 | 3 | 1 | 1 | 0 | 1 |
| 2062617 | 0 | 1 | 1 | 3 | 1 | 5 | 1 | 2 | 3 | 3 | 0 | 0 | 1 | 1 | 2 | 1 | 1 | 0 | 1 |
| 2062618 | 1 | 5 | 1 | 1 | 1 | 2 | 2 | 2 | 3 | 3 | 0 | 0 | 1 | 1 | 2 | 1 | 1 | 0 | 1 |
| 2062619 | 0 | 2 | 1 | 2 | 1 | 2 | 2 | 2 | 3 | 4 | 0 | 0 | 1 | 1 | 3 | 1 | 1 | 0 | 1 |
| 2062620 | 0 | 2 | 1 | 1 | 1 | 2 | 2 | 2 | 3 | 2 | 0 | 0 | 1 | 1 | 2 | 1 | 1 | 0 | 1 |
| 2062621 | 0 | 2 | 1 | 1 | 1 | 1 | 1 | 2 | 3 | 2 | 0 | 0 | 1 | 1 | 1 | 1 | 0 | 0 | 0 |
| 2062622 | 0 | 2 | 1 | 1 | 1 | 3 | 3 | 2 | 3 | 1 | 0 | 0 | 1 | 1 | 1 | 1 | 0 | 0 | 0 |
| 2062623 | 0 | 4 | 1 | 1 | 1 | 2 | 2 | 1 | 3 | 2 | 0 | 0 | 1 | 1 | 3 | 1 | 1 | 0 | 1 |
| 2062624 | 1 | 3 | 1 | 2 | 1 | 2 | 2 | 2 | 3 | 1 | 0 | 0 | 1 | 1 | 1 | 1 | 0 | 0 | 0 |
| 2062625 | 1 | 3 | 1 | 3 | 1 | 2 | 2 | 1 | 3 | 2 | 0 | 0 | 1 | 1 | 3 | 1 | 1 | 0 | 1 |
| 206271 | 0 | 4 | 1 | 1 | 1 | 2 | 1 | 2 | 3 | 1 | 0 | 0 | 1 | 1 | 2 | 1 | 1 | 0 | 1 |
| 206272 | 0 | 1 | 1 | 4 | 1 | 2 | 3 | 1 | 3 | 1 | 0 | 0 | 1 | 1 | 2 | 1 | 1 | 0 | 1 |
| 206273 | 0 | 2 | 1 | 2 | 1 | 2 | 2 | 2 | 3 | 1 | 0 | 0 | 1 | 1 | 2 | 1 | 1 | 0 | 1 |
| 206274 | 0 | 1 | 1 | 2 | 1 | 1 | 3 | 1 | 3 | 1 | 0 | 0 | 1 | 1 | 2 | 1 | 1 | 0 | 1 |
| 206275 | 1 | 1 | 1 | 1 | 1 | 2 | 1 | 2 | 3 | 1 | 0 | 0 | 1 | 1 | 2 | 1 | 1 | 0 | 1 |
| 206276 | 0 | 3 | 1 | 2 | 1 | 2 | 2 | 2 | 3 | 4 | 0 | 0 | 1 | 1 | 2 | 1 | 1 | 0 | 1 |
| 206277 | 1 | 1 | 1 | 4 | 1 | 2 | 2 | 1 | 3 | 4 | 0 | 0 | 1 | 1 | 1 | 1 | 0 | 0 | 0 |
| 206278 | 0 | 2 | 1 | 4 | 1 | 2 | 2 | 1 | 3 | 4 | 0 | 0 | 1 | 1 | 2 | 1 | 1 | 0 | 1 |
| 206279 | 1 | 3 | 1 | 2 | 1 | 2 | 1 | 2 | 3 | 4 | 0 | 0 | 1 | 1 | 1 | 1 | 0 | 0 | 0 |
| 2062710 | 0 | 2 | 1 | 2 | 1 | 2 | 1 | 1 | 3 | 4 | 0 | 0 | 1 | 1 | 1 | 1 | 0 | 0 | 0 |
| 2062711 | 0 | 3 | 1 | 1 | 1 | 2 | 1 | 2 | 1 | 2 | 0 | 0 | 1 | 1 | 2 | 1 | 1 | 0 | 1 |
| 2062712 | 0 | 2 | 1 | 1 | 1 | 2 | 2 | 1 | 3 | 4 | 0 | 0 | 1 | 1 | 1 | 1 | 0 | 0 | 0 |
| 2062713 | 1 | 1 | 1 | 3 | 1 | 2 | 2 | 2 | 3 | 4 | 0 | 0 | 1 | 1 | 1 | 1 | 0 | 0 | 0 |
| 2062714 | 1 | 1 | 1 | 2 | 1 | 2 | 2 | 1 | 3 | 4 | 0 | 0 | 1 | 1 | 2 | 1 | 1 | 0 | 1 |
| 2062715 | 1 | 1 | 1 | 1 | 1 | 2 | 2 | 2 | 3 | 4 | 0 | 0 | 1 | 1 | 1 | 1 | 0 | 0 | 0 |
| 2062716 | 1 | 2 | 1 | 3 | 1 | 2 | 1 | 2 | 3 | 4 | 0 | 0 | 1 | 1 | 1 | 1 | 0 | 0 | 0 |
| 2062717 | 0 | 1 | 1 | 3 | 1 | 2 | 2 | 1 | 3 | 4 | 0 | 0 | 1 | 1 | 2 | 1 | 1 | 0 | 1 |
| 2062718 | 1 | 1 | 1 | 1 | 1 | 2 | 1 | 1 | 3 | 4 | 0 | 0 | 1 | 1 | 2 | 1 | 1 | 0 | 1 |
| 2062719 | 0 | 2 | 1 | 2 | 1 | 2 | 2 | 1 | 3 | 4 | 0 | 0 | 1 | 1 | 2 | 1 | 1 | 0 | 1 |
| 2062720 | 0 | 1 | 1 | 3 | 1 | 1 | 2 | 1 | 3 | 4 | 0 | 0 | 1 | 1 | 2 | 1 | 1 | 0 | 1 |
| 2062721 | 1 | 4 | 1 | 1 | 1 | 2 | 1 | 2 | 3 | 4 | 0 | 0 | 1 | 1 | 2 | 1 | 1 | 0 | 1 |
| 2062722 | 0 | 2 | 1 | 3 | 1 | 2 | 1 | 2 | 3 | 4 | 0 | 0 | 1 | 1 | 1 | 1 | 0 | 0 | 0 |
| 2062723 | 1 | 3 | 1 | 2 | 1 | 2 | 1 | 2 | 3 | 4 | 0 | 0 | 1 | 1 | 1 | 1 | 0 | 0 | 0 |
| 2062724 | 0 | 2 | 1 | 2 | 1 | 2 | 2 | 2 | 3 | 4 | 0 | 0 | 1 | 1 | 1 | 1 | 0 | 0 | 0 |
| 2062725 | 1 | 2 | 1 | 2 | 1 | 2 | 2 | 2 | 3 | 4 | 0 | 0 | 1 | 1 | 1 | 1 | 0 | 0 | 0 |
| 2062726 | 1 | 1 | 1 | 1 | 1 | 2 | 1 | 1 | 1 | 2 | 0 | 0 | 1 | 1 | 4 | 5 | 1 | 1 | 1 |
| 2062727 | 1 | 3 | 1 | 1 | 1 | 2 | 2 | 2 | 3 | 2 | 0 | 0 | 1 | 1 | 3 | 6 | 1 | 1 | 1 |
| 2062728 | 0 | 1 | 1 | 2 | 1 | 2 | 2 | 2 | 3 | 2 | 0 | 0 | 1 | 1 | 2 | 1 | 1 | 0 | 1 |
| 2062729 | 0 | 3 | 1 | 1 | 1 | 2 | 2 | 2 | 3 | 2 | 0 | 0 | 1 | 1 | 2 | 1 | 1 | 0 | 1 |
| 2062730 | 0 | 2 | 1 | 2 | 1 | 1 | 1 | 2 | 3 | 2 | 0 | 0 | 1 | 1 | 2 | 1 | 1 | 0 | 1 |
| 206281 | 0 | 3 | 1 | 3 | 1 | 1 | 2 | 2 | 3 | 3 | 0 | 0 | 1 | 1 | 2 | 2 | 1 | 0 | 1 |
| 206282 | 1 | 2 | 1 | 4 | 1 | 2 | 2 | 2 | 3 | 2 | 0 | 0 | 1 | 1 | 5 | 5 | 1 | 1 | 1 |
| 206283 | 1 | 3 | 1 | 2 | 1 | 2 | 2 | 2 | 3 | 1 | 0 | 0 | 1 | 1 | 1 | 1 | 0 | 0 | 0 |
| 206284 | 1 | 3 | 1 | 2 | 1 | 2 | 2 | 2 | 3 | 1 | 0 | 0 | 1 | 1 | 2 | 1 | 1 | 0 | 1 |
| 206285 | 0 | 2 | 1 | 2 | 1 | 3 | 2 | 2 | 3 | 1 | 0 | 0 | 1 | 1 | 3 | 1 | 1 | 0 | 1 |
| 206286 | 0 | 5 | 1 | 2 | 1 | 1 | 2 | 2 | 3 | 1 | 0 | 0 | 1 | 1 | 2 | 1 | 1 | 0 | 1 |
| 206287 | 0 | 2 | 1 | 2 | 1 | 3 | 2 | 2 | 3 | 1 | 0 | 0 | 1 | 1 | 2 | 1 | 1 | 0 | 1 |
| 206288 | 0 | 2 | 1 | 1 | 1 | 1 | 2 | 2 | 2 | 4 | 0 | 0 | 1 | 1 | 2 | 1 | 1 | 0 | 1 |
| 206289 | 0 | 1 | 1 | 4 | 1 | 1 | 2 | 2 | 2 | 4 | 0 | 0 | 1 | 1 | 4 | 1 | 1 | 0 | 1 |
| 2062810 | 1 | 2 | 1 | 1 | 1 | 2 | 2 | 2 | 2 | 4 | 0 | 0 | 1 | 1 | 2 | 1 | 1 | 0 | 1 |
| 2062811 | 1 | 4 | 1 | 1 | 1 | 2 | 1 | 2 | 2 | 4 | 0 | 0 | 1 | 0 | 5 | 5 | 1 | 1 | 1 |
| 2062812 | 1 | 2 | 1 | 1 | 1 | 2 | 1 | 2 | 2 | 4 | 0 | 0 | 1 | 1 | 4 | 2 | 1 | 0 | 1 |
| 2062813 | 0 | 1 | 1 | 3 | 1 | 3 | 1 | 1 | 2 | 4 | 0 | 0 | 1 | 1 | 2 | 1 | 1 | 0 | 1 |
| 2062814 | 1 | 4 | 1 | 4 | 2 | 2 | 2 | 2 | 1 | 1 | 0 | 0 | 1 | 1 | 4 | 2 | 1 | 0 | 1 |

| | | | | | | | | | | | | | | | | | | | |
|---|---|---|---|---|---|---|---|---|---|---|---|---|---|---|---|---|---|---|---|
| 2062815 | 0 | 3 | 1 | 2 | 1 | 1 | 1 | 2 | 1 | 3 | 0 | 0 | 1 | 1 | 5 | 5 | 1 | 1 | 1 |
| 2062816 | 1 | 1 | 1 | 2 | 2 | 2 | 2 | 1 | 1 | 2 | 0 | 0 | 1 | 1 | 2 | 2 | 1 | 0 | 1 |
| 2062817 | 0 | 4 | 1 | 2 | 1 | 1 | 2 | 2 | 1 | 1 | 0 | 0 | 1 | 1 | 1 | 1 | 0 | 0 | 0 |
| 2062818 | 0 | 3 | 1 | 1 | 1 | 1 | 2 | 2 | 2 | 4 | 0 | 0 | 1 | 1 | 4 | 1 | 1 | 0 | 1 |
| 2062819 | 0 | 4 | 1 | 2 | 2 | 5 | 2 | 2 | 1 | 2 | 0 | 0 | 1 | 1 | 2 | 1 | 1 | 0 | 1 |
| 2062820 | 1 | 4 | 1 | 1 | 2 | 2 | 3 | 2 | 1 | 4 | 0 | 0 | 1 | 0 | 5 | 5 | 1 | 1 | 1 |
| 2062821 | 0 | 3 | 1 | 1 | 2 | 2 | 2 | 2 | 2 | 1 | 0 | 0 | 1 | 1 | 3 | 2 | 1 | 0 | 1 |
| 2062822 | 1 | 3 | 1 | 2 | 2 | 2 | 3 | 2 | 1 | 2 | 0 | 0 | 1 | 1 | 1 | 6 | 0 | 1 | 1 |
| 2062823 | 1 | 3 | 1 | 1 | 1 | 2 | 3 | 2 | 1 | 1 | 0 | 0 | 1 | 1 | 2 | 1 | 1 | 0 | 1 |
| 2062824 | 0 | 3 | 1 | 1 | 2 | 2 | 2 | 2 | 1 | 1 | 0 | 0 | 1 | 1 | 1 | 1 | 0 | 0 | 0 |
| 2062825 | 0 | 2 | 1 | 2 | 2 | 1 | 2 | 2 | 1 | 1 | 0 | 0 | 1 | 1 | 1 | 1 | 0 | 0 | 0 |
| 2062826 | 1 | 3 | 1 | 1 | 2 | 2 | 1 | 1 | 1 | 1 | 0 | 0 | 1 | 1 | 3 | 5 | 1 | 1 | 1 |
| 2062827 | 1 | 3 | 1 | 4 | 1 | 3 | 3 | 2 | 1 | 2 | 0 | 0 | 1 | 1 | 5 | 7 | 1 | 1 | 1 |
| 2062828 | 1 | 5 | 1 | 2 | 1 | 2 | 2 | 1 | 3 | 3 | 0 | 0 | 1 | 1 | 2 | 2 | 1 | 0 | 1 |
| 2062829 | 1 | 5 | 1 | 1 | 1 | 2 | 2 | 2 | 3 | 1 | 0 | 0 | 1 | 1 | 3 | 1 | 1 | 0 | 1 |
| 2062830 | 1 | 4 | 1 | 4 | 2 | 2 | 3 | 2 | 1 | 1 | 0 | 0 | 1 | 1 | 1 | 1 | 0 | 0 | 0 |
| 206291 | 1 | 5 | 1 | 1 | 1 | 2 | 2 | 1 | 1 | 1 | 0 | 0 | 1 | 1 | 2 | 1 | 1 | 0 | 1 |
| 206292 | 0 | 3 | 1 | 2 | 1 | 2 | 2 | 2 | 1 | 1 | 0 | 0 | 1 | 1 | 1 | 1 | 0 | 0 | 0 |
| 206293 | 0 | 3 | 1 | 1 | 1 | 3 | 2 | 1 | 1 | 2 | 0 | 0 | 1 | 1 | 1 | 1 | 0 | 0 | 0 |
| 206294 | 1 | 2 | 1 | 2 | 1 | 2 | 2 | 2 | 1 | 3 | 0 | 0 | 1 | 1 | 2 | 1 | 1 | 0 | 1 |
| 206295 | 1 | 5 | 1 | 1 | 1 | 2 | 2 | 2 | 1 | 2 | 1 | 0 | 1 | 1 | 2 | 1 | 1 | 0 | 1 |
| 206296 | 0 | 4 | 1 | 1 | 1 | 2 | 2 | 2 | 1 | 3 | 0 | 0 | 1 | 1 | 4 | 1 | 1 | 0 | 1 |
| 206297 | 1 | 5 | 1 | 1 | 1 | 2 | 2 | 2 | 1 | 4 | 0 | 0 | 1 | 1 | 2 | 1 | 1 | 0 | 1 |
| 206298 | 1 | 5 | 1 | 1 | 1 | 2 | 1 | 1 | 1 | 4 | 0 | 0 | 0 | 99 | 99 | 99 | 99 | 99 | 99 |
| 206299 | 0 | 4 | 1 | 2 | 1 | 2 | 3 | 2 | 1 | 4 | 0 | 0 | 1 | 1 | 1 | 1 | 0 | 0 | 0 |
| 2062910 | 1 | 5 | 1 | 1 | 1 | 2 | 3 | 1 | 1 | 4 | 0 | 0 | 0 | 99 | 99 | 99 | 99 | 99 | 99 |
| 2062911 | 0 | 2 | 1 | 2 | 2 | 1 | 2 | 2 | 1 | 4 | 1 | 1 | 1 | 1 | 2 | 1 | 1 | 0 | 1 |
| 2062912 | 0 | 3 | 1 | 1 | 2 | 2 | 2 | 2 | 1 | 4 | 0 | 0 | 1 | 1 | 2 | 1 | 1 | 0 | 1 |
| 2062913 | 0 | 2 | 1 | 1 | 1 | 2 | 2 | 2 | 1 | 1 | 0 | 0 | 1 | 1 | 1 | 1 | 0 | 0 | 0 |
| 2062914 | 0 | 3 | 1 | 1 | 1 | 2 | 2 | 2 | 1 | 1 | 0 | 0 | 1 | 1 | 2 | 1 | 1 | 0 | 1 |
| 2062915 | 1 | 1 | 1 | 5 | 1 | 4 | 2 | 2 | 1 | 1 | 1 | 1 | 1 | 1 | 1 | 1 | 0 | 0 | 0 |
| 2062916 | 1 | 2 | 1 | 4 | 1 | 5 | 3 | 2 | 1 | 1 | 1 | 1 | 1 | 1 | 2 | 1 | 1 | 0 | 1 |
| 2062917 | 1 | 5 | 1 | 1 | 1 | 2 | 2 | 2 | 1 | 3 | 0 | 0 | 1 | 1 | 1 | 1 | 0 | 0 | 0 |
| 2062918 | 0 | 2 | 1 | 3 | 1 | 1 | 2 | 2 | 1 | 2 | 0 | 0 | 1 | 1 | 2 | 1 | 1 | 0 | 1 |
| 2062919 | 0 | 3 | 1 | 1 | 1 | 1 | 2 | 2 | 1 | 1 | 0 | 0 | 1 | 1 | 2 | 4 | 1 | 1 | 1 |
| 2062920 | 0 | 1 | 1 | 4 | 1 | 3 | 3 | 2 | 1 | 1 | 0 | 0 | 1 | 1 | 2 | 6 | 1 | 1 | 1 |
| 2062921 | 1 | 3 | 1 | 2 | 1 | 2 | 3 | 2 | 1 | 1 | 0 | 0 | 1 | 1 | 1 | 1 | 0 | 0 | 0 |
| 2062922 | 0 | 2 | 1 | 1 | 1 | 1 | 2 | 2 | 1 | 2 | 0 | 0 | 1 | 1 | 3 | 1 | 1 | 0 | 1 |
| 2062923 | 0 | 3 | 1 | 4 | 1 | 1 | 2 | 2 | 1 | 3 | 0 | 0 | 1 | 1 | 2 | 1 | 1 | 0 | 1 |
| 2062924 | 1 | 4 | 1 | 1 | 1 | 2 | 3 | 2 | 1 | 1 | 0 | 0 | 1 | 1 | 3 | 6 | 1 | 1 | 1 |
| 2062925 | 1 | 2 | 1 | 1 | 1 | 2 | 2 | 2 | 1 | 1 | 0 | 0 | 1 | 1 | 2 | 1 | 1 | 0 | 1 |
| 2062926 | 0 | 2 | 1 | 2 | 1 | 1 | 2 | 2 | 1 | 1 | 0 | 0 | 1 | 1 | 2 | 5 | 1 | 1 | 1 |
| 2062927 | 0 | 2 | 1 | 2 | 1 | 1 | 2 | 2 | 1 | 1 | 0 | 0 | 1 | 1 | 1 | 2 | 0 | 0 | 0 |
| 2062928 | 0 | 2 | 1 | 1 | 1 | 1 | 2 | 2 | 1 | 2 | 0 | 0 | 1 | 1 | 1 | 1 | 0 | 0 | 0 |
| 2062929 | 0 | 2 | 1 | 1 | 1 | 1 | 2 | 2 | 1 | 3 | 0 | 0 | 1 | 1 | 2 | 1 | 1 | 0 | 1 |
| 2062930 | 1 | 4 | 1 | 2 | 1 | 2 | 2 | 2 | 1 | 3 | 0 | 0 | 1 | 1 | 2 | 2 | 1 | 0 | 1 |
| 206301 | 0 | 5 | 1 | 2 | 1 | 3 | 2 | 1 | 1 | 1 | 0 | 0 | 1 | 1 | 1 | 1 | 0 | 0 | 0 |
| 206302 | 0 | 4 | 1 | 1 | 1 | 3 | 2 | 1 | 1 | 2 | 0 | 0 | 1 | 1 | 1 | 1 | 0 | 0 | 0 |
| 206303 | 0 | 5 | 1 | 4 | 2 | 1 | 1 | 1 | 1 | 1 | 0 | 0 | 0 | 99 | 99 | 99 | 99 | 99 | 99 |
| 206304 | 0 | 1 | 1 | 4 | 2 | 1 | 2 | 1 | 1 | 4 | 1 | 1 | 1 | 1 | 1 | 1 | 0 | 0 | 0 |
| 206305 | 0 | 5 | 1 | 1 | 1 | 3 | 2 | 1 | 1 | 2 | 0 | 0 | 0 | 99 | 99 | 99 | 99 | 99 | 99 |
| 206306 | 1 | 4 | 1 | 3 | 1 | 2 | 2 | 2 | 1 | 1 | 0 | 0 | 1 | 1 | 1 | 1 | 0 | 0 | 0 |
| 206307 | 1 | 3 | 1 | 3 | 1 | 4 | 2 | 1 | 1 | 1 | 0 | 0 | 1 | 1 | 1 | 1 | 0 | 0 | 0 |
| 206308 | 0 | 5 | 1 | 1 | 1 | 1 | 2 | 1 | 1 | 3 | 0 | 0 | 0 | 99 | 99 | 99 | 99 | 99 | 99 |
| 206309 | 0 | 2 | 1 | 5 | 1 | 5 | 2 | 1 | 1 | 1 | 1 | 1 | 1 | 1 | 1 | 1 | 0 | 0 | 0 |
| 2063010 | 1 | 3 | 1 | 1 | 2 | 4 | 3 | 1 | 1 | 3 | 1 | 0 | 1 | 1 | 1 | 1 | 0 | 0 | 0 |

| | | | | | | | | | | | | | | | | | | |
|---|---|---|---|---|---|---|---|---|---|---|---|---|---|---|---|---|---|---|
| 2063011 | 1 | 4 | 1 | 2 | 1 | 2 | 3 | 1 | 1 | 1 | 1 | 1 | 1 | 1 | 1 | 1 | 0 | 0 | 0 |
| 2063012 | 0 | 2 | 1 | 2 | 2 | 1 | 2 | 1 | 1 | 1 | 0 | 0 | 1 | 1 | 1 | 1 | 0 | 0 | 0 |
| 2063013 | 0 | 1 | 1 | 5 | 2 | 1 | 3 | 1 | 1 | 4 | 0 | 0 | 1 | 1 | 1 | 1 | 0 | 0 | 0 |
| 2063014 | 0 | 5 | 1 | 4 | 2 | 3 | 3 | 1 | 1 | 2 | 1 | 1 | 1 | 1 | 1 | 1 | 0 | 0 | 0 |
| 2063015 | 1 | 5 | 1 | 1 | 2 | 2 | 2 | 1 | 1 | 3 | 0 | 0 | 0 | 99 | 99 | 99 | 99 | 99 | 99 |
| 2063016 | 0 | 3 | 1 | 2 | 1 | 1 | 2 | 1 | 1 | 4 | 0 | 0 | 1 | 1 | 1 | 1 | 0 | 0 | 0 |
| 2063017 | 1 | 4 | 1 | 3 | 1 | 4 | 3 | 1 | 1 | 4 | 1 | 1 | 1 | 1 | 1 | 1 | 0 | 0 | 0 |
| 2063018 | 0 | 3 | 1 | 4 | 1 | 1 | 2 | 1 | 1 | 3 | 1 | 0 | 1 | 1 | 1 | 1 | 0 | 0 | 0 |
| 2063019 | 0 | 3 | 1 | 2 | 2 | 1 | 1 | 1 | 1 | 1 | 0 | 0 | 1 | 1 | 1 | 5 | 0 | 1 | 1 |
| 2063020 | 0 | 2 | 1 | 2 | 2 | 3 | 3 | 1 | 1 | 1 | 1 | 0 | 1 | 1 | 1 | 1 | 0 | 0 | 0 |
| 2063021 | 0 | 1 | 1 | 2 | 1 | 3 | 2 | 1 | 1 | 4 | 0 | 0 | 1 | 1 | 1 | 1 | 0 | 0 | 0 |
| 2063022 | 1 | 2 | 1 | 2 | 1 | 2 | 2 | 1 | 1 | 1 | 0 | 0 | 1 | 1 | 1 | 1 | 0 | 0 | 0 |
| 2063023 | 1 | 2 | 1 | 2 | 1 | 2 | 2 | 1 | 1 | 1 | 0 | 0 | 1 | 1 | 1 | 1 | 0 | 0 | 0 |
| 2063024 | 1 | 3 | 1 | 2 | 1 | 2 | 2 | 1 | 1 | 3 | 0 | 0 | 1 | 1 | 1 | 5 | 0 | 1 | 1 |
| 2063025 | 0 | 5 | 1 | 2 | 1 | 2 | 2 | 1 | 1 | 1 | 0 | 0 | 0 | 99 | 99 | 99 | 99 | 99 | 99 |
| 206311 | 0 | 1 | 1 | 2 | 1 | 1 | 2 | 1 | 4 | 4 | 0 | 0 | 1 | 1 | 1 | 1 | 0 | 0 | 0 |
| 206312 | 0 | 2 | 1 | 3 | 1 | 1 | 2 | 1 | 4 | 4 | 0 | 0 | 1 | 1 | 1 | 1 | 0 | 0 | 0 |
| 206313 | 1 | 2 | 1 | 2 | 1 | 2 | 2 | 2 | 4 | 3 | 0 | 0 | 0 | 99 | 99 | 99 | 99 | 99 | 99 |
| 206314 | 1 | 3 | 1 | 1 | 1 | 2 | 2 | 1 | 4 | 4 | 0 | 0 | 1 | 1 | 1 | 1 | 0 | 0 | 0 |
| 206315 | 0 | 2 | 1 | 2 | 1 | 1 | 2 | 1 | 4 | 2 | 1 | 0 | 1 | 1 | 1 | 1 | 0 | 0 | 0 |
| 206316 | 0 | 1 | 1 | 5 | 1 | 1 | 3 | 1 | 4 | 4 | 0 | 0 | 1 | 1 | 1 | 1 | 0 | 0 | 0 |
| 206317 | 0 | 2 | 1 | 1 | 1 | 3 | 2 | 1 | 4 | 4 | 1 | 0 | 1 | 1 | 1 | 1 | 0 | 0 | 0 |
| 206318 | 1 | 3 | 1 | 1 | 1 | 2 | 2 | 1 | 4 | 1 | 1 | 0 | 1 | 1 | 1 | 1 | 0 | 0 | 0 |
| 206319 | 0 | 5 | 1 | 1 | 1 | 1 | 1 | 1 | 4 | 1 | 0 | 0 | 0 | 99 | 99 | 99 | 99 | 99 | 99 |
| 2063110 | 1 | 2 | 1 | 2 | 1 | 2 | 2 | 1 | 4 | 3 | 1 | 0 | 1 | 1 | 1 | 1 | 0 | 0 | 0 |
| 2063111 | 0 | 3 | 1 | 1 | 1 | 3 | 2 | 1 | 4 | 4 | 0 | 0 | 1 | 1 | 1 | 1 | 0 | 0 | 0 |
| 2063112 | 0 | 2 | 1 | 1 | 1 | 1 | 2 | 1 | 4 | 3 | 0 | 0 | 1 | 1 | 1 | 1 | 0 | 0 | 0 |
| 2063113 | 1 | 2 | 1 | 2 | 1 | 2 | 2 | 1 | 4 | 2 | 0 | 0 | 1 | 1 | 1 | 1 | 0 | 0 | 0 |
| 2063114 | 1 | 3 | 1 | 2 | 1 | 2 | 2 | 1 | 4 | 2 | 1 | 0 | 1 | 1 | 1 | 1 | 0 | 0 | 0 |
| 2063115 | 0 | 3 | 1 | 2 | 1 | 1 | 2 | 1 | 4 | 4 | 0 | 0 | 0 | 99 | 99 | 99 | 99 | 99 | 99 |
| 2063116 | 1 | 2 | 1 | 2 | 1 | 2 | 2 | 1 | 4 | 3 | 1 | 0 | 1 | 1 | 1 | 1 | 0 | 0 | 0 |
| 2063117 | 0 | 3 | 1 | 3 | 2 | 1 | 2 | 1 | 4 | 4 | 0 | 0 | 1 | 1 | 1 | 1 | 0 | 0 | 0 |
| 2063118 | 1 | 3 | 1 | 2 | 1 | 3 | 2 | 1 | 4 | 3 | 0 | 0 | 1 | 1 | 4 | 1 | 1 | 0 | 1 |
| 2063119 | 0 | 2 | 1 | 2 | 1 | 1 | 2 | 1 | 4 | 3 | 0 | 0 | 1 | 1 | 1 | 1 | 0 | 0 | 0 |
| 2063120 | 1 | 1 | 1 | 2 | 1 | 2 | 2 | 1 | 4 | 1 | 0 | 0 | 1 | 1 | 1 | 1 | 0 | 0 | 0 |
| 2063121 | 1 | 2 | 1 | 2 | 1 | 2 | 2 | 1 | 4 | 4 | 0 | 0 | 1 | 1 | 1 | 1 | 0 | 0 | 0 |
| 2063122 | 1 | 4 | 1 | 1 | 1 | 2 | 2 | 1 | 4 | 3 | 0 | 0 | 1 | 1 | 1 | 1 | 0 | 0 | 0 |
| 2063123 | 0 | 2 | 1 | 1 | 1 | 1 | 2 | 1 | 4 | 3 | 0 | 0 | 1 | 1 | 1 | 1 | 0 | 0 | 0 |
| 2063124 | 0 | 1 | 1 | 1 | 1 | 1 | 3 | 1 | 4 | 1 | 0 | 0 | 1 | 1 | 1 | 1 | 0 | 0 | 0 |
| 2063125 | 1 | 2 | 1 | 2 | 1 | 2 | 3 | 2 | 4 | 4 | 0 | 0 | 1 | 1 | 1 | 1 | 0 | 0 | 0 |
| 206321 | 0 | 2 | 1 | 1 | 1 | 1 | 1 | 2 | 4 | 4 | 0 | 0 | 1 | 1 | 2 | 1 | 1 | 0 | 1 |
| 206322 | 0 | 2 | 1 | 1 | 1 | 3 | 1 | 1 | 4 | 4 | 0 | 0 | 1 | 1 | 4 | 1 | 1 | 0 | 1 |
| 206323 | 0 | 1 | 1 | 2 | 1 | 1 | 2 | 1 | 4 | 4 | 0 | 0 | 1 | 1 | 3 | 1 | 1 | 0 | 1 |
| 206324 | 1 | 4 | 1 | 2 | 1 | 2 | 1 | 1 | 4 | 4 | 0 | 0 | 1 | 1 | 2 | 1 | 1 | 0 | 1 |
| 206325 | 0 | 1 | 1 | 3 | 1 | 1 | 2 | 1 | 4 | 4 | 0 | 0 | 1 | 1 | 3 | 1 | 1 | 0 | 1 |
| 206326 | 0 | 2 | 1 | 1 | 1 | 2 | 1 | 2 | 4 | 4 | 0 | 0 | 1 | 1 | 3 | 1 | 1 | 0 | 1 |
| 206327 | 0 | 5 | 1 | 1 | 1 | 2 | 1 | 2 | 4 | 4 | 0 | 0 | 1 | 1 | 3 | 1 | 1 | 0 | 1 |
| 206328 | 0 | 3 | 1 | 1 | 1 | 1 | 1 | 2 | 4 | 4 | 0 | 0 | 1 | 1 | 3 | 2 | 1 | 0 | 1 |
| 206329 | 0 | 1 | 1 | 3 | 1 | 1 | 3 | 2 | 1 | 1 | 0 | 0 | 1 | 1 | 3 | 1 | 1 | 0 | 1 |
| 2063210 | 0 | 1 | 1 | 3 | 1 | 3 | 1 | 2 | 1 | 1 | 0 | 0 | 1 | 1 | 2 | 1 | 1 | 0 | 1 |
| 2063211 | 1 | 4 | 1 | 1 | 1 | 2 | 1 | 2 | 1 | 1 | 0 | 0 | 1 | 1 | 3 | 1 | 1 | 0 | 1 |
| 2063212 | 1 | 3 | 1 | 2 | 1 | 2 | 2 | 2 | 1 | 1 | 1 | 1 | 1 | 1 | 3 | 1 | 1 | 0 | 1 |
| 2063213 | 0 | 3 | 1 | 1 | 1 | 2 | 2 | 2 | 1 | 1 | 0 | 0 | 1 | 1 | 2 | 1 | 1 | 0 | 1 |
| 2063214 | 0 | 2 | 1 | 2 | 2 | 1 | 2 | 2 | 4 | 4 | 0 | 0 | 1 | 1 | 2 | 1 | 1 | 0 | 1 |
| 2063215 | 0 | 1 | 1 | 3 | 1 | 5 | 2 | 2 | 4 | 4 | 0 | 0 | 1 | 1 | 2 | 1 | 1 | 0 | 1 |
| 2063216 | 0 | 4 | 1 | 1 | 1 | 3 | 2 | 1 | 4 | 4 | 0 | 0 | 1 | 1 | 2 | 1 | 1 | 0 | 1 |

| | | | | | | | | | | | | | | | | | | |
|---|---|---|---|---|---|---|---|---|---|---|---|---|---|---|---|---|---|---|
| 2063217 | 1 | 2 | 1 | 2 | 1 | 2 | 2 | 2 | 1 | 2 | 0 | 0 | 1 | 1 | 1 | 1 | 0 | 0 | 0 |
| 2063218 | 1 | 1 | 1 | 4 | 1 | 2 | 2 | 2 | 1 | 2 | 0 | 0 | 1 | 1 | 2 | 1 | 1 | 0 | 1 |
| 2063219 | 0 | 5 | 1 | 2 | 1 | 2 | 2 | 2 | 1 | 1 | 0 | 0 | 1 | 1 | 3 | 1 | 1 | 0 | 1 |
| 2063220 | 0 | 2 | 1 | 1 | 1 | 1 | 2 | 2 | 1 | 1 | 0 | 0 | 1 | 1 | 3 | 1 | 1 | 0 | 1 |
| 2063221 | 1 | 2 | 1 | 2 | 1 | 2 | 2 | 1 | 1 | 4 | 0 | 0 | 1 | 1 | 2 | 1 | 1 | 0 | 1 |
| 2063222 | 0 | 3 | 1 | 2 | 1 | 1 | 3 | 2 | 1 | 1 | 0 | 0 | 1 | 1 | 3 | 1 | 1 | 0 | 1 |
| 2063223 | 0 | 2 | 1 | 2 | 1 | 1 | 2 | 2 | 4 | 1 | 0 | 0 | 1 | 1 | 4 | 1 | 1 | 0 | 1 |
| 2063224 | 0 | 5 | 1 | 1 | 1 | 3 | 2 | 1 | 1 | 1 | 0 | 0 | 1 | 1 | 4 | 1 | 1 | 0 | 1 |
| 2063225 | 0 | 3 | 1 | 1 | 1 | 3 | 1 | 2 | 4 | 4 | 0 | 0 | 1 | 1 | 2 | 4 | 1 | 1 | 1 |
| 2063226 | 1 | 2 | 1 | 2 | 1 | 2 | 1 | 1 | 1 | 2 | 0 | 0 | 1 | 1 | 2 | 1 | 1 | 0 | 1 |
| 2063227 | 1 | 3 | 1 | 3 | 1 | 2 | 2 | 2 | 1 | 2 | 0 | 0 | 1 | 1 | 2 | 1 | 1 | 0 | 1 |
| 2063228 | 1 | 1 | 1 | 3 | 1 | 2 | 2 | 2 | 1 | 2 | 0 | 0 | 1 | 1 | 3 | 1 | 1 | 0 | 1 |
| 2063229 | 0 | 1 | 1 | 4 | 1 | 1 | 3 | 2 | 1 | 1 | 0 | 0 | 1 | 1 | 1 | 1 | 0 | 0 | 0 |
| 2063230 | 1 | 4 | 1 | 1 | 1 | 2 | 1 | 1 | 4 | 4 | 0 | 0 | 1 | 1 | 5 | 5 | 1 | 1 | 1 |
| 206331 | 1 | 3 | 1 | 1 | 1 | 2 | 1 | 1 | 3 | 4 | 0 | 0 | 1 | 1 | 3 | 1 | 1 | 0 | 1 |
| 206332 | 1 | 3 | 1 | 1 | 1 | 2 | 1 | 2 | 3 | 4 | 0 | 0 | 1 | 1 | 3 | 1 | 1 | 0 | 1 |
| 206333 | 0 | 2 | 1 | 2 | 1 | 2 | 1 | 2 | 3 | 4 | 0 | 0 | 1 | 1 | 2 | 1 | 1 | 0 | 1 |
| 206334 | 1 | 5 | 1 | 3 | 1 | 2 | 1 | 1 | 3 | 3 | 0 | 0 | 1 | 0 | 5 | 5 | 1 | 1 | 1 |
| 206335 | 1 | 3 | 1 | 2 | 1 | 2 | 1 | 2 | 1 | 2 | 0 | 0 | 1 | 1 | 1 | 1 | 0 | 0 | 0 |
| 206336 | 1 | 2 | 1 | 2 | 1 | 2 | 2 | 2 | 1 | 1 | 0 | 0 | 1 | 1 | 1 | 1 | 0 | 0 | 0 |
| 206337 | 0 | 3 | 1 | 1 | 1 | 2 | 1 | 2 | 1 | 1 | 0 | 0 | 1 | 1 | 1 | 1 | 0 | 0 | 0 |
| 206338 | 0 | 4 | 1 | 2 | 1 | 1 | 1 | 2 | 1 | 1 | 0 | 0 | 1 | 1 | 1 | 1 | 0 | 0 | 0 |
| 206339 | 1 | 3 | 1 | 2 | 1 | 2 | 2 | 2 | 1 | 2 | 0 | 0 | 1 | 1 | 1 | 1 | 0 | 0 | 0 |
| 2063310 | 0 | 2 | 1 | 1 | 1 | 2 | 1 | 2 | 1 | 1 | 0 | 0 | 1 | 1 | 3 | 5 | 1 | 1 | 1 |
| 2063311 | 1 | 5 | 1 | 2 | 1 | 2 | 1 | 1 | 1 | 1 | 0 | 0 | 1 | 1 | 2 | 1 | 1 | 0 | 1 |
| 2063312 | 1 | 2 | 1 | 1 | 1 | 4 | 1 | 1 | 1 | 1 | 0 | 0 | 1 | 1 | 3 | 1 | 1 | 0 | 1 |
| 2063313 | 1 | 2 | 1 | 2 | 1 | 2 | 2 | 1 | 3 | 1 | 0 | 0 | 1 | 1 | 3 | 1 | 1 | 0 | 1 |
| 2063314 | 0 | 4 | 1 | 2 | 1 | 2 | 1 | 1 | 4 | 2 | 0 | 0 | 1 | 1 | 1 | 1 | 0 | 0 | 0 |
| 2063315 | 1 | 4 | 1 | 2 | 1 | 2 | 2 | 1 | 3 | 1 | 0 | 0 | 1 | 1 | 1 | 1 | 0 | 0 | 0 |
| 2063316 | 0 | 1 | 1 | 2 | 1 | 2 | 1 | 2 | 3 | 1 | 0 | 0 | 1 | 1 | 2 | 1 | 1 | 0 | 1 |
| 2063317 | 0 | 2 | 1 | 1 | 1 | 2 | 1 | 2 | 3 | 1 | 0 | 0 | 1 | 1 | 1 | 1 | 0 | 0 | 0 |
| 2063318 | 0 | 4 | 1 | 1 | 1 | 2 | 1 | 2 | 3 | 1 | 0 | 0 | 1 | 1 | 1 | 1 | 0 | 0 | 0 |
| 2063319 | 1 | 2 | 1 | 1 | 1 | 2 | 1 | 2 | 3 | 1 | 0 | 0 | 1 | 1 | 1 | 1 | 0 | 0 | 0 |
| 2063320 | 1 | 3 | 1 | 2 | 1 | 2 | 2 | 2 | 3 | 1 | 0 | 0 | 1 | 1 | 3 | 1 | 1 | 0 | 1 |
| 2063321 | 0 | 3 | 1 | 4 | 1 | 2 | 1 | 2 | 3 | 2 | 0 | 0 | 1 | 1 | 1 | 1 | 0 | 0 | 0 |
| 2063322 | 0 | 1 | 1 | 2 | 1 | 2 | 1 | 1 | 3 | 1 | 0 | 0 | 1 | 1 | 2 | 1 | 1 | 0 | 1 |
| 2063323 | 1 | 2 | 1 | 4 | 1 | 2 | 2 | 2 | 3 | 3 | 0 | 0 | 1 | 1 | 2 | 1 | 1 | 0 | 1 |
| 2063324 | 1 | 4 | 1 | 2 | 1 | 2 | 1 | 1 | 3 | 3 | 0 | 0 | 1 | 1 | 3 | 1 | 1 | 0 | 1 |
| 2063325 | 0 | 5 | 1 | 2 | 1 | 2 | 1 | 1 | 3 | 3 | 0 | 0 | 1 | 1 | 2 | 1 | 1 | 0 | 1 |
| 2063326 | 1 | 5 | 1 | 1 | 1 | 4 | 1 | 1 | 3 | 4 | 0 | 0 | 1 | 1 | 2 | 1 | 1 | 0 | 1 |
| 2063327 | 0 | 5 | 1 | 2 | 1 | 2 | 1 | 1 | 1 | 1 | 0 | 0 | 1 | 1 | 1 | 1 | 0 | 0 | 0 |
| 2063328 | 1 | 3 | 1 | 1 | 1 | 2 | 1 | 2 | 1 | 1 | 0 | 0 | 1 | 1 | 1 | 1 | 0 | 0 | 0 |
| 2063329 | 0 | 2 | 1 | 1 | 1 | 2 | 2 | 2 | 3 | 1 | 0 | 0 | 1 | 1 | 2 | 1 | 1 | 0 | 1 |
| 2063330 | 0 | 1 | 1 | 2 | 1 | 2 | 1 | 1 | 3 | 1 | 0 | 0 | 1 | 1 | 1 | 1 | 0 | 0 | 0 |
| 307341 | 1 | 5 | 3 | 4 | 4 | 5 | 3 | 2 | 4 | 1 | 1 | 1 | 1 | 1 | 1 | 1 | 0 | 0 | 0 |
| 307342 | 1 | 5 | 3 | 5 | 4 | 5 | 2 | 1 | 4 | 1 | 1 | 1 | 1 | 1 | 1 | 1 | 0 | 0 | 0 |
| 307343 | 0 | 2 | 3 | 3 | 4 | 1 | 1 | 2 | 4 | 2 | 0 | 0 | 1 | 1 | 1 | 1 | 0 | 0 | 0 |
| 307344 | 1 | 4 | 3 | 2 | 4 | 2 | 2 | 1 | 4 | 1 | 0 | 0 | 1 | 1 | 1 | 1 | 0 | 0 | 0 |
| 307345 | 1 | 4 | 3 | 3 | 4 | 2 | 2 | 2 | 4 | 1 | 0 | 0 | 1 | 1 | 1 | 1 | 0 | 0 | 0 |
| 307346 | 1 | 4 | 3 | 5 | 4 | 5 | 2 | 2 | 4 | 1 | 1 | 1 | 1 | 1 | 1 | 1 | 0 | 0 | 0 |
| 307347 | 0 | 3 | 3 | 4 | 4 | 1 | 2 | 2 | 4 | 3 | 1 | 1 | 1 | 1 | 1 | 1 | 0 | 0 | 0 |
| 307348 | 1 | 2 | 3 | 4 | 4 | 5 | 1 | 2 | 4 | 3 | 0 | 0 | 1 | 1 | 1 | 1 | 0 | 0 | 0 |
| 307349 | 1 | 3 | 3 | 3 | 4 | 2 | 2 | 1 | 4 | 3 | 0 | 0 | 1 | 1 | 1 | 1 | 0 | 0 | 0 |
| 3073410 | 0 | 2 | 3 | 3 | 4 | 1 | 1 | 2 | 4 | 3 | 0 | 0 | 1 | 1 | 1 | 1 | 0 | 0 | 0 |
| 3073411 | 1 | 1 | 3 | 3 | 4 | 2 | 2 | 2 | 4 | 3 | 0 | 0 | 1 | 1 | 1 | 1 | 0 | 0 | 0 |
| 3073412 | 0 | 1 | 3 | 5 | 4 | 5 | 3 | 1 | 4 | 3 | 1 | 1 | 1 | 1 | 1 | 1 | 0 | 0 | 0 |

| | | | | | | | | | | | | | | | | | | |
|---|---|---|---|---|---|---|---|---|---|---|---|---|---|---|---|---|---|---|
| 3073413 | 0 | 1 | 3 | 2 | 4 | 1 | 1 | 2 | 4 | 3 | 0 | 0 | 1 | 1 | 1 | 1 | 0 | 0 | 0 |
| 3073414 | 1 | 5 | 3 | 3 | 4 | 2 | 2 | 2 | 4 | 3 | 0 | 0 | 0 | 99 | 99 | 99 | 99 | 99 | 99 |
| 3073415 | 1 | 4 | 3 | 2 | 4 | 2 | 1 | 2 | 4 | 3 | 0 | 0 | 1 | 1 | 1 | 1 | 0 | 0 | 0 |
| 3073416 | 1 | 2 | 3 | 3 | 4 | 2 | 1 | 1 | 4 | 3 | 0 | 0 | 1 | 1 | 1 | 1 | 0 | 0 | 0 |
| 3073417 | 1 | 2 | 3 | 3 | 4 | 2 | 1 | 2 | 4 | 3 | 0 | 0 | 1 | 1 | 1 | 1 | 0 | 0 | 0 |
| 3073418 | 1 | 4 | 3 | 2 | 4 | 2 | 1 | 1 | 4 | 3 | 0 | 0 | 0 | 99 | 99 | 99 | 99 | 99 | 99 |
| 3073419 | 1 | 2 | 3 | 3 | 4 | 2 | 1 | 2 | 4 | 3 | 0 | 0 | 1 | 1 | 1 | 1 | 0 | 0 | 0 |
| 3073420 | 1 | 2 | 3 | 2 | 4 | 2 | 1 | 2 | 4 | 3 | 0 | 0 | 1 | 1 | 1 | 1 | 0 | 0 | 0 |
| 3073421 | 1 | 3 | 3 | 2 | 4 | 2 | 2 | 1 | 4 | 3 | 0 | 0 | 1 | 1 | 1 | 1 | 0 | 0 | 0 |
| 3073422 | 1 | 3 | 3 | 2 | 4 | 2 | 2 | 2 | 4 | 3 | 0 | 0 | 1 | 1 | 1 | 1 | 0 | 0 | 0 |
| 3073423 | 1 | 4 | 3 | 4 | 4 | 5 | 2 | 2 | 4 | 3 | 0 | 0 | 1 | 1 | 1 | 4 | 0 | 1 | 1 |
| 3073424 | 1 | 2 | 3 | 5 | 4 | 5 | 3 | 2 | 4 | 3 | 1 | 1 | 1 | 1 | 1 | 1 | 0 | 0 | 0 |
| 3073425 | 1 | 3 | 3 | 2 | 4 | 2 | 1 | 2 | 4 | 3 | 0 | 0 | 1 | 1 | 1 | 1 | 0 | 0 | 0 |
| 3073426 | 1 | 4 | 3 | 2 | 4 | 2 | 1 | 1 | 4 | 3 | 0 | 0 | 1 | 1 | 1 | 1 | 0 | 0 | 0 |
| 3073427 | 1 | 4 | 3 | 2 | 4 | 2 | 1 | 2 | 4 | 3 | 0 | 0 | 1 | 1 | 1 | 1 | 0 | 0 | 0 |
| 3073428 | 1 | 1 | 3 | 5 | 4 | 5 | 3 | 2 | 4 | 3 | 1 | 1 | 1 | 1 | 1 | 1 | 0 | 0 | 0 |
| 3073429 | 1 | 2 | 3 | 4 | 4 | 2 | 2 | 1 | 4 | 3 | 0 | 0 | 1 | 1 | 1 | 1 | 0 | 0 | 0 |
| 3073430 | 1 | 4 | 3 | 4 | 4 | 2 | 1 | 1 | 4 | 3 | 0 | 0 | 0 | 99 | 99 | 99 | 99 | 99 | 99 |
| 3073431 | 1 | 4 | 3 | 2 | 4 | 2 | 1 | 1 | 4 | 3 | 0 | 0 | 1 | 1 | 1 | 1 | 0 | 0 | 0 |
| 3073432 | 1 | 2 | 3 | 5 | 4 | 5 | 2 | 2 | 4 | 3 | 1 | 1 | 1 | 1 | 1 | 1 | 0 | 0 | 0 |
| 307351 | 0 | 2 | 3 | 5 | 4 | 5 | 3 | 2 | 4 | 2 | 1 | 1 | 1 | 1 | 1 | 1 | 0 | 0 | 0 |
| 307352 | 1 | 2 | 3 | 2 | 4 | 2 | 1 | 2 | 4 | 2 | 0 | 0 | 1 | 1 | 1 | 1 | 0 | 0 | 0 |
| 307353 | 1 | 2 | 3 | 2 | 4 | 2 | 1 | 2 | 4 | 2 | 0 | 0 | 1 | 1 | 1 | 1 | 0 | 0 | 0 |
| 307354 | 0 | 4 | 3 | 2 | 4 | 1 | 1 | 2 | 4 | 2 | 0 | 0 | 1 | 1 | 1 | 1 | 0 | 0 | 0 |
| 307355 | 1 | 3 | 3 | 3 | 4 | 2 | 2 | 2 | 4 | 2 | 0 | 0 | 1 | 1 | 1 | 1 | 0 | 0 | 0 |
| 307356 | 0 | 2 | 3 | 2 | 4 | 1 | 1 | 2 | 4 | 2 | 0 | 0 | 1 | 1 | 1 | 1 | 0 | 0 | 0 |
| 307357 | 0 | 5 | 3 | 2 | 4 | 1 | 1 | 1 | 4 | 2 | 0 | 0 | 1 | 1 | 1 | 1 | 0 | 0 | 0 |
| 307358 | 0 | 4 | 3 | 2 | 4 | 1 | 1 | 1 | 4 | 2 | 0 | 0 | 1 | 1 | 1 | 1 | 0 | 0 | 0 |
| 307359 | 0 | 2 | 3 | 3 | 4 | 1 | 1 | 1 | 4 | 2 | 0 | 0 | 1 | 1 | 1 | 1 | 0 | 0 | 0 |
| 3073510 | 1 | 2 | 3 | 3 | 4 | 2 | 1 | 2 | 4 | 2 | 0 | 0 | 1 | 1 | 1 | 1 | 0 | 0 | 0 |
| 3073511 | 1 | 5 | 3 | 2 | 4 | 2 | 2 | 1 | 4 | 2 | 0 | 0 | 1 | 1 | 1 | 1 | 0 | 0 | 0 |
| 3073512 | 0 | 1 | 3 | 4 | 4 | 1 | 2 | 2 | 4 | 2 | 1 | 1 | 1 | 1 | 1 | 1 | 0 | 0 | 0 |
| 3073513 | 1 | 3 | 3 | 2 | 4 | 2 | 1 | 2 | 4 | 2 | 0 | 0 | 1 | 1 | 1 | 1 | 0 | 0 | 0 |
| 3073514 | 1 | 3 | 3 | 3 | 4 | 2 | 1 | 2 | 4 | 2 | 0 | 0 | 1 | 1 | 1 | 1 | 0 | 0 | 0 |
| 3073515 | 0 | 3 | 3 | 2 | 4 | 1 | 1 | 2 | 4 | 2 | 0 | 0 | 1 | 1 | 1 | 1 | 0 | 0 | 0 |
| 3073516 | 0 | 3 | 3 | 2 | 4 | 1 | 1 | 2 | 4 | 2 | 0 | 0 | 1 | 1 | 1 | 1 | 0 | 0 | 0 |
| 3073517 | 0 | 4 | 3 | 2 | 4 | 1 | 1 | 2 | 4 | 2 | 0 | 0 | 1 | 1 | 1 | 1 | 0 | 0 | 0 |
| 3073518 | 0 | 2 | 3 | 3 | 4 | 1 | 1 | 1 | 4 | 2 | 0 | 0 | 1 | 1 | 1 | 1 | 0 | 0 | 0 |
| 3073519 | 0 | 2 | 3 | 3 | 4 | 1 | 1 | 2 | 4 | 2 | 0 | 0 | 1 | 1 | 1 | 1 | 0 | 0 | 0 |
| 3073520 | 1 | 2 | 3 | 2 | 4 | 2 | 1 | 2 | 4 | 2 | 0 | 0 | 1 | 1 | 1 | 1 | 0 | 0 | 0 |
| 3073521 | 0 | 2 | 3 | 5 | 4 | 5 | 2 | 1 | 4 | 1 | 1 | 1 | 1 | 1 | 1 | 1 | 0 | 0 | 0 |
| 3073522 | 1 | 2 | 3 | 5 | 4 | 5 | 2 | 1 | 4 | 1 | 1 | 1 | 1 | 1 | 1 | 1 | 0 | 0 | 0 |
| 3073523 | 0 | 3 | 3 | 3 | 4 | 1 | 2 | 2 | 4 | 2 | 0 | 0 | 0 | 99 | 99 | 99 | 99 | 99 | 99 |
| 3073524 | 1 | 3 | 3 | 5 | 4 | 5 | 3 | 2 | 4 | 2 | 1 | 1 | 1 | 1 | 1 | 1 | 0 | 0 | 0 |
| 3073525 | 1 | 2 | 3 | 5 | 4 | 5 | 3 | 1 | 4 | 1 | 1 | 1 | 1 | 1 | 1 | 1 | 0 | 0 | 0 |
| 3073526 | 1 | 4 | 3 | 4 | 4 | 5 | 3 | 1 | 4 | 2 | 1 | 1 | 1 | 1 | 1 | 1 | 0 | 0 | 0 |
| 3073527 | 1 | 1 | 3 | 5 | 4 | 5 | 2 | 1 | 4 | 1 | 1 | 1 | 1 | 1 | 1 | 1 | 0 | 0 | 0 |
| 3073528 | 1 | 3 | 3 | 4 | 4 | 5 | 3 | 2 | 4 | 2 | 1 | 1 | 1 | 1 | 1 | 1 | 0 | 0 | 0 |
| 3073529 | 1 | 2 | 3 | 5 | 4 | 5 | 3 | 1 | 4 | 1 | 1 | 1 | 1 | 1 | 1 | 1 | 0 | 0 | 0 |
| 3073530 | 1 | 2 | 3 | 2 | 4 | 2 | 2 | 1 | 4 | 1 | 0 | 0 | 1 | 1 | 1 | 1 | 0 | 0 | 0 |
| 3073531 | 1 | 2 | 3 | 3 | 4 | 2 | 2 | 2 | 4 | 1 | 0 | 0 | 1 | 1 | 1 | 1 | 0 | 0 | 0 |
| 3073532 | 1 | 5 | 3 | 4 | 4 | 5 | 2 | 1 | 4 | 1 | 1 | 0 | 1 | 1 | 1 | 1 | 0 | 0 | 0 |
| 3073533 | 1 | 2 | 3 | 5 | 4 | 5 | 2 | 1 | 4 | 1 | 1 | 1 | 1 | 1 | 1 | 1 | 0 | 0 | 0 |
| 307361 | 1 | 2 | 3 | 4 | 4 | 2 | 2 | 2 | 3 | 1 | 1 | 1 | 1 | 1 | 1 | 1 | 0 | 0 | 0 |
| 307362 | 1 | 2 | 3 | 5 | 4 | 5 | 3 | 1 | 3 | 1 | 1 | 1 | 1 | 1 | 1 | 1 | 0 | 0 | 0 |
| 307363 | 1 | 5 | 3 | 5 | 4 | 5 | 2 | 1 | 3 | 1 | 1 | 1 | 1 | 1 | 3 | 1 | 1 | 0 | 1 |

| | | | | | | | | | | | | | | | | | | | |
|---|---|---|---|---|---|---|---|---|---|---|---|---|---|---|---|---|---|---|---|
| 307364 | 1 | 5 | 3 | 2 | 4 | 2 | 2 | 1 | 3 | 1 | 0 | 0 | 1 | 1 | 1 | 1 | 0 | 0 | 0 |
| 307365 | 0 | 3 | 3 | 3 | 4 | 1 | 2 | 2 | 3 | 1 | 0 | 0 | 1 | 1 | 1 | 1 | 0 | 0 | 0 |
| 307366 | 1 | 4 | 3 | 4 | 4 | 2 | 2 | 2 | 3 | 1 | 0 | 0 | 1 | 1 | 3 | 1 | 1 | 0 | 1 |
| 307367 | 1 | 5 | 3 | 2 | 4 | 2 | 1 | 1 | 3 | 4 | 0 | 0 | 1 | 1 | 1 | 1 | 0 | 0 | 0 |
| 307368 | 1 | 4 | 3 | 5 | 2 | 5 | 3 | 2 | 3 | 1 | 1 | 1 | 1 | 1 | 1 | 2 | 0 | 0 | 0 |
| 307369 | 0 | 4 | 3 | 5 | 4 | 5 | 3 | 2 | 3 | 1 | 1 | 1 | 1 | 1 | 1 | 1 | 0 | 0 | 0 |
| 3073610 | 0 | 2 | 3 | 2 | 4 | 1 | 2 | 1 | 3 | 1 | 0 | 0 | 1 | 1 | 4 | 1 | 1 | 0 | 1 |
| 3073611 | 0 | 4 | 3 | 2 | 4 | 2 | 2 | 1 | 3 | 1 | 0 | 0 | 1 | 1 | 1 | 1 | 0 | 0 | 0 |
| 3073612 | 1 | 3 | 3 | 5 | 4 | 5 | 3 | 1 | 3 | 2 | 0 | 0 | 1 | 1 | 1 | 1 | 0 | 0 | 0 |
| 3073613 | 0 | 2 | 3 | 4 | 4 | 5 | 2 | 1 | 3 | 2 | 1 | 0 | 1 | 1 | 3 | 1 | 1 | 0 | 1 |
| 3073614 | 1 | 5 | 3 | 2 | 4 | 2 | 1 | 1 | 3 | 2 | 0 | 0 | 1 | 1 | 3 | 1 | 1 | 0 | 1 |
| 3073615 | 0 | 3 | 3 | 4 | 4 | 5 | 3 | 1 | 3 | 2 | 1 | 1 | 1 | 1 | 1 | 2 | 0 | 0 | 0 |
| 3073616 | 1 | 4 | 3 | 2 | 4 | 2 | 1 | 2 | 3 | 2 | 0 | 0 | 1 | 1 | 4 | 1 | 1 | 0 | 1 |
| 3073617 | 1 | 4 | 3 | 5 | 4 | 5 | 3 | 1 | 3 | 2 | 1 | 1 | 1 | 1 | 1 | 1 | 0 | 0 | 0 |
| 3073618 | 1 | 2 | 3 | 4 | 4 | 5 | 3 | 1 | 3 | 2 | 0 | 0 | 1 | 1 | 3 | 1 | 1 | 0 | 1 |
| 3073619 | 1 | 5 | 3 | 2 | 4 | 2 | 2 | 1 | 3 | 2 | 0 | 0 | 1 | 1 | 3 | 1 | 1 | 0 | 1 |
| 3073620 | 1 | 2 | 3 | 2 | 4 | 2 | 2 | 2 | 3 | 2 | 0 | 0 | 1 | 1 | 1 | 1 | 0 | 0 | 0 |
| 3073621 | 1 | 2 | 3 | 4 | 4 | 2 | 2 | 2 | 3 | 2 | 0 | 0 | 1 | 1 | 3 | 1 | 1 | 0 | 1 |
| 3073622 | 0 | 3 | 3 | 3 | 4 | 2 | 2 | 2 | 3 | 4 | 0 | 0 | 1 | 1 | 3 | 1 | 1 | 0 | 1 |
| 3073623 | 1 | 5 | 3 | 4 | 4 | 2 | 2 | 1 | 3 | 4 | 0 | 0 | 1 | 1 | 3 | 1 | 1 | 0 | 1 |
| 3073624 | 1 | 4 | 3 | 3 | 4 | 2 | 1 | 2 | 3 | 4 | 0 | 0 | 1 | 1 | 3 | 1 | 1 | 0 | 1 |
| 3073625 | 0 | 5 | 3 | 2 | 4 | 1 | 2 | 1 | 3 | 4 | 0 | 0 | 1 | 1 | 3 | 1 | 1 | 0 | 1 |
| 3073626 | 1 | 4 | 3 | 4 | 4 | 2 | 2 | 2 | 3 | 4 | 0 | 0 | 1 | 1 | 4 | 1 | 1 | 0 | 1 |
| 3073627 | 0 | 3 | 3 | 2 | 4 | 1 | 1 | 1 | 3 | 4 | 0 | 0 | 1 | 1 | 1 | 4 | 0 | 1 | 1 |
| 3073628 | 1 | 5 | 3 | 2 | 4 | 2 | 2 | 2 | 3 | 4 | 0 | 0 | 1 | 1 | 4 | 1 | 1 | 0 | 1 |
| 3073629 | 1 | 2 | 3 | 3 | 4 | 2 | 2 | 2 | 3 | 4 | 0 | 0 | 1 | 1 | 3 | 1 | 1 | 0 | 1 |
| 3073630 | 1 | 4 | 3 | 3 | 4 | 2 | 1 | 2 | 3 | 2 | 0 | 0 | 1 | 1 | 4 | 1 | 1 | 0 | 1 |
| 307371 | 0 | 1 | 3 | 4 | 4 | 1 | 2 | 1 | 1 | 3 | 0 | 0 | 1 | 1 | 1 | 1 | 0 | 0 | 0 |
| 307372 | 0 | 2 | 3 | 4 | 4 | 1 | 2 | 1 | 1 | 3 | 0 | 0 | 1 | 1 | 1 | 1 | 0 | 0 | 0 |
| 307373 | 0 | 2 | 3 | 2 | 4 | 1 | 2 | 2 | 1 | 2 | 0 | 0 | 1 | 1 | 1 | 1 | 0 | 0 | 0 |
| 307374 | 0 | 3 | 3 | 2 | 4 | 1 | 2 | 2 | 1 | 1 | 0 | 0 | 0 | 99 | 99 | 99 | 99 | 99 | 99 |
| 307375 | 0 | 2 | 3 | 2 | 4 | 2 | 1 | 2 | 1 | 4 | 0 | 0 | 1 | 1 | 3 | 1 | 1 | 0 | 1 |
| 307376 | 0 | 1 | 3 | 4 | 4 | 2 | 1 | 1 | 1 | 4 | 0 | 0 | 1 | 1 | 4 | 1 | 1 | 0 | 1 |
| 307377 | 1 | 2 | 3 | 2 | 4 | 2 | 2 | 2 | 1 | 1 | 1 | 1 | 1 | 1 | 1 | 1 | 0 | 0 | 0 |
| 307378 | 0 | 5 | 3 | 2 | 4 | 1 | 2 | 1 | 1 | 4 | 0 | 0 | 0 | 99 | 99 | 99 | 99 | 99 | 99 |
| 307379 | 1 | 2 | 3 | 2 | 4 | 2 | 2 | 2 | 1 | 1 | 0 | 0 | 1 | 1 | 1 | 1 | 0 | 0 | 0 |
| 3073710 | 0 | 3 | 3 | 3 | 4 | 1 | 2 | 1 | 1 | 1 | 0 | 0 | 1 | 1 | 1 | 1 | 0 | 0 | 0 |
| 3073711 | 1 | 1 | 3 | 2 | 4 | 2 | 1 | 1 | 1 | 4 | 0 | 0 | 0 | 99 | 99 | 99 | 99 | 99 | 99 |
| 3073712 | 1 | 5 | 3 | 1 | 4 | 2 | 1 | 1 | 1 | 4 | 0 | 0 | 0 | 99 | 99 | 99 | 99 | 99 | 99 |
| 3073713 | 1 | 1 | 3 | 2 | 2 | 2 | 1 | 1 | 1 | 3 | 0 | 0 | 1 | 1 | 3 | 1 | 1 | 0 | 1 |
| 3073714 | 1 | 4 | 3 | 4 | 4 | 2 | 2 | 2 | 1 | 3 | 0 | 0 | 1 | 1 | 2 | 1 | 1 | 0 | 1 |
| 3073715 | 0 | 2 | 3 | 3 | 4 | 3 | 3 | 2 | 1 | 2 | 1 | 0 | 1 | 1 | 1 | 1 | 0 | 0 | 0 |
| 3073716 | 1 | 2 | 3 | 2 | 4 | 2 | 2 | 1 | 4 | 3 | 0 | 0 | 1 | 1 | 1 | 1 | 0 | 0 | 0 |
| 3073717 | 0 | 1 | 3 | 3 | 4 | 1 | 1 | 1 | 4 | 4 | 0 | 0 | 1 | 1 | 2 | 1 | 1 | 0 | 1 |
| 3073718 | 0 | 2 | 3 | 3 | 4 | 1 | 2 | 2 | 4 | 4 | 0 | 0 | 0 | 99 | 99 | 99 | 99 | 99 | 99 |
| 3073719 | 1 | 5 | 3 | 2 | 4 | 2 | 1 | 1 | 4 | 4 | 0 | 0 | 0 | 99 | 99 | 99 | 99 | 99 | 99 |
| 3073720 | 0 | 2 | 3 | 3 | 4 | 1 | 1 | 1 | 4 | 4 | 0 | 0 | 1 | 1 | 2 | 1 | 1 | 0 | 1 |
| 3073721 | 1 | 5 | 3 | 2 | 4 | 2 | 2 | 2 | 4 | 3 | 0 | 0 | 1 | 1 | 2 | 1 | 1 | 0 | 1 |
| 3073722 | 0 | 2 | 3 | 2 | 4 | 1 | 1 | 1 | 4 | 4 | 0 | 0 | 1 | 1 | 2 | 2 | 1 | 0 | 1 |
| 3073723 | 0 | 3 | 3 | 2 | 4 | 1 | 2 | 2 | 4 | 4 | 0 | 0 | 1 | 1 | 1 | 1 | 0 | 0 | 0 |
| 3073724 | 0 | 1 | 3 | 2 | 4 | 1 | 2 | 1 | 4 | 2 | 0 | 0 | 1 | 1 | 1 | 1 | 0 | 0 | 0 |
| 3073725 | 0 | 4 | 3 | 3 | 4 | 3 | 2 | 1 | 4 | 3 | 0 | 0 | 1 | 1 | 1 | 1 | 0 | 0 | 0 |
| 3073726 | 1 | 2 | 3 | 2 | 4 | 4 | 2 | 1 | 4 | 3 | 0 | 0 | 0 | 99 | 99 | 99 | 99 | 99 | 99 |
| 3073727 | 1 | 2 | 3 | 2 | 4 | 4 | 2 | 2 | 4 | 3 | 0 | 0 | 1 | 1 | 1 | 1 | 0 | 0 | 0 |
| 3073728 | 0 | 1 | 3 | 2 | 4 | 1 | 1 | 2 | 4 | 2 | 0 | 0 | 0 | 99 | 99 | 99 | 99 | 99 | 99 |
| 3073729 | 0 | 2 | 3 | 3 | 4 | 1 | 2 | 2 | 4 | 3 | 0 | 0 | 0 | 99 | 99 | 99 | 99 | 99 | 99 |

| | | | | | | | | | | | | | | | | | | | |
|---|---|---|---|---|---|---|---|---|---|---|---|---|---|---|---|---|---|---|---|
| 3073730 | 0 | 1 | 3 | 2 | 3 | 1 | 1 | 1 | 4 | 2 | 0 | 0 | 0 | 99 | 99 | 99 | 99 | 99 | 99 |
| 307381 | 0 | 2 | 3 | 2 | 4 | 2 | 2 | 1 | 4 | 1 | 0 | 0 | 1 | 1 | 1 | 4 | 0 | 1 | 1 |
| 307382 | 1 | 4 | 3 | 3 | 4 | 2 | 2 | 2 | 4 | 1 | 0 | 0 | 1 | 1 | 1 | 4 | 0 | 1 | 1 |
| 307383 | 0 | 2 | 3 | 3 | 4 | 2 | 2 | 2 | 4 | 1 | 0 | 0 | 1 | 1 | 1 | 4 | 0 | 1 | 1 |
| 307384 | 1 | 3 | 3 | 2 | 4 | 2 | 2 | 2 | 4 | 1 | 0 | 0 | 1 | 1 | 1 | 4 | 0 | 1 | 1 |
| 307385 | 0 | 1 | 3 | 4 | 4 | 2 | 2 | 2 | 4 | 1 | 0 | 0 | 1 | 1 | 1 | 1 | 0 | 0 | 0 |
| 307386 | 1 | 3 | 3 | 2 | 4 | 2 | 2 | 2 | 4 | 1 | 0 | 0 | 1 | 1 | 1 | 4 | 0 | 1 | 1 |
| 307387 | 0 | 1 | 3 | 3 | 4 | 2 | 2 | 2 | 4 | 1 | 0 | 0 | 1 | 1 | 1 | 4 | 0 | 1 | 1 |
| 307388 | 1 | 3 | 3 | 3 | 4 | 2 | 2 | 1 | 4 | 1 | 0 | 0 | 1 | 1 | 3 | 1 | 1 | 0 | 1 |
| 307389 | 1 | 3 | 3 | 4 | 4 | 2 | 2 | 1 | 4 | 1 | 0 | 0 | 1 | 1 | 3 | 1 | 1 | 0 | 1 |
| 3073810 | 0 | 2 | 3 | 4 | 4 | 3 | 2 | 1 | 4 | 1 | 1 | 0 | 1 | 1 | 1 | 1 | 0 | 0 | 0 |
| 3073811 | 0 | 2 | 3 | 5 | 4 | 5 | 2 | 2 | 4 | 1 | 1 | 0 | 1 | 1 | 1 | 1 | 0 | 0 | 0 |
| 3073812 | 1 | 2 | 3 | 3 | 4 | 2 | 2 | 2 | 4 | 1 | 0 | 0 | 1 | 1 | 1 | 1 | 0 | 0 | 0 |
| 3073813 | 0 | 3 | 3 | 2 | 4 | 2 | 2 | 1 | 4 | 1 | 0 | 0 | 1 | 1 | 1 | 4 | 0 | 1 | 1 |
| 3073814 | 1 | 2 | 3 | 5 | 4 | 5 | 2 | 2 | 4 | 1 | 0 | 0 | 1 | 1 | 3 | 1 | 1 | 0 | 1 |
| 3073815 | 0 | 2 | 3 | 5 | 4 | 5 | 3 | 1 | 4 | 1 | 1 | 0 | 1 | 1 | 3 | 1 | 1 | 0 | 1 |
| 3073816 | 1 | 2 | 3 | 2 | 4 | 2 | 1 | 2 | 4 | 3 | 0 | 0 | 1 | 1 | 1 | 1 | 0 | 0 | 0 |
| 3073817 | 0 | 2 | 3 | 5 | 4 | 5 | 3 | 1 | 4 | 1 | 1 | 0 | 1 | 1 | 1 | 1 | 0 | 0 | 0 |
| 3073818 | 1 | 4 | 3 | 3 | 4 | 2 | 2 | 2 | 4 | 2 | 0 | 0 | 1 | 1 | 1 | 1 | 0 | 0 | 0 |
| 3073819 | 0 | 1 | 3 | 4 | 4 | 3 | 3 | 1 | 4 | 1 | 1 | 0 | 1 | 1 | 1 | 1 | 0 | 0 | 0 |
| 3073820 | 1 | 3 | 3 | 4 | 4 | 2 | 2 | 2 | 4 | 2 | 0 | 0 | 1 | 1 | 3 | 1 | 1 | 0 | 1 |
| 3073821 | 0 | 2 | 3 | 5 | 4 | 5 | 2 | 1 | 4 | 4 | 0 | 0 | 1 | 1 | 1 | 1 | 0 | 0 | 0 |
| 3073822 | 1 | 4 | 3 | 2 | 4 | 2 | 2 | 2 | 4 | 3 | 0 | 0 | 1 | 1 | 1 | 1 | 0 | 0 | 0 |
| 3073823 | 0 | 2 | 3 | 3 | 4 | 2 | 2 | 2 | 4 | 2 | 0 | 0 | 1 | 1 | 1 | 1 | 0 | 0 | 0 |
| 3073824 | 1 | 4 | 3 | 2 | 4 | 2 | 2 | 2 | 4 | 3 | 0 | 0 | 1 | 1 | 1 | 1 | 0 | 0 | 0 |
| 3073825 | 0 | 1 | 3 | 4 | 4 | 2 | 2 | 1 | 4 | 3 | 0 | 0 | 1 | 1 | 1 | 1 | 0 | 0 | 0 |
| 3073826 | 0 | 4 | 3 | 2 | 4 | 1 | 2 | 1 | 4 | 3 | 0 | 0 | 1 | 1 | 1 | 1 | 0 | 0 | 0 |
| 3073827 | 1 | 4 | 3 | 2 | 2 | 2 | 2 | 2 | 4 | 3 | 0 | 0 | 1 | 1 | 1 | 1 | 0 | 0 | 0 |
| 3073828 | 1 | 4 | 3 | 4 | 4 | 5 | 3 | 2 | 4 | 2 | 1 | 1 | 1 | 1 | 1 | 1 | 0 | 0 | 0 |
| 3073829 | 0 | 2 | 3 | 2 | 4 | 2 | 2 | 2 | 4 | 4 | 0 | 0 | 1 | 1 | 1 | 4 | 0 | 1 | 1 |
| 3073830 | 1 | 4 | 3 | 2 | 4 | 2 | 2 | 2 | 4 | 3 | 0 | 0 | 1 | 1 | 1 | 1 | 0 | 0 | 0 |
| 307391 | 1 | 3 | 3 | 4 | 4 | 5 | 2 | 2 | 1 | 3 | 1 | 1 | 1 | 1 | 1 | 1 | 0 | 0 | 0 |
| 307392 | 1 | 1 | 3 | 4 | 4 | 2 | 2 | 2 | 2 | 3 | 0 | 0 | 1 | 1 | 1 | 6 | 0 | 1 | 1 |
| 307393 | 1 | 2 | 3 | 2 | 4 | 2 | 2 | 2 | 2 | 3 | 0 | 0 | 1 | 1 | 2 | 1 | 1 | 0 | 1 |
| 307394 | 0 | 2 | 3 | 2 | 4 | 1 | 2 | 2 | 2 | 3 | 0 | 0 | 0 | 99 | 99 | 99 | 99 | 99 | 99 |
| 307395 | 1 | 1 | 3 | 4 | 4 | 2 | 2 | 1 | 2 | 3 | 0 | 0 | 0 | 99 | 99 | 99 | 99 | 99 | 99 |
| 307396 | 0 | 2 | 3 | 2 | 4 | 1 | 2 | 2 | 3 | 2 | 0 | 0 | 0 | 99 | 99 | 99 | 99 | 99 | 99 |
| 307397 | 1 | 3 | 3 | 2 | 4 | 2 | 2 | 2 | 3 | 2 | 0 | 0 | 0 | 99 | 99 | 99 | 99 | 99 | 99 |
| 307398 | 1 | 4 | 3 | 3 | 4 | 2 | 2 | 1 | 2 | 3 | 0 | 0 | 0 | 99 | 99 | 99 | 99 | 99 | 99 |
| 307399 | 0 | 4 | 3 | 3 | 4 | 1 | 1 | 1 | 2 | 2 | 0 | 0 | 1 | 1 | 4 | 1 | 1 | 0 | 1 |
| 3073910 | 1 | 4 | 3 | 2 | 4 | 2 | 2 | 2 | 2 | 2 | 0 | 0 | 1 | 1 | 1 | 1 | 0 | 0 | 0 |
| 3073911 | 1 | 5 | 3 | 3 | 4 | 2 | 1 | 1 | 2 | 2 | 0 | 0 | 1 | 1 | 1 | 1 | 0 | 0 | 0 |
| 3073912 | 0 | 1 | 3 | 5 | 4 | 5 | 2 | 1 | 2 | 1 | 0 | 0 | 1 | 1 | 1 | 1 | 0 | 0 | 0 |
| 3073913 | 1 | 3 | 3 | 5 | 4 | 5 | 2 | 1 | 2 | 1 | 1 | 0 | 1 | 1 | 1 | 1 | 0 | 0 | 0 |
| 3073914 | 0 | 2 | 3 | 2 | 4 | 1 | 1 | 2 | 2 | 2 | 0 | 0 | 0 | 99 | 99 | 99 | 99 | 99 | 99 |
| 3073915 | 0 | 1 | 3 | 3 | 4 | 1 | 2 | 1 | 2 | 1 | 0 | 0 | 0 | 99 | 99 | 99 | 99 | 99 | 99 |
| 3073916 | 0 | 2 | 3 | 2 | 4 | 2 | 1 | 2 | 2 | 2 | 0 | 0 | 0 | 99 | 99 | 99 | 99 | 99 | 99 |
| 3073917 | 0 | 2 | 3 | 2 | 4 | 1 | 1 | 2 | 2 | 1 | 0 | 0 | 0 | 99 | 99 | 99 | 99 | 99 | 99 |
| 3073918 | 1 | 4 | 3 | 2 | 4 | 2 | 1 | 1 | 2 | 2 | 0 | 0 | 0 | 99 | 99 | 99 | 99 | 99 | 99 |
| 3073919 | 1 | 4 | 3 | 2 | 4 | 2 | 1 | 2 | 2 | 1 | 0 | 0 | 0 | 99 | 99 | 99 | 99 | 99 | 99 |
| 3073920 | 0 | 2 | 3 | 2 | 4 | 1 | 2 | 1 | 2 | 2 | 0 | 0 | 0 | 99 | 99 | 99 | 99 | 99 | 99 |
| 3073921 | 1 | 2 | 3 | 2 | 4 | 2 | 1 | 1 | 2 | 2 | 0 | 0 | 1 | 1 | 3 | 1 | 1 | 0 | 1 |
| 3073922 | 1 | 5 | 3 | 2 | 4 | 2 | 1 | 2 | 2 | 2 | 0 | 0 | 1 | 1 | 2 | 1 | 1 | 0 | 1 |
| 3073923 | 1 | 2 | 3 | 2 | 4 | 2 | 1 | 1 | 2 | 2 | 0 | 0 | 0 | 99 | 99 | 99 | 99 | 99 | 99 |
| 3073924 | 0 | 2 | 3 | 2 | 4 | 1 | 2 | 2 | 2 | 2 | 0 | 0 | 0 | 99 | 99 | 99 | 99 | 99 | 99 |
| 3073925 | 1 | 5 | 3 | 2 | 4 | 2 | 1 | 1 | 2 | 2 | 0 | 0 | 0 | 99 | 99 | 99 | 99 | 99 | 99 |

| | | | | | | | | | | | | | | | | | | | |
|---|---|---|---|---|---|---|---|---|---|---|---|---|---|---|---|---|---|---|---|
| 3073926 | 0 | 2 | 3 | 2 | 4 | 1 | 1 | 1 | 2 | 2 | 0 | 0 | 1 | 1 | 2 | 1 | 1 | 0 | 1 |
| 3073927 | 0 | 2 | 3 | 2 | 4 | 1 | 2 | 2 | 2 | 2 | 0 | 0 | 1 | 1 | 2 | 6 | 1 | 1 | 1 |
| 3073928 | 0 | 3 | 3 | 2 | 4 | 1 | 2 | 1 | 4 | 2 | 0 | 0 | 0 | 99 | 99 | 99 | 99 | 99 | 99 |
| 3073929 | 0 | 3 | 3 | 2 | 4 | 2 | 1 | 1 | 2 | 2 | 0 | 0 | 0 | 99 | 99 | 99 | 99 | 99 | 99 |
| 3073930 | 1 | 2 | 3 | 4 | 4 | 5 | 2 | 2 | 2 | 3 | 0 | 0 | 0 | 99 | 99 | 99 | 99 | 99 | 99 |
| 307401 | 0 | 3 | 3 | 3 | 4 | 2 | 2 | 1 | 2 | 4 | 0 | 0 | 1 | 1 | 3 | 1 | 1 | 0 | 1 |
| 307402 | 1 | 3 | 3 | 2 | 4 | 2 | 1 | 1 | 2 | 4 | 0 | 0 | 1 | 1 | 3 | 1 | 1 | 0 | 1 |
| 307403 | 0 | 2 | 3 | 2 | 4 | 2 | 1 | 2 | 2 | 4 | 0 | 0 | 1 | 1 | 1 | 4 | 0 | 1 | 1 |
| 307404 | 1 | 3 | 3 | 3 | 4 | 2 | 2 | 2 | 2 | 4 | 0 | 0 | 1 | 1 | 1 | 4 | 0 | 1 | 1 |
| 307405 | 1 | 3 | 3 | 2 | 4 | 2 | 2 | 2 | 2 | 4 | 0 | 0 | 1 | 1 | 1 | 4 | 0 | 1 | 1 |
| 307406 | 1 | 3 | 3 | 3 | 4 | 2 | 2 | 2 | 2 | 4 | 0 | 0 | 1 | 1 | 1 | 4 | 0 | 1 | 1 |
| 307407 | 0 | 2 | 3 | 2 | 4 | 2 | 2 | 2 | 2 | 4 | 0 | 0 | 1 | 1 | 3 | 1 | 1 | 0 | 1 |
| 307408 | 0 | 1 | 3 | 4 | 4 | 2 | 2 | 2 | 2 | 4 | 0 | 0 | 1 | 1 | 3 | 1 | 1 | 0 | 1 |
| 307409 | 0 | 3 | 3 | 3 | 4 | 2 | 2 | 2 | 2 | 4 | 0 | 0 | 1 | 1 | 1 | 4 | 0 | 1 | 1 |
| 3074010 | 1 | 1 | 3 | 5 | 4 | 5 | 2 | 1 | 2 | 4 | 0 | 0 | 1 | 1 | 1 | 1 | 0 | 0 | 0 |
| 3074011 | 1 | 3 | 3 | 2 | 4 | 2 | 1 | 1 | 2 | 1 | 0 | 0 | 1 | 1 | 3 | 1 | 1 | 0 | 1 |
| 3074012 | 1 | 2 | 3 | 5 | 4 | 5 | 3 | 1 | 2 | 1 | 1 | 0 | 1 | 1 | 3 | 1 | 1 | 0 | 1 |
| 3074013 | 0 | 1 | 3 | 4 | 4 | 2 | 2 | 1 | 2 | 1 | 0 | 0 | 1 | 1 | 3 | 1 | 1 | 0 | 1 |
| 3074014 | 0 | 2 | 3 | 3 | 4 | 2 | 2 | 2 | 2 | 1 | 0 | 0 | 1 | 1 | 1 | 4 | 0 | 1 | 1 |
| 3074015 | 0 | 2 | 3 | 4 | 4 | 2 | 2 | 1 | 2 | 1 | 0 | 0 | 1 | 1 | 1 | 4 | 0 | 1 | 1 |
| 3074016 | 1 | 2 | 3 | 5 | 4 | 5 | 3 | 2 | 2 | 1 | 1 | 1 | 1 | 1 | 1 | 1 | 0 | 0 | 0 |
| 3074017 | 0 | 2 | 3 | 2 | 4 | 2 | 1 | 2 | 2 | 1 | 0 | 0 | 1 | 1 | 1 | 4 | 0 | 1 | 1 |
| 3074018 | 1 | 2 | 3 | 2 | 4 | 2 | 1 | 1 | 2 | 3 | 0 | 0 | 1 | 1 | 1 | 4 | 0 | 1 | 1 |
| 3074019 | 1 | 1 | 3 | 3 | 4 | 2 | 1 | 2 | 2 | 3 | 0 | 0 | 1 | 1 | 1 | 1 | 0 | 0 | 0 |
| 3074020 | 1 | 1 | 3 | 2 | 4 | 2 | 1 | 1 | 2 | 1 | 0 | 0 | 1 | 1 | 1 | 4 | 0 | 1 | 1 |
| 3074021 | 0 | 2 | 3 | 3 | 4 | 2 | 2 | 2 | 2 | 4 | 1 | 0 | 1 | 1 | 1 | 4 | 0 | 1 | 1 |
| 3074022 | 0 | 2 | 3 | 2 | 4 | 2 | 1 | 2 | 2 | 3 | 0 | 0 | 1 | 1 | 1 | 1 | 0 | 0 | 0 |
| 3074023 | 0 | 2 | 3 | 3 | 4 | 2 | 2 | 2 | 2 | 3 | 0 | 0 | 1 | 1 | 1 | 1 | 0 | 0 | 0 |
| 3074024 | 1 | 5 | 3 | 2 | 4 | 2 | 2 | 2 | 2 | 3 | 0 | 0 | 1 | 1 | 1 | 1 | 0 | 0 | 0 |
| 3074025 | 0 | 3 | 3 | 2 | 4 | 2 | 1 | 2 | 2 | 3 | 0 | 0 | 1 | 1 | 1 | 1 | 0 | 0 | 0 |
| 3074026 | 0 | 2 | 3 | 4 | 4 | 2 | 2 | 1 | 2 | 3 | 0 | 0 | 1 | 1 | 3 | 1 | 1 | 0 | 1 |
| 3074027 | 1 | 1 | 3 | 3 | 4 | 2 | 2 | 1 | 2 | 3 | 0 | 0 | 1 | 1 | 1 | 1 | 0 | 0 | 0 |
| 3074028 | 0 | 3 | 3 | 2 | 4 | 2 | 2 | 2 | 2 | 3 | 0 | 0 | 1 | 1 | 1 | 4 | 0 | 1 | 1 |
| 3074029 | 1 | 2 | 3 | 3 | 4 | 2 | 2 | 2 | 2 | 3 | 0 | 0 | 1 | 1 | 1 | 4 | 0 | 1 | 1 |
| 3074030 | 1 | 3 | 3 | 3 | 4 | 2 | 1 | 1 | 2 | 1 | 0 | 0 | 1 | 1 | 1 | 4 | 0 | 1 | 1 |
| 3074031 | 1 | 2 | 3 | 2 | 4 | 2 | 2 | 2 | 2 | 2 | 0 | 0 | 1 | 1 | 1 | 1 | 0 | 0 | 0 |
| 3074032 | 0 | 1 | 3 | 4 | 4 | 2 | 2 | 1 | 2 | 1 | 0 | 0 | 1 | 1 | 1 | 1 | 0 | 0 | 0 |
| 3074033 | 0 | 1 | 3 | 5 | 4 | 5 | 2 | 1 | 2 | 1 | 0 | 0 | 1 | 1 | 1 | 1 | 0 | 0 | 0 |
| 3074034 | 1 | 4 | 3 | 4 | 4 | 2 | 1 | 1 | 2 | 1 | 0 | 0 | 1 | 1 | 3 | 1 | 1 | 0 | 1 |
| 3074035 | 0 | 2 | 3 | 5 | 4 | 5 | 2 | 1 | 2 | 1 | 1 | 0 | 1 | 1 | 3 | 1 | 1 | 0 | 1 |
| 307411 | 0 | 2 | 3 | 2 | 2 | 1 | 2 | 2 | 3 | 2 | 0 | 0 | 1 | 1 | 1 | 1 | 0 | 0 | 0 |
| 307412 | 0 | 4 | 3 | 2 | 4 | 1 | 2 | 1 | 3 | 2 | 0 | 0 | 1 | 1 | 1 | 1 | 0 | 0 | 0 |
| 307413 | 0 | 3 | 3 | 2 | 4 | 1 | 2 | 2 | 3 | 3 | 0 | 0 | 1 | 1 | 1 | 1 | 0 | 0 | 0 |
| 307414 | 0 | 1 | 3 | 4 | 4 | 1 | 1 | 2 | 3 | 3 | 0 | 0 | 1 | 1 | 1 | 1 | 0 | 0 | 0 |
| 307415 | 0 | 2 | 3 | 2 | 4 | 1 | 2 | 2 | 2 | 2 | 0 | 0 | 1 | 1 | 1 | 1 | 0 | 0 | 0 |
| 307416 | 1 | 3 | 3 | 2 | 4 | 2 | 2 | 2 | 2 | 2 | 0 | 0 | 1 | 1 | 2 | 1 | 1 | 0 | 1 |
| 307417 | 0 | 1 | 3 | 2 | 4 | 1 | 2 | 2 | 3 | 2 | 0 | 0 | 1 | 1 | 1 | 1 | 0 | 0 | 0 |
| 307418 | 0 | 2 | 3 | 4 | 4 | 3 | 2 | 1 | 3 | 3 | 0 | 0 | 1 | 1 | 2 | 1 | 1 | 0 | 1 |
| 307419 | 0 | 2 | 3 | 5 | 2 | 5 | 2 | 2 | 3 | 3 | 0 | 0 | 1 | 1 | 2 | 1 | 1 | 0 | 1 |
| 3074110 | 0 | 2 | 3 | 2 | 4 | 1 | 2 | 2 | 3 | 2 | 0 | 0 | 1 | 1 | 2 | 1 | 1 | 0 | 1 |
| 3074111 | 1 | 1 | 3 | 3 | 4 | 2 | 2 | 2 | 3 | 2 | 0 | 0 | 1 | 1 | 2 | 1 | 1 | 0 | 1 |
| 3074112 | 0 | 2 | 3 | 4 | 4 | 1 | 1 | 2 | 3 | 2 | 0 | 0 | 1 | 1 | 2 | 1 | 1 | 0 | 1 |
| 3074113 | 0 | 2 | 3 | 2 | 4 | 1 | 2 | 2 | 3 | 2 | 0 | 0 | 1 | 1 | 1 | 1 | 0 | 0 | 0 |
| 3074114 | 1 | 2 | 3 | 3 | 4 | 2 | 2 | 1 | 3 | 2 | 0 | 0 | 1 | 1 | 1 | 1 | 0 | 0 | 0 |
| 3074115 | 1 | 3 | 3 | 3 | 4 | 4 | 2 | 1 | 3 | 2 | 0 | 0 | 1 | 1 | 1 | 1 | 0 | 0 | 0 |
| 3074116 | 1 | 3 | 3 | 2 | 4 | 3 | 3 | 2 | 3 | 2 | 0 | 0 | 1 | 1 | 1 | 1 | 0 | 0 | 0 |

| | | | | | | | | | | | | | | | | | | | |
|---|---|---|---|---|---|---|---|---|---|---|---|---|---|---|---|---|---|---|---|
| 3074117 | 1 | 3 | 3 | 2 | 4 | 2 | 2 | 2 | 3 | 2 | 0 | 0 | 1 | 1 | 1 | 1 | 0 | 0 | 0 |
| 3074118 | 0 | 3 | 3 | 3 | 4 | 1 | 2 | 2 | 3 | 2 | 0 | 0 | 1 | 1 | 1 | 1 | 0 | 0 | 0 |
| 3074119 | 0 | 4 | 3 | 5 | 4 | 5 | 2 | 1 | 3 | 2 | 0 | 0 | 1 | 1 | 1 | 1 | 0 | 0 | 0 |
| 3074120 | 1 | 5 | 3 | 5 | 4 | 5 | 3 | 2 | 3 | 1 | 1 | 1 | 1 | 1 | 1 | 1 | 0 | 0 | 0 |
| 3074121 | 1 | 4 | 3 | 3 | 4 | 2 | 2 | 1 | 3 | 3 | 0 | 0 | 1 | 1 | 1 | 1 | 0 | 0 | 0 |
| 3074122 | 1 | 3 | 3 | 3 | 4 | 5 | 2 | 1 | 3 | 1 | 0 | 0 | 1 | 1 | 1 | 1 | 0 | 0 | 0 |
| 3074123 | 0 | 4 | 3 | 3 | 4 | 1 | 2 | 1 | 3 | 1 | 0 | 0 | 1 | 1 | 1 | 1 | 0 | 0 | 0 |
| 3074124 | 0 | 5 | 3 | 2 | 4 | 1 | 1 | 1 | 3 | 1 | 0 | 0 | 1 | 1 | 1 | 1 | 0 | 0 | 0 |
| 3074125 | 1 | 5 | 3 | 2 | 4 | 2 | 2 | 2 | 3 | 1 | 0 | 0 | 1 | 1 | 1 | 1 | 0 | 0 | 0 |
| 3074126 | 0 | 2 | 3 | 5 | 4 | 5 | 2 | 1 | 3 | 1 | 0 | 0 | 1 | 1 | 1 | 1 | 0 | 0 | 0 |
| 3074127 | 0 | 5 | 3 | 2 | 4 | 1 | 2 | 2 | 3 | 1 | 0 | 0 | 1 | 1 | 1 | 1 | 0 | 0 | 0 |
| 3074128 | 0 | 5 | 3 | 2 | 4 | 1 | 2 | 2 | 3 | 3 | 0 | 0 | 1 | 1 | 1 | 1 | 0 | 0 | 0 |
| 3074129 | 1 | 4 | 3 | 5 | 4 | 5 | 2 | 2 | 3 | 3 | 1 | 0 | 1 | 1 | 4 | 1 | 1 | 0 | 1 |
| 3074130 | 0 | 2 | 3 | 3 | 4 | 1 | 3 | 2 | 3 | 2 | 1 | 0 | 1 | 1 | 1 | 1 | 0 | 0 | 0 |
| 3074131 | 1 | 5 | 3 | 2 | 4 | 2 | 2 | 2 | 3 | 2 | 0 | 0 | 1 | 1 | 2 | 1 | 1 | 0 | 1 |
| 3074132 | 0 | 2 | 3 | 2 | 4 | 1 | 2 | 2 | 4 | 4 | 0 | 0 | 0 | 99 | 99 | 99 | 99 | 99 | 99 |
| 3074133 | 1 | 4 | 3 | 2 | 4 | 2 | 2 | 1 | 4 | 4 | 0 | 0 | 0 | 99 | 99 | 99 | 99 | 99 | 99 |
| 3074134 | 0 | 2 | 3 | 5 | 4 | 5 | 3 | 1 | 4 | 3 | 1 | 1 | 1 | 1 | 1 | 1 | 0 | 0 | 0 |
| 3074135 | 1 | 5 | 3 | 2 | 4 | 2 | 2 | 1 | 4 | 3 | 0 | 0 | 1 | 1 | 2 | 1 | 1 | 0 | 1 |
| 3074136 | 1 | 5 | 3 | 2 | 4 | 2 | 2 | 1 | 4 | 4 | 0 | 0 | 1 | 1 | 1 | 1 | 0 | 0 | 0 |
| 3074137 | 1 | 5 | 3 | 2 | 4 | 2 | 2 | 2 | 4 | 3 | 0 | 0 | 0 | 99 | 99 | 99 | 99 | 99 | 99 |
| 3074138 | 0 | 3 | 3 | 3 | 4 | 1 | 2 | 1 | 4 | 4 | 0 | 0 | 1 | 1 | 2 | 1 | 1 | 0 | 1 |
| 3074139 | 1 | 3 | 3 | 2 | 4 | 4 | 2 | 1 | 4 | 3 | 0 | 0 | 0 | 99 | 99 | 99 | 99 | 99 | 99 |
| 3074140 | 1 | 5 | 3 | 2 | 4 | 2 | 2 | 2 | 4 | 4 | 0 | 0 | 0 | 99 | 99 | 99 | 99 | 99 | 99 |
| 308421 | 0 | 4 | 3 | 2 | 4 | 2 | 1 | 1 | 4 | 4 | 0 | 0 | 0 | 99 | 99 | 99 | 99 | 99 | 99 |
| 308422 | 1 | 4 | 3 | 2 | 4 | 2 | 2 | 2 | 4 | 4 | 0 | 0 | 1 | 1 | 4 | 1 | 1 | 0 | 1 |
| 308423 | 0 | 3 | 3 | 3 | 4 | 2 | 3 | 2 | 2 | 4 | 0 | 0 | 1 | 1 | 4 | 1 | 1 | 0 | 1 |
| 308424 | 0 | 2 | 3 | 2 | 4 | 2 | 1 | 1 | 2 | 4 | 0 | 0 | 1 | 1 | 4 | 1 | 1 | 0 | 1 |
| 308425 | 0 | 3 | 3 | 2 | 4 | 2 | 3 | 2 | 2 | 4 | 0 | 0 | 1 | 1 | 1 | 1 | 0 | 0 | 0 |
| 308426 | 0 | 2 | 3 | 2 | 4 | 2 | 3 | 2 | 2 | 4 | 0 | 0 | 1 | 1 | 1 | 1 | 0 | 0 | 0 |
| 308427 | 1 | 2 | 3 | 2 | 4 | 2 | 1 | 2 | 4 | 4 | 0 | 0 | 0 | 99 | 99 | 99 | 99 | 99 | 99 |
| 308428 | 1 | 4 | 3 | 2 | 4 | 2 | 2 | 2 | 4 | 4 | 0 | 0 | 1 | 1 | 2 | 1 | 1 | 0 | 1 |
| 308429 | 0 | 1 | 3 | 4 | 4 | 2 | 2 | 2 | 4 | 4 | 0 | 0 | 1 | 1 | 3 | 1 | 1 | 0 | 1 |
| 3084210 | 0 | 2 | 3 | 2 | 4 | 2 | 2 | 1 | 2 | 4 | 0 | 0 | 0 | 99 | 99 | 99 | 99 | 99 | 99 |
| 3084211 | 1 | 3 | 3 | 2 | 4 | 2 | 1 | 2 | 2 | 3 | 0 | 0 | 0 | 99 | 99 | 99 | 99 | 99 | 99 |
| 3084212 | 0 | 3 | 3 | 1 | 4 | 2 | 2 | 1 | 2 | 1 | 0 | 0 | 0 | 99 | 99 | 99 | 99 | 99 | 99 |
| 3084213 | 0 | 2 | 3 | 2 | 4 | 2 | 2 | 2 | 4 | 4 | 0 | 0 | 0 | 99 | 99 | 99 | 99 | 99 | 99 |
| 3084214 | 0 | 2 | 3 | 2 | 4 | 2 | 2 | 1 | 4 | 4 | 0 | 0 | 0 | 99 | 99 | 99 | 99 | 99 | 99 |
| 3084215 | 1 | 2 | 3 | 2 | 4 | 2 | 1 | 1 | 2 | 4 | 0 | 0 | 0 | 99 | 99 | 99 | 99 | 99 | 99 |
| 3084216 | 1 | 3 | 3 | 2 | 4 | 2 | 2 | 2 | 2 | 4 | 0 | 0 | 0 | 99 | 99 | 99 | 99 | 99 | 99 |
| 3084217 | 1 | 2 | 3 | 2 | 4 | 2 | 2 | 1 | 2 | 1 | 0 | 0 | 0 | 99 | 99 | 99 | 99 | 99 | 99 |
| 3084218 | 1 | 2 | 3 | 2 | 4 | 2 | 2 | 1 | 4 | 1 | 0 | 0 | 1 | 1 | 3 | 1 | 1 | 0 | 1 |
| 3084219 | 1 | 3 | 3 | 3 | 4 | 2 | 2 | 1 | 4 | 1 | 0 | 0 | 1 | 1 | 1 | 1 | 0 | 0 | 0 |
| 3084220 | 1 | 4 | 3 | 4 | 4 | 5 | 3 | 2 | 4 | 1 | 1 | 1 | 0 | 99 | 99 | 99 | 99 | 99 | 99 |
| 3084221 | 0 | 2 | 3 | 2 | 4 | 3 | 2 | 2 | 4 | 1 | 1 | 1 | 1 | 1 | 1 | 1 | 0 | 0 | 0 |
| 3084222 | 0 | 4 | 3 | 3 | 4 | 1 | 2 | 2 | 4 | 1 | 0 | 0 | 0 | 99 | 99 | 99 | 99 | 99 | 99 |
| 3084223 | 0 | 4 | 3 | 2 | 4 | 1 | 1 | 1 | 4 | 1 | 0 | 0 | 1 | 1 | 1 | 1 | 0 | 0 | 0 |
| 3084224 | 0 | 2 | 3 | 3 | 4 | 1 | 2 | 2 | 4 | 1 | 0 | 0 | 0 | 99 | 99 | 99 | 99 | 99 | 99 |
| 3084225 | 1 | 3 | 3 | 2 | 4 | 2 | 2 | 2 | 4 | 2 | 0 | 0 | 0 | 99 | 99 | 99 | 99 | 99 | 99 |
| 3084226 | 0 | 2 | 3 | 4 | 4 | 1 | 3 | 2 | 4 | 3 | 0 | 0 | 1 | 1 | 1 | 1 | 0 | 0 | 0 |
| 3084227 | 1 | 4 | 3 | 2 | 4 | 2 | 2 | 2 | 4 | 3 | 0 | 0 | 0 | 99 | 99 | 99 | 99 | 99 | 99 |
| 3084228 | 1 | 5 | 3 | 1 | 4 | 2 | 2 | 1 | 4 | 2 | 0 | 0 | 0 | 99 | 99 | 99 | 99 | 99 | 99 |
| 3084229 | 0 | 1 | 3 | 3 | 4 | 1 | 3 | 2 | 4 | 2 | 0 | 0 | 1 | 1 | 1 | 1 | 0 | 0 | 0 |
| 3084230 | 1 | 4 | 3 | 2 | 4 | 5 | 2 | 2 | 4 | 1 | 1 | 1 | 0 | 99 | 99 | 99 | 99 | 99 | 99 |
| 308431 | 0 | 3 | 3 | 2 | 4 | 1 | 2 | 2 | 2 | 4 | 0 | 0 | 1 | 1 | 1 | 1 | 0 | 0 | 0 |
| 308432 | 1 | 5 | 3 | 4 | 4 | 5 | 2 | 1 | 2 | 4 | 0 | 0 | 1 | 1 | 2 | 1 | 1 | 0 | 1 |

| | | | | | | | | | | | | | | | | | | | |
|---|---|---|---|---|---|---|---|---|---|---|---|---|---|---|---|---|---|---|---|
| 308433 | 0 | 2 | 3 | 2 | 4 | 2 | 1 | 1 | 2 | 4 | 0 | 0 | 1 | 1 | 2 | 1 | 1 | 0 | 1 |
| 308434 | 0 | 4 | 3 | 2 | 4 | 2 | 2 | 1 | 2 | 4 | 0 | 0 | 0 | 99 | 99 | 99 | 99 | 99 | 99 |
| 308435 | 0 | 2 | 3 | 4 | 4 | 2 | 1 | 1 | 2 | 4 | 0 | 0 | 1 | 1 | 1 | 1 | 0 | 0 | 0 |
| 308436 | 1 | 1 | 3 | 4 | 4 | 5 | 2 | 1 | 2 | 4 | 0 | 0 | 1 | 1 | 2 | 1 | 1 | 0 | 1 |
| 308437 | 0 | 1 | 3 | 5 | 4 | 5 | 2 | 2 | 2 | 4 | 1 | 0 | 0 | 99 | 99 | 99 | 99 | 99 | 99 |
| 308438 | 1 | 2 | 3 | 2 | 4 | 3 | 2 | 1 | 2 | 3 | 0 | 0 | 0 | 99 | 99 | 99 | 99 | 99 | 99 |
| 308439 | 0 | 2 | 3 | 2 | 4 | 2 | 2 | 2 | 2 | 4 | 0 | 0 | 0 | 99 | 99 | 99 | 99 | 99 | 99 |
| 3084310 | 1 | 3 | 3 | 3 | 4 | 2 | 1 | 2 | 2 | 3 | 0 | 0 | 1 | 1 | 1 | 1 | 0 | 0 | 0 |
| 3084311 | 0 | 2 | 3 | 2 | 4 | 2 | 2 | 2 | 2 | 4 | 0 | 0 | 1 | 1 | 3 | 1 | 1 | 0 | 1 |
| 3084312 | 0 | 2 | 3 | 2 | 2 | 1 | 2 | 2 | 4 | 1 | 0 | 0 | 0 | 99 | 99 | 99 | 99 | 99 | 99 |
| 3084313 | 1 | 5 | 3 | 2 | 4 | 2 | 2 | 1 | 4 | 2 | 0 | 0 | 0 | 99 | 99 | 99 | 99 | 99 | 99 |
| 3084314 | 0 | 1 | 3 | 4 | 4 | 1 | 2 | 2 | 4 | 4 | 0 | 0 | 0 | 99 | 99 | 99 | 99 | 99 | 99 |
| 3084315 | 1 | 4 | 3 | 4 | 4 | 3 | 3 | 2 | 4 | 2 | 0 | 0 | 0 | 99 | 99 | 99 | 99 | 99 | 99 |
| 3084316 | 1 | 1 | 3 | 2 | 4 | 2 | 3 | 1 | 4 | 4 | 0 | 0 | 1 | 1 | 1 | 6 | 0 | 1 | 1 |
| 3084317 | 1 | 1 | 3 | 4 | 2 | 2 | 2 | 2 | 4 | 3 | 0 | 0 | 1 | 1 | 1 | 6 | 0 | 1 | 1 |
| 3084318 | 0 | 1 | 3 | 2 | 4 | 1 | 2 | 1 | 4 | 4 | 0 | 0 | 0 | 99 | 99 | 99 | 99 | 99 | 99 |
| 3084319 | 0 | 2 | 3 | 2 | 4 | 1 | 2 | 2 | 4 | 4 | 0 | 0 | 0 | 99 | 99 | 99 | 99 | 99 | 99 |
| 3084320 | 1 | 2 | 3 | 3 | 4 | 2 | 3 | 2 | 4 | 4 | 0 | 0 | 0 | 99 | 99 | 99 | 99 | 99 | 99 |
| 3084321 | 1 | 5 | 3 | 2 | 4 | 2 | 2 | 2 | 4 | 2 | 0 | 0 | 1 | 1 | 4 | 1 | 1 | 0 | 1 |
| 3084322 | 0 | 3 | 3 | 2 | 4 | 3 | 3 | 2 | 4 | 2 | 0 | 0 | 0 | 99 | 99 | 99 | 99 | 99 | 99 |
| 3084323 | 0 | 3 | 3 | 2 | 4 | 2 | 2 | 2 | 4 | 3 | 0 | 0 | 0 | 99 | 99 | 99 | 99 | 99 | 99 |
| 3084324 | 1 | 1 | 3 | 2 | 4 | 2 | 3 | 1 | 4 | 4 | 0 | 0 | 0 | 99 | 99 | 99 | 99 | 99 | 99 |
| 3084325 | 0 | 3 | 3 | 4 | 4 | 1 | 3 | 2 | 4 | 1 | 0 | 0 | 0 | 99 | 99 | 99 | 99 | 99 | 99 |
| 3084326 | 0 | 1 | 3 | 2 | 4 | 2 | 1 | 1 | 4 | 4 | 0 | 0 | 0 | 99 | 99 | 99 | 99 | 99 | 99 |
| 3084327 | 0 | 1 | 3 | 2 | 4 | 2 | 1 | 1 | 4 | 4 | 0 | 0 | 0 | 99 | 99 | 99 | 99 | 99 | 99 |
| 3084328 | 0 | 1 | 3 | 5 | 4 | 5 | 2 | 2 | 4 | 4 | 0 | 0 | 0 | 99 | 99 | 99 | 99 | 99 | 99 |
| 3084329 | 1 | 5 | 3 | 5 | 4 | 5 | 3 | 2 | 4 | 1 | 1 | 1 | 1 | 1 | 2 | 1 | 1 | 0 | 1 |
| 3084330 | 0 | 2 | 3 | 2 | 4 | 2 | 2 | 1 | 4 | 4 | 0 | 0 | 0 | 99 | 99 | 99 | 99 | 99 | 99 |
| 3084331 | 1 | 4 | 3 | 4 | 4 | 2 | 2 | 1 | 4 | 4 | 0 | 0 | 1 | 1 | 4 | 1 | 1 | 0 | 1 |
| 3084332 | 1 | 2 | 3 | 5 | 4 | 5 | 2 | 1 | 4 | 3 | 1 | 1 | 1 | 1 | 1 | 6 | 0 | 1 | 1 |
| 3084333 | 0 | 2 | 3 | 5 | 4 | 5 | 3 | 1 | 4 | 4 | 1 | 1 | 0 | 99 | 99 | 99 | 99 | 99 | 99 |
| 3084334 | 1 | 4 | 3 | 5 | 4 | 5 | 3 | 2 | 4 | 4 | 1 | 1 | 0 | 99 | 99 | 99 | 99 | 99 | 99 |
| 3084335 | 1 | 4 | 3 | 5 | 4 | 5 | 3 | 1 | 4 | 3 | 0 | 0 | 1 | 1 | 1 | 1 | 0 | 0 | 0 |
| 3084336 | 1 | 4 | 3 | 5 | 3 | 5 | 3 | 2 | 4 | 4 | 1 | 1 | 1 | 1 | 1 | 1 | 0 | 0 | 0 |
| 3084337 | 0 | 4 | 3 | 2 | 4 | 1 | 3 | 1 | 4 | 4 | 0 | 0 | 1 | 1 | 1 | 1 | 0 | 0 | 0 |
| 3084338 | 1 | 3 | 3 | 4 | 4 | 3 | 3 | 2 | 4 | 3 | 1 | 0 | 0 | 99 | 99 | 99 | 99 | 99 | 99 |
| 3084339 | 1 | 4 | 3 | 3 | 4 | 2 | 3 | 1 | 4 | 4 | 0 | 0 | 1 | 1 | 3 | 1 | 1 | 0 | 1 |
| 308441 | 0 | 2 | 3 | 3 | 4 | 2 | 2 | 1 | 2 | 4 | 0 | 0 | 1 | 1 | 2 | 1 | 1 | 0 | 1 |
| 308442 | 0 | 2 | 3 | 2 | 4 | 2 | 2 | 2 | 2 | 4 | 0 | 0 | 0 | 99 | 99 | 99 | 99 | 99 | 99 |
| 308443 | 0 | 2 | 3 | 2 | 4 | 2 | 2 | 2 | 2 | 1 | 0 | 0 | 0 | 99 | 99 | 99 | 99 | 99 | 99 |
| 308444 | 0 | 5 | 3 | 2 | 4 | 2 | 2 | 1 | 2 | 1 | 0 | 0 | 1 | 1 | 1 | 1 | 0 | 0 | 0 |
| 308445 | 0 | 5 | 3 | 2 | 4 | 2 | 2 | 1 | 2 | 1 | 0 | 0 | 1 | 1 | 3 | 1 | 1 | 0 | 1 |
| 308446 | 1 | 4 | 3 | 2 | 4 | 2 | 2 | 2 | 2 | 1 | 0 | 0 | 1 | 1 | 3 | 1 | 1 | 0 | 1 |
| 308447 | 1 | 5 | 3 | 2 | 4 | 2 | 2 | 1 | 2 | 3 | 1 | 1 | 0 | 99 | 99 | 99 | 99 | 99 | 99 |
| 308448 | 1 | 5 | 3 | 2 | 4 | 2 | 2 | 2 | 2 | 4 | 0 | 0 | 0 | 99 | 99 | 99 | 99 | 99 | 99 |
| 308449 | 0 | 1 | 3 | 2 | 4 | 2 | 1 | 1 | 2 | 4 | 0 | 0 | 1 | 1 | 3 | 1 | 1 | 0 | 1 |
| 3084410 | 1 | 3 | 3 | 3 | 4 | 2 | 2 | 1 | 2 | 4 | 0 | 0 | 0 | 99 | 99 | 99 | 99 | 99 | 99 |
| 3084411 | 1 | 5 | 3 | 1 | 4 | 2 | 2 | 2 | 2 | 2 | 0 | 0 | 0 | 99 | 99 | 99 | 99 | 99 | 99 |
| 3084412 | 1 | 3 | 3 | 4 | 4 | 3 | 3 | 1 | 2 | 1 | 1 | 0 | 1 | 1 | 4 | 6 | 1 | 1 | 1 |
| 3084413 | 1 | 5 | 3 | 2 | 4 | 2 | 2 | 1 | 2 | 1 | 0 | 0 | 1 | 1 | 4 | 1 | 1 | 0 | 1 |
| 3084414 | 1 | 3 | 3 | 4 | 4 | 5 | 3 | 2 | 2 | 1 | 0 | 0 | 1 | 1 | 1 | 1 | 0 | 0 | 0 |
| 3084415 | 1 | 1 | 3 | 2 | 4 | 4 | 3 | 2 | 2 | 1 | 0 | 0 | 1 | 1 | 3 | 1 | 1 | 0 | 1 |
| 3084416 | 0 | 3 | 3 | 2 | 4 | 2 | 2 | 1 | 2 | 1 | 0 | 0 | 0 | 99 | 99 | 99 | 99 | 99 | 99 |
| 3084417 | 0 | 1 | 3 | 5 | 4 | 5 | 3 | 2 | 2 | 3 | 0 | 0 | 1 | 1 | 3 | 1 | 1 | 0 | 1 |
| 3084418 | 1 | 2 | 3 | 3 | 4 | 2 | 3 | 1 | 2 | 3 | 1 | 1 | 0 | 99 | 99 | 99 | 99 | 99 | 99 |
| 3084419 | 1 | 1 | 3 | 4 | 4 | 2 | 2 | 1 | 2 | 3 | 0 | 0 | 1 | 1 | 3 | 1 | 1 | 0 | 1 |

| | | | | | | | | | | | | | | | | | | | |
|---|---|---|---|---|---|---|---|---|---|---|---|---|---|---|---|---|---|---|---|
| 3084420 | 1 | 5 | 3 | 2 | 4 | 2 | 1 | 2 | 2 | 1 | 0 | 0 | 0 | 99 | 99 | 99 | 99 | 99 | 99 |
| 3084421 | 0 | 2 | 3 | 3 | 4 | 1 | 3 | 1 | 2 | 4 | 0 | 0 | 0 | 99 | 99 | 99 | 99 | 99 | 99 |
| 3084422 | 0 | 2 | 3 | 2 | 4 | 2 | 2 | 2 | 2 | 4 | 0 | 0 | 0 | 99 | 99 | 99 | 99 | 99 | 99 |
| 3084423 | 1 | 2 | 3 | 5 | 4 | 5 | 3 | 1 | 2 | 4 | 0 | 0 | 1 | 1 | 1 | 1 | 0 | 0 | 0 |
| 3084424 | 1 | 1 | 3 | 5 | 4 | 5 | 3 | 1 | 2 | 4 | 0 | 0 | 1 | 1 | 1 | 6 | 0 | 1 | 1 |
| 3084425 | 1 | 2 | 3 | 4 | 4 | 5 | 3 | 2 | 2 | 4 | 1 | 0 | 0 | 99 | 99 | 99 | 99 | 99 | 99 |
| 3084426 | 0 | 1 | 3 | 5 | 4 | 5 | 3 | 1 | 2 | 3 | 1 | 0 | 1 | 1 | 1 | 1 | 0 | 0 | 0 |
| 308451 | 0 | 4 | 3 | 1 | 4 | 2 | 2 | 2 | 3 | 2 | 0 | 0 | 1 | 1 | 1 | 1 | 0 | 0 | 0 |
| 308452 | 1 | 2 | 3 | 2 | 4 | 2 | 1 | 2 | 3 | 2 | 0 | 0 | 1 | 1 | 1 | 4 | 0 | 1 | 1 |
| 308453 | 0 | 3 | 3 | 2 | 4 | 2 | 2 | 1 | 3 | 2 | 0 | 0 | 0 | 99 | 99 | 99 | 99 | 99 | 99 |
| 308454 | 0 | 2 | 3 | 2 | 4 | 2 | 1 | 2 | 3 | 3 | 0 | 0 | 0 | 99 | 99 | 99 | 99 | 99 | 99 |
| 308455 | 0 | 4 | 3 | 1 | 4 | 2 | 1 | 1 | 3 | 2 | 0 | 0 | 1 | 1 | 1 | 6 | 0 | 1 | 1 |
| 308456 | 1 | 2 | 3 | 2 | 4 | 2 | 2 | 2 | 1 | 2 | 0 | 0 | 1 | 1 | 1 | 1 | 0 | 0 | 0 |
| 308457 | 1 | 2 | 3 | 2 | 4 | 2 | 1 | 2 | 3 | 3 | 0 | 0 | 1 | 1 | 1 | 4 | 0 | 1 | 1 |
| 308458 | 0 | 3 | 3 | 2 | 4 | 2 | 2 | 1 | 3 | 3 | 0 | 0 | 0 | 99 | 99 | 99 | 99 | 99 | 99 |
| 308459 | 0 | 2 | 3 | 1 | 4 | 2 | 1 | 1 | 3 | 2 | 0 | 0 | 1 | 1 | 1 | 4 | 0 | 1 | 1 |
| 3084510 | 0 | 2 | 3 | 2 | 4 | 2 | 2 | 1 | 3 | 2 | 0 | 0 | 1 | 1 | 1 | 4 | 0 | 1 | 1 |
| 3084511 | 1 | 4 | 3 | 1 | 4 | 2 | 1 | 2 | 3 | 2 | 0 | 0 | 1 | 1 | 1 | 4 | 0 | 1 | 1 |
| 3084512 | 0 | 2 | 3 | 2 | 4 | 2 | 1 | 2 | 3 | 2 | 0 | 0 | 1 | 1 | 1 | 4 | 0 | 1 | 1 |
| 3084513 | 0 | 1 | 3 | 2 | 4 | 2 | 1 | 1 | 3 | 2 | 0 | 0 | 1 | 1 | 1 | 4 | 0 | 1 | 1 |
| 3084514 | 0 | 1 | 3 | 2 | 4 | 2 | 1 | 1 | 3 | 2 | 0 | 0 | 1 | 1 | 1 | 4 | 0 | 1 | 1 |
| 3084515 | 1 | 4 | 3 | 1 | 4 | 2 | 1 | 1 | 3 | 2 | 0 | 0 | 1 | 1 | 1 | 4 | 0 | 1 | 1 |
| 3084516 | 1 | 4 | 3 | 2 | 4 | 2 | 1 | 2 | 3 | 2 | 0 | 0 | 1 | 1 | 1 | 4 | 0 | 1 | 1 |
| 3084517 | 0 | 4 | 3 | 2 | 4 | 2 | 1 | 2 | 3 | 2 | 0 | 0 | 1 | 1 | 3 | 4 | 1 | 1 | 1 |
| 3084518 | 1 | 3 | 3 | 2 | 4 | 2 | 1 | 2 | 3 | 2 | 0 | 0 | 1 | 1 | 1 | 4 | 0 | 1 | 1 |
| 3084519 | 0 | 4 | 3 | 1 | 4 | 2 | 1 | 1 | 3 | 2 | 0 | 0 | 1 | 1 | 1 | 4 | 0 | 1 | 1 |
| 3084520 | 0 | 5 | 3 | 2 | 4 | 2 | 1 | 1 | 3 | 2 | 0 | 0 | 1 | 1 | 1 | 4 | 0 | 1 | 1 |
| 3084521 | 0 | 4 | 3 | 2 | 4 | 2 | 1 | 2 | 3 | 2 | 0 | 0 | 1 | 1 | 1 | 4 | 0 | 1 | 1 |
| 3084522 | 0 | 2 | 3 | 2 | 4 | 2 | 2 | 2 | 3 | 2 | 0 | 0 | 1 | 1 | 1 | 6 | 0 | 1 | 1 |
| 3084523 | 0 | 1 | 3 | 2 | 4 | 2 | 1 | 1 | 3 | 2 | 0 | 0 | 1 | 1 | 1 | 6 | 0 | 1 | 1 |
| 3084524 | 1 | 4 | 3 | 2 | 4 | 2 | 1 | 2 | 3 | 2 | 0 | 0 | 1 | 1 | 1 | 1 | 0 | 0 | 0 |
| 3084525 | 1 | 5 | 3 | 2 | 4 | 2 | 1 | 1 | 3 | 2 | 0 | 0 | 1 | 1 | 1 | 4 | 0 | 1 | 1 |
| 308461 | 0 | 4 | 3 | 3 | 4 | 2 | 3 | 2 | 3 | 2 | 1 | 1 | 1 | 1 | 1 | 6 | 0 | 1 | 1 |
| 308462 | 1 | 3 | 3 | 1 | 4 | 2 | 1 | 1 | 3 | 2 | 0 | 0 | 1 | 1 | 1 | 1 | 0 | 0 | 0 |
| 308463 | 1 | 2 | 3 | 2 | 4 | 2 | 2 | 1 | 3 | 2 | 0 | 0 | 1 | 1 | 1 | 4 | 0 | 1 | 1 |
| 308464 | 1 | 4 | 3 | 1 | 4 | 2 | 1 | 2 | 3 | 2 | 0 | 0 | 1 | 1 | 1 | 4 | 0 | 1 | 1 |
| 308465 | 0 | 5 | 3 | 2 | 4 | 2 | 1 | 1 | 3 | 2 | 0 | 0 | 1 | 1 | 1 | 4 | 0 | 1 | 1 |
| 308466 | 1 | 3 | 3 | 2 | 4 | 2 | 1 | 1 | 3 | 2 | 0 | 0 | 1 | 1 | 2 | 1 | 1 | 0 | 1 |
| 308467 | 0 | 4 | 3 | 2 | 4 | 2 | 2 | 1 | 3 | 2 | 0 | 0 | 1 | 1 | 1 | 6 | 0 | 1 | 1 |
| 308468 | 0 | 3 | 3 | 2 | 4 | 2 | 2 | 2 | 3 | 2 | 0 | 0 | 1 | 1 | 1 | 1 | 0 | 0 | 0 |
| 308469 | 0 | 5 | 3 | 2 | 4 | 2 | 1 | 1 | 3 | 2 | 0 | 0 | 1 | 1 | 1 | 4 | 0 | 1 | 1 |
| 3084610 | 1 | 2 | 3 | 2 | 4 | 2 | 1 | 1 | 3 | 2 | 0 | 0 | 1 | 1 | 1 | 1 | 0 | 0 | 0 |
| 3084611 | 1 | 2 | 3 | 2 | 4 | 2 | 1 | 2 | 3 | 2 | 0 | 0 | 1 | 1 | 1 | 4 | 0 | 1 | 1 |
| 3084612 | 0 | 3 | 3 | 2 | 4 | 2 | 2 | 1 | 3 | 2 | 0 | 0 | 1 | 1 | 1 | 6 | 0 | 1 | 1 |
| 3084613 | 0 | 3 | 3 | 4 | 4 | 2 | 2 | 1 | 3 | 2 | 0 | 0 | 1 | 1 | 1 | 1 | 0 | 0 | 0 |
| 3084614 | 1 | 5 | 3 | 2 | 4 | 2 | 1 | 1 | 3 | 2 | 0 | 0 | 1 | 1 | 1 | 4 | 0 | 1 | 1 |
| 3084615 | 1 | 5 | 3 | 2 | 4 | 2 | 2 | 1 | 3 | 2 | 0 | 0 | 1 | 1 | 1 | 4 | 0 | 1 | 1 |
| 3084616 | 1 | 4 | 3 | 4 | 4 | 2 | 1 | 1 | 3 | 2 | 0 | 0 | 1 | 1 | 1 | 4 | 0 | 1 | 1 |
| 3084617 | 0 | 4 | 3 | 2 | 4 | 2 | 1 | 2 | 3 | 2 | 0 | 0 | 1 | 1 | 1 | 4 | 0 | 1 | 1 |
| 3084618 | 1 | 4 | 3 | 2 | 4 | 2 | 1 | 2 | 3 | 2 | 0 | 0 | 1 | 1 | 1 | 4 | 0 | 1 | 1 |
| 3084619 | 1 | 4 | 3 | 2 | 4 | 2 | 1 | 2 | 3 | 2 | 0 | 0 | 1 | 1 | 1 | 4 | 0 | 1 | 1 |
| 3084620 | 1 | 3 | 3 | 2 | 4 | 2 | 1 | 1 | 3 | 2 | 0 | 0 | 1 | 1 | 1 | 4 | 0 | 1 | 1 |
| 3084621 | 0 | 5 | 3 | 1 | 4 | 2 | 1 | 2 | 3 | 2 | 0 | 0 | 1 | 1 | 1 | 4 | 0 | 1 | 1 |
| 3084622 | 1 | 3 | 3 | 2 | 4 | 2 | 1 | 1 | 3 | 2 | 0 | 0 | 1 | 1 | 1 | 4 | 0 | 1 | 1 |
| 3084623 | 1 | 4 | 3 | 2 | 4 | 2 | 1 | 1 | 3 | 2 | 0 | 0 | 1 | 1 | 1 | 4 | 0 | 1 | 1 |
| 3084624 | 1 | 3 | 3 | 2 | 4 | 2 | 1 | 2 | 3 | 2 | 0 | 0 | 1 | 1 | 1 | 4 | 0 | 1 | 1 |

| | | | | | | | | | | | | | | | | | | | |
|---|---|---|---|---|---|---|---|---|---|---|---|---|---|---|---|---|---|---|---|
| 3084625 | 0 | 4 | 3 | 2 | 4 | 2 | 1 | 1 | 3 | 2 | 0 | 0 | 1 | 1 | 1 | 4 | 0 | 1 | 1 |
| 309471 | 1 | 4 | 3 | 4 | 4 | 2 | 2 | 2 | 4 | 2 | 1 | 1 | 1 | 1 | 1 | 1 | 0 | 0 | 0 |
| 309472 | 0 | 3 | 3 | 3 | 4 | 2 | 1 | 1 | 4 | 1 | 0 | 0 | 1 | 1 | 2 | 1 | 1 | 0 | 1 |
| 309473 | 1 | 3 | 3 | 2 | 4 | 2 | 1 | 2 | 4 | 2 | 0 | 0 | 1 | 1 | 1 | 1 | 0 | 0 | 0 |
| 309474 | 1 | 4 | 3 | 2 | 4 | 2 | 1 | 2 | 4 | 1 | 1 | 0 | 1 | 1 | 1 | 1 | 0 | 0 | 0 |
| 309475 | 0 | 2 | 3 | 2 | 4 | 2 | 1 | 1 | 4 | 1 | 1 | 1 | 1 | 1 | 1 | 1 | 0 | 0 | 0 |
| 309476 | 0 | 3 | 3 | 4 | 4 | 1 | 3 | 2 | 4 | 4 | 1 | 1 | 1 | 1 | 1 | 1 | 0 | 0 | 0 |
| 309477 | 1 | 2 | 3 | 2 | 4 | 2 | 2 | 2 | 4 | 1 | 0 | 0 | 1 | 1 | 2 | 1 | 1 | 0 | 1 |
| 309478 | 1 | 4 | 3 | 4 | 4 | 2 | 1 | 2 | 4 | 1 | 1 | 0 | 1 | 1 | 1 | 1 | 0 | 0 | 0 |
| 309479 | 1 | 5 | 3 | 2 | 4 | 2 | 1 | 1 | 4 | 4 | 1 | 0 | 1 | 1 | 2 | 4 | 1 | 1 | 1 |
| 3094710 | 1 | 2 | 3 | 2 | 4 | 2 | 3 | 1 | 4 | 4 | 0 | 0 | 1 | 1 | 2 | 1 | 1 | 0 | 1 |
| 3094711 | 0 | 2 | 3 | 2 | 4 | 2 | 1 | 1 | 4 | 4 | 0 | 0 | 1 | 1 | 2 | 1 | 1 | 0 | 1 |
| 3094712 | 0 | 2 | 3 | 5 | 4 | 5 | 2 | 2 | 4 | 3 | 0 | 0 | 1 | 1 | 1 | 1 | 0 | 0 | 0 |
| 3094713 | 1 | 1 | 3 | 2 | 4 | 2 | 2 | 2 | 4 | 4 | 0 | 0 | 1 | 1 | 2 | 4 | 1 | 1 | 1 |
| 3094714 | 1 | 1 | 3 | 2 | 4 | 2 | 1 | 1 | 4 | 4 | 0 | 0 | 1 | 1 | 3 | 1 | 1 | 0 | 1 |
| 3094715 | 1 | 4 | 3 | 2 | 4 | 2 | 1 | 1 | 4 | 4 | 0 | 0 | 1 | 1 | 1 | 4 | 0 | 1 | 1 |
| 3094716 | 1 | 2 | 3 | 2 | 4 | 2 | 2 | 2 | 4 | 4 | 0 | 0 | 1 | 1 | 3 | 1 | 1 | 0 | 1 |
| 3094717 | 1 | 2 | 3 | 5 | 4 | 5 | 3 | 1 | 4 | 1 | 1 | 0 | 1 | 1 | 1 | 1 | 0 | 0 | 0 |
| 3094718 | 1 | 2 | 3 | 2 | 4 | 2 | 1 | 2 | 4 | 2 | 0 | 0 | 1 | 1 | 3 | 1 | 1 | 0 | 1 |
| 3094719 | 0 | 2 | 3 | 3 | 4 | 2 | 2 | 2 | 4 | 2 | 0 | 0 | 1 | 1 | 2 | 1 | 1 | 0 | 1 |
| 3094720 | 0 | 4 | 3 | 3 | 4 | 2 | 1 | 1 | 4 | 2 | 0 | 0 | 1 | 1 | 2 | 1 | 1 | 0 | 1 |
| 3094721 | 1 | 3 | 3 | 2 | 4 | 2 | 1 | 2 | 4 | 2 | 0 | 0 | 1 | 1 | 2 | 1 | 1 | 0 | 1 |
| 3094722 | 0 | 3 | 3 | 2 | 4 | 2 | 1 | 2 | 4 | 2 | 0 | 0 | 1 | 1 | 2 | 1 | 1 | 0 | 1 |
| 3094723 | 1 | 3 | 3 | 3 | 4 | 2 | 1 | 2 | 4 | 2 | 0 | 0 | 1 | 1 | 2 | 1 | 1 | 0 | 1 |
| 3094724 | 0 | 1 | 3 | 5 | 4 | 5 | 2 | 2 | 4 | 1 | 0 | 0 | 1 | 1 | 2 | 1 | 1 | 0 | 1 |
| 3094725 | 1 | 5 | 3 | 2 | 4 | 2 | 1 | 1 | 4 | 1 | 0 | 0 | 1 | 1 | 2 | 1 | 1 | 0 | 1 |
| 3094726 | 0 | 4 | 3 | 2 | 4 | 2 | 1 | 1 | 4 | 1 | 0 | 0 | 1 | 1 | 2 | 1 | 1 | 0 | 1 |
| 3094727 | 1 | 3 | 3 | 3 | 4 | 2 | 1 | 2 | 4 | 1 | 0 | 0 | 1 | 1 | 2 | 1 | 1 | 0 | 1 |
| 3094728 | 0 | 1 | 3 | 4 | 4 | 2 | 2 | 1 | 4 | 1 | 0 | 0 | 1 | 1 | 2 | 1 | 1 | 0 | 1 |
| 3094729 | 0 | 3 | 3 | 4 | 4 | 2 | 1 | 1 | 4 | 2 | 0 | 0 | 1 | 1 | 2 | 1 | 1 | 0 | 1 |
| 3094730 | 1 | 3 | 3 | 3 | 4 | 2 | 1 | 2 | 4 | 2 | 0 | 0 | 1 | 1 | 2 | 1 | 1 | 0 | 1 |
| 3094731 | 0 | 3 | 3 | 2 | 4 | 2 | 1 | 2 | 4 | 2 | 0 | 0 | 1 | 1 | 2 | 1 | 1 | 0 | 1 |
| 3094732 | 1 | 2 | 3 | 2 | 4 | 2 | 1 | 1 | 4 | 2 | 0 | 0 | 1 | 1 | 2 | 1 | 1 | 0 | 1 |
| 3094733 | 0 | 2 | 3 | 2 | 4 | 2 | 2 | 2 | 4 | 1 | 0 | 0 | 1 | 1 | 2 | 1 | 1 | 0 | 1 |
| 3094734 | 0 | 2 | 3 | 4 | 4 | 2 | 3 | 2 | 4 | 1 | 0 | 0 | 1 | 1 | 2 | 1 | 1 | 0 | 1 |
| 3094735 | 0 | 3 | 3 | 4 | 4 | 2 | 1 | 1 | 4 | 2 | 0 | 0 | 1 | 1 | 2 | 1 | 1 | 0 | 1 |
| 3094736 | 0 | 3 | 3 | 4 | 4 | 2 | 1 | 1 | 4 | 1 | 0 | 0 | 1 | 1 | 2 | 1 | 1 | 0 | 1 |
| 3094737 | 0 | 4 | 3 | 3 | 4 | 2 | 2 | 1 | 4 | 4 | 0 | 0 | 1 | 1 | 3 | 1 | 1 | 0 | 1 |
| 3094738 | 0 | 3 | 3 | 3 | 4 | 2 | 1 | 1 | 4 | 2 | 0 | 0 | 1 | 1 | 2 | 1 | 1 | 0 | 1 |
| 3094739 | 0 | 3 | 3 | 2 | 4 | 2 | 1 | 2 | 4 | 2 | 0 | 0 | 1 | 1 | 4 | 1 | 1 | 0 | 1 |
| 3094740 | 0 | 3 | 3 | 4 | 4 | 2 | 2 | 1 | 4 | 2 | 1 | 0 | 1 | 1 | 1 | 1 | 0 | 0 | 0 |
| 309481 | 1 | 2 | 3 | 3 | 4 | 2 | 1 | 1 | 2 | 1 | 0 | 0 | 1 | 1 | 2 | 1 | 1 | 0 | 1 |
| 309482 | 1 | 2 | 3 | 3 | 4 | 2 | 1 | 1 | 2 | 4 | 0 | 0 | 1 | 1 | 2 | 1 | 1 | 0 | 1 |
| 309483 | 0 | 2 | 3 | 5 | 4 | 5 | 1 | 2 | 2 | 1 | 0 | 0 | 1 | 1 | 3 | 1 | 1 | 0 | 1 |
| 309484 | 0 | 5 | 3 | 2 | 4 | 2 | 1 | 2 | 2 | 1 | 0 | 0 | 1 | 1 | 3 | 1 | 1 | 0 | 1 |
| 309485 | 0 | 2 | 3 | 4 | 4 | 2 | 2 | 2 | 2 | 1 | 0 | 0 | 1 | 1 | 2 | 1 | 1 | 0 | 1 |
| 309486 | 0 | 3 | 3 | 2 | 4 | 2 | 2 | 2 | 2 | 1 | 0 | 0 | 1 | 1 | 1 | 1 | 0 | 0 | 0 |
| 309487 | 0 | 3 | 3 | 2 | 4 | 2 | 1 | 2 | 2 | 1 | 0 | 0 | 1 | 1 | 3 | 1 | 1 | 0 | 1 |
| 309488 | 0 | 4 | 3 | 2 | 4 | 2 | 1 | 2 | 2 | 1 | 0 | 0 | 1 | 1 | 1 | 1 | 0 | 0 | 0 |
| 309489 | 0 | 3 | 3 | 2 | 4 | 2 | 2 | 1 | 2 | 1 | 0 | 0 | 1 | 1 | 2 | 1 | 1 | 0 | 1 |
| 3094810 | 0 | 3 | 3 | 2 | 4 | 2 | 1 | 1 | 2 | 2 | 0 | 0 | 1 | 1 | 3 | 1 | 1 | 0 | 1 |
| 3094811 | 0 | 4 | 3 | 2 | 4 | 2 | 1 | 2 | 2 | 2 | 0 | 0 | 1 | 1 | 3 | 1 | 1 | 0 | 1 |
| 3094812 | 1 | 5 | 3 | 2 | 4 | 2 | 1 | 2 | 2 | 2 | 0 | 0 | 1 | 1 | 2 | 1 | 1 | 0 | 1 |
| 3094813 | 1 | 3 | 3 | 2 | 4 | 2 | 1 | 1 | 2 | 1 | 0 | 0 | 1 | 1 | 1 | 1 | 0 | 0 | 0 |
| 3094814 | 1 | 2 | 3 | 2 | 4 | 2 | 2 | 1 | 2 | 1 | 0 | 0 | 1 | 1 | 3 | 1 | 1 | 0 | 1 |
| 3094815 | 1 | 2 | 3 | 4 | 4 | 2 | 1 | 2 | 2 | 1 | 0 | 0 | 1 | 1 | 2 | 1 | 1 | 0 | 1 |

| | | | | | | | | | | | | | | | | | | | |
|---|---|---|---|---|---|---|---|---|---|---|---|---|---|---|---|---|---|---|---|
| 3094816 | 0 | 2 | 3 | 2 | 4 | 2 | 1 | 2 | 2 | 2 | 0 | 0 | 1 | 1 | 2 | 1 | 1 | 0 | 1 |
| 3094817 | 1 | 2 | 3 | 2 | 4 | 2 | 1 | 1 | 2 | 2 | 0 | 0 | 1 | 1 | 3 | 1 | 1 | 0 | 1 |
| 3094818 | 1 | 1 | 3 | 2 | 4 | 2 | 1 | 1 | 2 | 2 | 0 | 0 | 1 | 1 | 3 | 1 | 1 | 0 | 1 |
| 3094819 | 1 | 3 | 3 | 2 | 4 | 2 | 1 | 2 | 2 | 2 | 0 | 0 | 1 | 1 | 3 | 1 | 1 | 0 | 1 |
| 3094820 | 0 | 2 | 3 | 2 | 4 | 2 | 1 | 1 | 2 | 2 | 0 | 0 | 1 | 1 | 2 | 1 | 1 | 0 | 1 |
| 3094821 | 1 | 3 | 3 | 4 | 4 | 2 | 1 | 2 | 2 | 2 | 0 | 0 | 1 | 1 | 3 | 1 | 1 | 0 | 1 |
| 3094822 | 1 | 5 | 3 | 2 | 4 | 2 | 1 | 2 | 2 | 1 | 0 | 0 | 1 | 1 | 2 | 1 | 1 | 0 | 1 |
| 3094823 | 0 | 4 | 3 | 2 | 4 | 2 | 1 | 1 | 2 | 1 | 0 | 0 | 1 | 1 | 2 | 1 | 1 | 0 | 1 |
| 3094824 | 1 | 5 | 3 | 2 | 4 | 2 | 1 | 2 | 2 | 2 | 0 | 0 | 1 | 1 | 2 | 1 | 1 | 0 | 1 |
| 3094825 | 1 | 4 | 3 | 5 | 4 | 2 | 2 | 2 | 2 | 2 | 0 | 0 | 1 | 1 | 2 | 1 | 1 | 0 | 1 |
| 309491 | 1 | 4 | 3 | 2 | 4 | 2 | 1 | 2 | 2 | 1 | 0 | 0 | 1 | 1 | 1 | 4 | 0 | 1 | 1 |
| 309492 | 0 | 2 | 3 | 2 | 4 | 2 | 1 | 1 | 2 | 4 | 0 | 0 | 1 | 1 | 4 | 1 | 1 | 0 | 1 |
| 309493 | 0 | 4 | 3 | 2 | 4 | 2 | 1 | 2 | 2 | 4 | 0 | 0 | 1 | 1 | 2 | 1 | 1 | 0 | 1 |
| 309494 | 0 | 5 | 3 | 2 | 4 | 2 | 1 | 1 | 2 | 3 | 0 | 0 | 1 | 1 | 3 | 1 | 1 | 0 | 1 |
| 309495 | 0 | 2 | 3 | 2 | 4 | 2 | 1 | 1 | 2 | 1 | 0 | 0 | 1 | 1 | 2 | 1 | 1 | 0 | 1 |
| 309496 | 1 | 3 | 3 | 5 | 4 | 5 | 2 | 1 | 2 | 1 | 1 | 1 | 1 | 1 | 1 | 1 | 0 | 0 | 0 |
| 309497 | 1 | 4 | 3 | 2 | 4 | 2 | 1 | 1 | 2 | 2 | 0 | 0 | 1 | 1 | 2 | 1 | 1 | 0 | 1 |
| 309498 | 0 | 1 | 3 | 2 | 4 | 2 | 1 | 1 | 2 | 4 | 0 | 0 | 1 | 1 | 3 | 1 | 1 | 0 | 1 |
| 309499 | 0 | 3 | 3 | 2 | 4 | 2 | 1 | 2 | 2 | 4 | 0 | 0 | 1 | 1 | 3 | 1 | 1 | 0 | 1 |
| 3094910 | 1 | 3 | 3 | 3 | 4 | 2 | 2 | 2 | 2 | 4 | 0 | 0 | 1 | 1 | 4 | 1 | 1 | 0 | 1 |
| 3094911 | 0 | 3 | 3 | 2 | 4 | 2 | 1 | 2 | 2 | 4 | 0 | 0 | 1 | 1 | 3 | 1 | 1 | 0 | 1 |
| 3094912 | 0 | 2 | 3 | 5 | 4 | 5 | 2 | 1 | 2 | 4 | 0 | 0 | 1 | 1 | 4 | 1 | 1 | 0 | 1 |
| 3094913 | 1 | 2 | 3 | 2 | 4 | 2 | 1 | 1 | 3 | 4 | 0 | 0 | 1 | 1 | 2 | 1 | 1 | 0 | 1 |
| 3094914 | 1 | 2 | 3 | 3 | 4 | 2 | 1 | 2 | 2 | 1 | 0 | 0 | 1 | 1 | 1 | 1 | 0 | 0 | 0 |
| 3094915 | 1 | 3 | 3 | 2 | 4 | 2 | 1 | 1 | 2 | 1 | 0 | 0 | 1 | 1 | 2 | 1 | 1 | 0 | 1 |
| 3094916 | 1 | 4 | 3 | 2 | 4 | 2 | 1 | 1 | 2 | 1 | 0 | 0 | 1 | 1 | 1 | 1 | 0 | 0 | 0 |
| 3094917 | 1 | 2 | 3 | 2 | 4 | 2 | 1 | 1 | 2 | 1 | 0 | 0 | 1 | 1 | 1 | 1 | 0 | 0 | 0 |
| 3094918 | 0 | 2 | 3 | 4 | 4 | 2 | 1 | 2 | 2 | 4 | 0 | 0 | 1 | 1 | 1 | 4 | 0 | 1 | 1 |
| 3094919 | 0 | 2 | 3 | 2 | 4 | 2 | 1 | 2 | 2 | 4 | 0 | 0 | 1 | 1 | 3 | 1 | 1 | 0 | 1 |
| 3094920 | 0 | 3 | 3 | 2 | 4 | 2 | 2 | 1 | 2 | 4 | 1 | 0 | 1 | 1 | 3 | 1 | 1 | 0 | 1 |
| 3094921 | 1 | 3 | 3 | 2 | 4 | 2 | 1 | 2 | 2 | 4 | 0 | 0 | 1 | 1 | 1 | 4 | 0 | 1 | 1 |
| 3094922 | 0 | 3 | 3 | 2 | 4 | 2 | 1 | 2 | 2 | 4 | 0 | 0 | 1 | 1 | 3 | 1 | 1 | 0 | 1 |
| 3094923 | 0 | 2 | 3 | 2 | 4 | 2 | 1 | 1 | 2 | 1 | 0 | 0 | 1 | 1 | 1 | 1 | 0 | 0 | 0 |
| 3094924 | 0 | 1 | 3 | 5 | 4 | 5 | 3 | 1 | 2 | 4 | 0 | 0 | 1 | 1 | 3 | 1 | 1 | 0 | 1 |
| 3094925 | 0 | 2 | 3 | 5 | 4 | 5 | 1 | 2 | 2 | 4 | 0 | 0 | 1 | 1 | 3 | 1 | 1 | 0 | 1 |
| 3094926 | 1 | 4 | 3 | 4 | 4 | 2 | 2 | 1 | 2 | 1 | 0 | 0 | 1 | 1 | 1 | 1 | 0 | 0 | 0 |
| 3094927 | 1 | 4 | 3 | 2 | 4 | 2 | 2 | 1 | 2 | 1 | 0 | 0 | 1 | 1 | 1 | 4 | 0 | 1 | 1 |
| 3094928 | 1 | 2 | 3 | 5 | 4 | 5 | 2 | 1 | 2 | 1 | 1 | 1 | 1 | 1 | 1 | 1 | 0 | 0 | 0 |
| 3094929 | 1 | 1 | 3 | 2 | 4 | 2 | 1 | 1 | 2 | 1 | 0 | 0 | 1 | 1 | 2 | 1 | 1 | 0 | 1 |
| 3094930 | 1 | 3 | 3 | 2 | 4 | 2 | 1 | 2 | 2 | 1 | 0 | 0 | 1 | 1 | 2 | 1 | 1 | 0 | 1 |

***Variables explanation***

RID Respondent ID

G Gender of participants

- 0 Female
- 1 Male

AG Age group of participants

- 1 < 30
- 2 30 – 39
- 3 40 – 49
- 4 50 – 59
- 5 ≥ 60

D District in ENTP

1 East Sumba
2 Belu
3 East Manggarai

Edu Education level of participants

1 No education
2 Primary school
3 Junior high school
4 Senior high school'
5 Diploma or above

Eth Ethnicity in ENTP

1 Sumba
2 Others
3 Timor
4 Manggarai

Job Occupation of participants

1 Housewife
2 Farmer
3 Enterpreneur/Salesman
4 Other
5 Govt. or non-govt. employment

SES Social economic status of participants

1 Low
2 Average
3 High

FZ Family size

1 ≤ 4
2 > 4

HF The nearest health facilities

1 Village maternity posts
2 Village health Post
3 Subsidiary Public Health centres
4 Public Health centres

DHF Distance to the nearest health facilities

1 < 1 Km
2 1 – 2 Km
3 2 – 3 Km
4 ≥ 3 Km

Inc1 Categori of Household Income based on Minimum Wages Province

0 Less than the provincial minimum wages (PMW)
1 At least PMW

Inc2 Categori of Household Income based on Minimum Wages Province

0 Less than the poverty line of Indonesia
1 At least the poverty line of Indonesia

Q1 Hearing malaria term

0 No
1 Yes

Q2 Seeking treatment if participants or their family members have symptoms of malaria

0 No
1 Yes
99 Not applicable

Q3 How fast finding treatment if malaria

1 One day (Within 24 hours)

2 2 days

3 3 days

4 4 days or more

5 I did not go for treatment

99 Not applicable

Q4 Place for seeking treatment if participants or their family members have symptoms of malaria

1 Public health facilities

2 Private health facilities

4 Traditional healer

5 Self-treatment

6 Buying medicine at kiosk

7 Self-treatment with consuming papaya leaves

99 Not applicable

O1 Seeking treatment after 24 hours

0 No

1 Yes

99 Not applicable

O2 Seeking treatment at non-health facilities

0 No

1 Yes

99 Not applicable

O3 Poor understanding of appropriate malaria treatment-seeking behaviour (AMTSB)
(Seeking treatment after 24 hours or at non-health facilities)

0 No

1 Yes

99 Not applicable
